# A unified framework for estimating country-specific cumulative incidence for 18 diseases stratified by polygenic risk

**DOI:** 10.1101/2023.06.12.23291186

**Authors:** Bradley Jermy, Kristi Läll, Brooke Wolford, Ying Wang, Kristina Zguro, Yipeng Cheng, Masahiro Kanai, Stavroula Kanoni, Zhiyu Yang, Tuomo Hartonen, Remo Monti, Julian Wanner, Omar Youssef, Estonian Biobank research team, FinnGen, Christoph Lippert, David van Heel, Yukinori Okada, Daniel L. McCartney, Caroline Hayward, Riccardo E. Marioni, Simone Furini, Alessandra Renieri, Alicia R. Martin, Benjamin M. Neale, Kristian Hveem, Reedik Mägi, Aarno Palotie, Henrike Heyne, Nina Mars, Andrea Ganna, Samuli Ripatti

## Abstract

Polygenic Scores (PGSs) offer the ability to predict genetic risk for complex disease across the life course; a key benefit over short-term prediction models. To produce risk estimates relevant for clinical and public health decision making, it is important to account for any varying effects due to common risk factors such as age and sex. Here, we develop a novel framework to estimate for cumulative incidences over the life course and produce country-, age-, and sex-specific estimates of cumulative incidence stratified by PGS for 18 high-burden diseases by integrating PGS associations from 7 studies in 4 countries (N=1,197,129) with disease incidences from the Global Burden of Disease. PGSs had a significant sex-specific effect for 5 diseases (asthma, hip osteoarthritis, gout, coronary heart disease, type 2 diabetes) with all but type 2 diabetes exhibiting a larger effect in men. PGS had a larger effect in younger individuals for 13 diseases, with the effects decreasing linearly with age. We showed for breast cancer that, relative to individuals in the bottom 20% of polygenic risk, the top 5% attain an absolute risk for screening eligibility 16.3 years earlier. For T2D, men and women in the top 1% reached the threshold aged 24.8 (95% CI: 22.5 – 27.6) and 22.3 (95% CI: 20.0 – 25.3) respectively. Individuals in the bottom 1% of PGS did not reach the risk threshold by age 80. Our easily extendable framework increases the generalizability of results from biobank studies and the accuracy of absolute risk estimates by appropriately accounting age and sex-specific PGS effects. Our results highlight the potential of PGS as a screening tool which may assist in the early prevention of common disease.

## Introduction

Clinical calculators are often used to estimate disease risk in common diseases to facilitate early identification and primordial prevention. For example, QDiabetes (1) (https://qdiabetes.org/), and the Pooled Cohort Equations (PCE) estimate risk for type 2 diabetes (T2D) and Atherosclerotic Cardiovascular Disease, respectively (2).

Polygenic scores (PGSs) use combined information from a person’s genome to estimate their genetic risk of developing a specific disease or trait (3). Most of the predictive ability of PGS is obtained by summing thousands of common genetic variants of small effect, but PGS can also incorporate rare genetic variants with large effect (4). There is extensive discussion about the clinical and public health value of PGSs (5–7) with varying value on short term prediction when integrated on top of existing clinical prediction models for cardiovascular diseases and prostate cancer (8–10). Other authors have highlighted how PGSs provide independent, and therefore complementary, information about disease risk compared to many key risk factors included in prediction models such as family history (7,11). PGSs have been shown to associate strongly with many diseases and stratify individuals based on their genetic risks (12,13). Further, their impact on global disease burden, as measured by disability adjusted life years (DALYs), is comparable to well-established modifiable risk factors (14).

Maybe the most attractive feature of PGSs is that they can be calculated at birth, allowing for risk estimation in younger individuals not typically targeted by current disease risk calculators (15,16). The static nature of PGS means risk can be computed over the lifetime using a single genetic test for many diseases simultaneously, making them a potentially cost-effective prediction tool (6). On the contrary, current clinical calculators provide the absolute risk of disease over a short time frame, typically the next 5 or 10 years and are only applicable within a limited age range (1,2), which may partially explain why such calculators typically fail to identify high risk individuals with early-onset disease causing the biggest burden to the society and for the affected individual (17).

Thus, PGSs are a potentially useful tool in overcoming the limitations of short-term risk prediction and are well suited to provide lifetime absolute risk estimates. Such estimates must be comprehensively reviewed if they are to be used in personalized screening approaches. First, given countries may differ in terms of disease incidence and the discriminative ability of PGS, thus, we must understand how PGS generalizes across countries and health systems. A recent international study examining PGS association with 14 diseases across 7 countries only focused on relative risk (18). Second, while there is some evidence of a larger genetic contribution to early-onset disease cases (19–21), a detailed understanding of how risk estimation of PGS varies by both age and sex and how this translates to estimates of cumulative incidence is required to improve accuracy. Third, most biobank studies are not representative of the general population (22), with only few studies having attempted to recalibrate the impact of PGS on disease prevalence (23).

We address these three questions as part of the INTERnational consortium of integratiVE geNomics prEdiction (INTERVENE) (See Web Resources). We introduce a novel framework to allow for country-specific stratification opportunities for risk-based prevention and screening strategies. We demonstrate our method by combining incidences with polygenic risk associations across 7 studies in 4 countries (N=1,197,129) for 18 high-burden diseases (24). We demonstrate that for many diseases, PGSs stratify individuals into distinct risk trajectories over the lifetime with large differences in cumulative incidence between groups.

Our results also show that in many diseases the PGS effects are sex and age specific. To put our results into context and demonstrate the potential translational utility of our approach, we provide examples of how these results can be used for improving risk-based disease screening in different countries.

## Methods

### Participating studies in INTERVENE

Data from approximately 1.2 million participants of European ancestries were used across 7 studies - UK Biobank (UKB) (25), FinnGen (26), Estonian Biobank (EstBB) (27), Trøndelag Health Study (HUNT) (28), Generation Scotland (GS) (29), Genomics England (GE) (30), and Mass General Brigham Biobank (MGB) (31). Each contributing study performed genotyping, imputation, variant quality control and ancestry assignment using their own methodology (Supplementary Methods).

### Disease selection

Diseases were selected according to their global burden as defined by DALYs from the Global Burden of Disease (GBD) 2019 (24), and the availability of GWAS summary statistics for the creation of PGS. Using these considerations, we selected 18 diseases contributing 17.87% of total global DALYs (Supplementary Table 1). These diseases contribute to 25.02% of total DALYs in high socio-demographic index countries, of which all studies included in this analysis are based.

### Phenotype harmonization

To harmonize disease phenotypes across studies, we used definitions curated by a team of clinical experts in FinnGen (26) (Supplementary Table 2). The presence of any ICD-9 and ICD-10 codes included within the FinnGen phenotype were used to define cases across the remaining studies (Supplementary Table 2). Controls were defined as individuals without the relevant ICD codes for the disease.

All data used to define disease phenotypes was registry based. Missingness within registry data may result from either incomplete follow-up or a diagnosis being received in a health care system not included within the registry data, i.e. primary care. For these reasons, it is difficult to quantify missingness for registry data, however, a comprehensive overview of each studies’ registry information is provided in the supplementary methods.

A key step in the estimation of cumulative incidence is in calculating the baseline hazard which requires reference statistics from a nationally representative sample. We used incidence, prevalence and mortality estimates from the GBD for this step. To quantify the degree of overlap between the phenotypes defined from GBD and our disease definitions, and therefore justify the baseline hazard for each disease, we computed the percentage overlap of ICD codes across the two definitions. The number of records for each ICD code was extracted from the UKB data showcase (https://biobank.ndph.ox.ac.uk/showcase/). Overlap was high, with 10 diseases having a 100% overlap, and 14 having above 95% overlap (Supplementary Table 3). We originally considered diseases with 70% phenotype overlap which included interstitial lung disease (ILD). However, the GBD baseline hazard estimates from GBD were highly heterogeneous and unrealistic for a generally rare disease. As such, it was decided that all phenotypes were suitable for baseline hazard estimation using GBD. While the overlap in ICD codes for Major Depressive Disorder was 100%, further inspection suggests a liberal definition is used by the GBD where individuals only need to have suffered either of the two cardinal symptoms of MDD (depressed mood or anhedonia) over a two week period (32). To reflect the fact this is not a clinical diagnosis, we use the term depression throughout the rest of the paper.

### Estimating polygenic scores (PGSs)

For each phenotype, we searched for the summary statistics from the GWAS with the greatest sample size that was publicly available within GWAS Catalog (Supplementary Table 4). Biobank and trait combinations were only studied if independent from the GWAS contributing studies.To enhance analytical consistency and ensure variants were of high quality, we used single nucleotide polymorphisms (SNPs) in the intersection of HapMap phase 3 SNPs (33) and the 1000 Genomes (34) with a minor allele frequency greater than 1% in at least one super population (M=1,330,820). MegaPRS — a collection of PGS tools which allow the expected heritability contributed by each SNP to vary — was chosen for SNP weight calculation (35) as a previous methods comparison paper has shown it to have equal or superior prediction across a range of phenotypes (36) (opain.github.io/GenoPred). We selected the BLD-LDAK heritability model as this is recommended by the authors and used a data driven approach to tool and hyperparameter selection (we allow the data to find the best tool/hyperparameters through specifying the ‘mega’ argument). Following weight calculation, PLINK (37,38) was used to generate PGS for participants in each study. PGS in analyses were standardized to have mean 0 and variance 1 for each study.

### Survival analysis models

We performed ancestry-specific Cox Proportional-Hazards (PH) regression with age at disease onset as the timescale in each study. Follow-up started at birth and ended at the age of first record of a disease diagnosis (for individuals with the diseases), age at death for a cause other than the disease, age at last record available in the registries or electronic health records or age 80, whatever happened first. If the study was included in the base GWAS used for PGS calculation, the model was not tested with the exception of FinnGen and EstBB where relevant cohorts were excluded to remove sample overlap (Supplementary Table 4). In addition to the standardized PGS, the first 10 genetic principal components and study specific covariates used to control for technical artifacts (i.e. genotype batch, assessment center) were used as covariates.

### Sex and Age Stratification

Four separate Cox-PH models were tested for each phenotype: 1) Using the full sample (no stratification), 2) Sex-stratified, 3) Age-stratified and 4) Age and sex-stratified). We first computed hazard ratios (HRs) per standard deviation within each study. We then performed a fixed-effects meta-analysis on the log HRs across all studies tested to understand the generalizability of any age or sex specific effects. Studies were only included within the meta-analysis if it was possible to estimate a log HR in every strata, i.e. all age or sex strata.

To test for sex differences, an interaction term of PGS with sex was added to the model. In addition, Cox-PH models were repeated in each sex and HRs compared for any significant differences (Supplementary Methods).

To test for age-specific effects, we stratified each disease into four intervals, calculated according to the mean age at onset quartiles across FinnGen, HUNT, UKB and EstBB (Supplementary Table 5). We then performed separate Cox models in each interval using a method previously described in (21). Briefly, disease onset was only considered within the interval of a given quartile and participants were considered censored at the end of the interval if they had not died of a separate cause. If the participant had the disease in a prior interval, they were excluded from any follow-up intervals.

When deciding upon the optimal model:

a) Sex-specific effects were chosen if:

1. The meta-analyzed interaction effect (PGS*Sex) was significant (p < 2.8x10^-3^, details below)
b) Age-specific effects were chosen if:

1. There was a significant heterogeneity across the four quartiles, estimated using a Cochran’s Q test (39).
c) Age and sex-specific effects were chosen if:

1. Separate age and sex specific effects were found in both tests a and b.
2. Age-specific effects were found in a single sex where not previously found.
3. Age-specific effects were found to differ significantly across sexes.

To test if the age-specific effects were significantly different between men and women, we compared the effect sizes (Beta_men_=Beta_women_, p-value < 0.05) for the weighted linear regression fit for log(HR) on age (Supplementary Methods).

In all instances, we use p < 2.8x10^-3^ as our significance threshold, which represents a Bonferroni correction of 18 tests (number of phenotypes). This is with the exception of testing age-specific effects across sexes where a nominal p-value threshold is used (p<0.05) due to the limited data available between the age quartiles and HR.

### Cumulative incidence estimation

To calculate cumulative incidence - defined as the cumulative probability of disease from birth up to age 80 accounting for the competing risk of death from other causes - country and sex-specific estimates of age-specific (5-year age groups) incidence, prevalence and mortality were extracted for each disease from the GBD 2019 (24).

Cumulative incidence was estimated using the method described in (40). Briefly, for each sex and age group, five year bins in the case of GBD, the disease incidence hazard for age group *[m, m+4]* was calculated as:

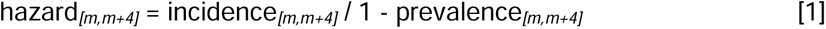

where incidence and prevalence values represent the number of new cases per year and the point prevalence assigned for the specific age group *[m, m+4], m={0,5,10,15,…75}.* The hazard for the age group therefore remains constant for all values within a given age group.

The hazard, in conjunction with the mortality rate due to other causes (overall mortality - cause specific mortality), was then used to calculate the probability of survival up to age *k*:

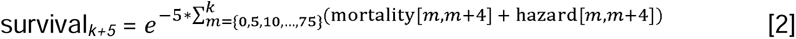

In equation 2, m increments in steps of 5 to correspond to the age groups specified above. The combined mortality and hazard were multiplied by 5 to account for the fact that the hazard and mortality reported values per year yet the age group covers 5 years. Survival is equal to 1 at age 0.

Similarly, the probability of a given disease from age *m to age m+4* was calculated as:

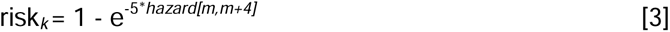

In equation 3, subscript k corresponds to the upper bound of the age group *[m, m+4].* Cumulative incidence was then calculated as the cumulative sum of survival at a given age multiplied by the probability of the disease at that age. For example, lifetime cumulative incidence is the cumulative sum until age 80:

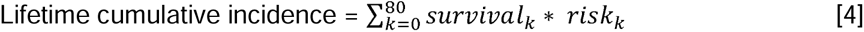

Cumulative incidence was calculated separately for each country (Estonia, UK, United States of America (USA), Norway and Finland) to ensure each study was calibrated to its population. As GE, GS and the UKB are all based in the UK, if association testing had been computed in more than one study, the HRs were meta-analyzed prior to computing cumulative incidence. As it was possible to estimate cumulative incidence at the state level, we calibrated the estimation to Massachusetts for MGB instead of using aggregated statistics for the USA.

To calculate cumulative incidence by PGS strata, it is necessary to group individuals according to their position in the PGS distribution. The default grouping was <20%, 20-40%, 40-60% (reference), 60-80%, 80-90%, 90-95%, >95%. Using these groupings, Cox-PH models were repeated in the full sample, and the sex, age, age and sex stratifications for each study.

Cumulative incidence for a given PGS group was calculated using the incidence estimates for the total population taken from the GBD and the HR from the relevant Cox-PH models. This method has been previously described in (41). Briefly, total incidence for a given age is equal to the weighted average of incidences for each PGS group. Baseline incidence (incidence rates for PGS group 40-60%) was computed as:

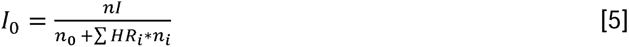

Where I0 is the baseline incidence, I is the age-specific total incidence across the population, n is the population size at a given age interval, ni the population size for the i-th PGS group and HRi is the HR for the ith PGS group.

The incidence attributable to an i-th PGS group was then estimated by:

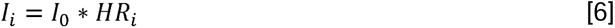

Incidences were then converted to probabilities of experiencing the disease using equations 1 and 3. Cumulative incidence for each PGS group was calculated using equations 1 through 4 with mortality due to other causes assumed to be equal across PGS groups.

To quantify the degree of uncertainty in the cumulative incidence estimation, we randomly sampled from the distribution of the error for each estimated parameter for both baseline and cox models and recalculated the cumulative incidence 1000 times. 95% confidence intervals were calculated by taking the 2.5 and 97.5 percentiles of the results.

To incorporate age-specific effects into the estimation of cumulative incidence, it was necessary to estimate HRs across the age span (0 to 80). For each disease with age-specific effects, a weighted linear regression was fit to the log HR estimates from the four age intervals with each estimate placed at the median age at onset for that interval. Predicted HRs from this regression were then incorporated into the cumulative incidence estimation. Ages outside of the range of the four HRs were assumed constant to the HR closest in age (Supplementary Figure 1).

### Translating estimates of cumulative incidence to other countries

To evaluate the impact of HR heterogeneity on the estimates of cumulative incidence, we meta-analyzed HRs for each PGS percentile group and re-evaluated cumulative incidence in each country using the country-specific baseline hazard. We then compared the cumulative incidence estimates for the study specific HRs to the meta-analyzed HRs.

### Sensitivity Analyses

#### The impact of relatedness on PGS association - full cohort vs unrelated subset

For EstBB and FinnGen, Plink v2 (37) and KING (42) were used to identify all related individuals up to degree 3 respectively. Individuals were then excluded as to remove relatedness from the sample while retaining the maximal sample size. HRs resulting from the full sample were then compared to the unrelated subset.

#### Robustness of registry-based disease definitions - primary care vs secondary care

Many diseases are first diagnosed in a primary care setting. To review the impact of not including this data, we created four disease definitions using Read v2 and CTV3 codes from the primary care data in the UKB (Supplementary Table 6). To increase consistency of our definitions to prior research, we used phenotype definitions from prior studies within the HDR UK Phenotype Library (https://phenotypes.healthdatagateway.org/). HRs calculated using the full sample were then and compared to associations from our main definitions which used secondary care data only.

#### The impact of the start of follow-up

To evaluate the impact of using age as the timescale in the analysis, we tested two different follow-up times within the UKB. Firstly, we tested using age at recruitment as the start of follow-up, equivalent to only including incident cases. Secondly, we tested age at registry linkage. This was calculated by assuming the birth country remained the country at which the registry was first linked. If the individual was born in Wales the date of registry linkage was 1st January 1998, in Scotland the date was set as 1st January 1981 and if born anywhere else, the date was set as 1st January 1997 (the date in which English registries were linked). All other aspects of the main analysis were the same with the exception that year of birth was added as a covariate.

### Ethics Statement

Patients and control subjects in FinnGen provided informed consent for biobank research, based on the Finnish Biobank Act. Alternatively, separate research cohorts, collected prior the Finnish Biobank Act came into effect (in September 2013) and start of FinnGen (August 2017), were collected based on study-specific consents and later transferred to the Finnish biobanks after approval by Fimea (Finnish Medicines Agency), the National Supervisory Authority for Welfare and Health. Recruitment protocols followed the biobank protocols approved by Fimea. The Coordinating Ethics Committee of the Hospital District of Helsinki and Uusimaa (HUS) statement number for the FinnGen study is Nr HUS/990/2017.

The FinnGen study is approved by Finnish Institute for Health and Welfare (permit numbers: THL/2031/6.02.00/2017, THL/1101/5.05.00/2017, THL/341/6.02.00/2018, THL/2222/6.02.00/2018, THL/283/6.02.00/2019, THL/1721/5.05.00/2019 and THL/1524/5.05.00/2020), Digital and population data service agency (permit numbers: VRK43431/2017-3, VRK/6909/2018-3, VRK/4415/2019-3), the Social Insurance Institution (permit numbers: KELA 58/522/2017, KELA 131/522/2018, KELA 70/522/2019, KELA 98/522/2019, KELA 134/522/2019, KELA 138/522/2019, KELA 2/522/2020, KELA 16/522/2020), Findata permit numbers THL/2364/14.02/2020, THL/4055/14.06.00/2020, THL/3433/14.06.00/2020, THL/4432/14.06/2020, THL/5189/14.06/2020, THL/5894/14.06.00/2020, THL/6619/14.06.00/2020, THL/209/14.06.00/2021, THL/688/14.06.00/2021, THL/1284/14.06.00/2021, THL/1965/14.06.00/2021, THL/5546/14.02.00/2020, THL/2658/14.06.00/2021, THL/4235/14.06.00/2021, Statistics Finland (permit numbers: TK-53-1041-17 and TK/143/07.03.00/2020 (earlier TK-53-90-20) TK/1735/07.03.00/2021, TK/3112/07.03.00/2021) and Finnish Registry for Kidney Diseases permission/extract from the meeting minutes on 4^th^ July 2019.

The Biobank Access Decisions for FinnGen samples and data utilized in FinnGen Data Freeze 10 include: THL Biobank BB2017_55, BB2017_111, BB2018_19, BB_2018_34, BB_2018_67, BB2018_71, BB2019_7, BB2019_8, BB2019_26, BB2020_1, BB2021_65, Finnish Red Cross Blood Service Biobank 7.12.2017, Helsinki Biobank HUS/359/2017, HUS/248/2020, HUS/150/2022 § 12, §13, §14, §15, §16, §17, §18, and §23, Auria Biobank AB17-5154 and amendment #1 (August 17 2020) and amendments BB_2021-0140, BB_2021-0156 (August 26 2021, Feb 2 2022), BB_2021-0169, BB_2021-0179, BB_2021-0161, AB20-5926 and amendment #1 (April 23 2020)and it’s modification (Sep 22 2021), Biobank Borealis of Northern Finland_2017_1013, 2021_5010, 2021_5018, 2021_5015, 2021_5023, 2021_5017, 2022_6001, Biobank of Eastern Finland 1186/2018 and amendment 22 § /2020, 53§/2021, 13§/2022, 14§/2022, 15§/2022, Finnish Clinical Biobank Tampere MH0004 and amendments (21.02.2020 & 06.10.2020), §8/2021, §9/2022, §10/2022, §12/2022, §20/2022, §21/2022, §22/2022, §23/2022, Central Finland Biobank 1-2017, and Terveystalo Biobank STB 2018001 and amendment 25^th^ Aug 2020, Finnish Hematological Registry and Clinical Biobank decision 18^th^ June 2021, Arctic biobank P0844: ARC_2021_1001.

Ethics approval for the UK Biobank study was obtained from the North West Centre for Research Ethics Committee (11/NW/0382). UK Biobank data used in this study were obtained under approved application 78537.

The genotyping in Trøndelag Health Study and work presented in here was approved by the Regional Committee for Ethics in Medical Research, Central Norway (2014/144, 2018/1622, 2018/411492). All participants signed informed consent for participation and the use of data in research.

Ethical approval for the GS:SFHS study was obtained from the Tayside Committee on Medical Research Ethics (on behalf of the National Health Service)

The activities of the EstBB are regulated by the Human Genes Research Act, which was adopted in 2000 specifically for the operations of the EstBB. Individual level data analysis in the EstBB was carried out under ethical approval 1.1-12/624 from the Estonian Committee on Bioethics and Human Research (Estonian Ministry of Social Affairs), using data according to release application S22, document number 6-7/GI/16259 from the Estonian Biobank.

The informed consent process for the Genomics England 100,000 Genomes Project has been approved by the National Research Ethics Service Research Ethics Committee for East of England—Cambridge South Research Ethics Committee.

The analysis using Mass General Brigham Biobank is approved under IRB protocol 2022P001736.

## Results

### Descriptive statistics

Contributing biobank size ranged from 7,018 participants (after filtering to GP and hospital data consented individuals) in GS to 447,332 participants in UKB. GE had the youngest median age of recruitment with 26.3 years (28 Interquartile Range (IQR)) and GS had the oldest median age of recruitment with 57 years (17.6 IQR, Table 1). FinnGen had the longest follow-up with 62 years (19 IQR) and MGB had the shortest follow-up with 10 years (13 IQR). EstBB had the largest percent of female participants (66%) and HUNT had the least (53%).

**Table 1.** Descriptive statistics by study. Abbreviations: yrs - years, IQR - Interquartile Range *Estonian Biobank uses as baseline the start of National Health Insurance Fund data from 2003.

| Study | Sample size | Age of recruitment (yrs) median (IQR) | Maximum follow-up time across traits (yrs) median (IQR) | % Female | Ascertainment strategy |
| --- | --- | --- | --- | --- | --- |
| Estonia Biobank | 199 868 | 43.5 (25.5) | 17.7 (0)* | 65.5 | Population |
| FinnGen | 412 090 | 55.8 (26.7) | 62 (19) | 55.9 | Population & Hospital |
| Genomics England | 29 427 | 26.3 (28) | 29 (0) | 59 | Hospital |
| Generation Scotland | 7 018 | 57 (17.6) | 39.8 (0) | 56.4 | Population |
| HUNT | 69 715 | 37 (21.1) | 25 (18) | 52.9 | Population |
| Mass General Brigham Biobank | 39 036 | 51 (22) | 10 (13) | 55.1 | Hospital |
| UK Biobank | 447 332 | 58 (12) | 24 (0) | 54.2 | Population |

Of the 18 phenotypes of interest, cancer was generally the most common phenotype (prevalence range 9-47%) except for EstBB which had a 26% prevalence of depression and a 9% prevalence of cancer. The least common phenotype was type 1 diabetes (T1D) in EstBB, MGB, and UKB, melanoma in FinnGen and HUNT, and rheumatoid arthritis in GS and GE, all at less than 1% prevalence. (Supplementary Table 7) Across phenotype and biobanks, the oldest median age of onset was prostate cancer in EstBB and GS, lung cancer in FinnGen, and MGB, atrial fibrillation in GE and HUNT, and gout in UKB (Supplementary Table 7). Appendicitis had the earliest age of onset for all biobanks (range=23.7-55.7 years) except for FinnGen and UKB, which had the youngest age of onset for T1D at 12.9 years (19.2 IQR) and 55.7 years (16.13 IQR) respectively.

### Association between PGS and 18 diseases

All PGSs were significantly associated with 18 respective diseases with a HR for 1 standard deviation in the PGS ranging from 1.06 (95% CI: 1.05 - 1.07) for appendicitis to 2.18 (95% CI: 2.13 - 2.23) for T1D. We observed significant heterogeneity, as tested by Cochran’s Q test, in estimates of relative risk across studies, partially driven by the large sample size which allowed us to detect small, yet significant differences (Supplementary Figure 2; Supplementary Table 8). Two examples of large study heterogeneity were observed for coronary heart disease (CHD), which PGS had HRs per SD ranging from 1.13 (95% CI: 1.07 - 1.19) in GS to 1.41 (95% CI: 1.40 - 1.43) in FinnGen and T1D with HRs per SD ranging from 1.41 (95% CI: 1.17 - 1.69) in MGB to 2.37 (95% CI: 2.31 - 2.44) in FinnGen.

### Sex, age-specific effects

We identified significant interactions between disease-specific PGS and sex for 5 diseases (p<2.8x10^-3^; Supplementary Table 9). PGS had a larger effect for CHD, gout, hip osteoarthritis, and asthma in men whereas for type 2 diabetes (T2D) the effect was larger in women (Figure 1a; Supplementary Table 8). The change in PGS effect with age was particularly prominent. In total, significant heterogeneity across age quartiles were detected in 13 of 18 phenotypes (Cochran’s Q P-value < 2.8x10^-3^ for all cancers, appendicitis, asthma, atrial fibrillation, CHD, epilepsy, gout, depression, knee osteoarthritis, prostate cancer, rheumatoid arthritis, T1D and T2D) (Figure 1b; Supplementary Table 10). The decreasing effect of PGS was approximately linear with age (Supplementary Figure 3) and relatively consistent across studies (Supplementary Figure 2c). The differences in age effects was large for T1D where the PGS effect per standard deviation was 2.57 (95% CI: 2.47 - 2.68) in the youngest quartile (age < 12.6) and 1.66 (95% CI: 1.58 - 1.74) in the oldest quartile (age > 33.3).

**Figure 1.**
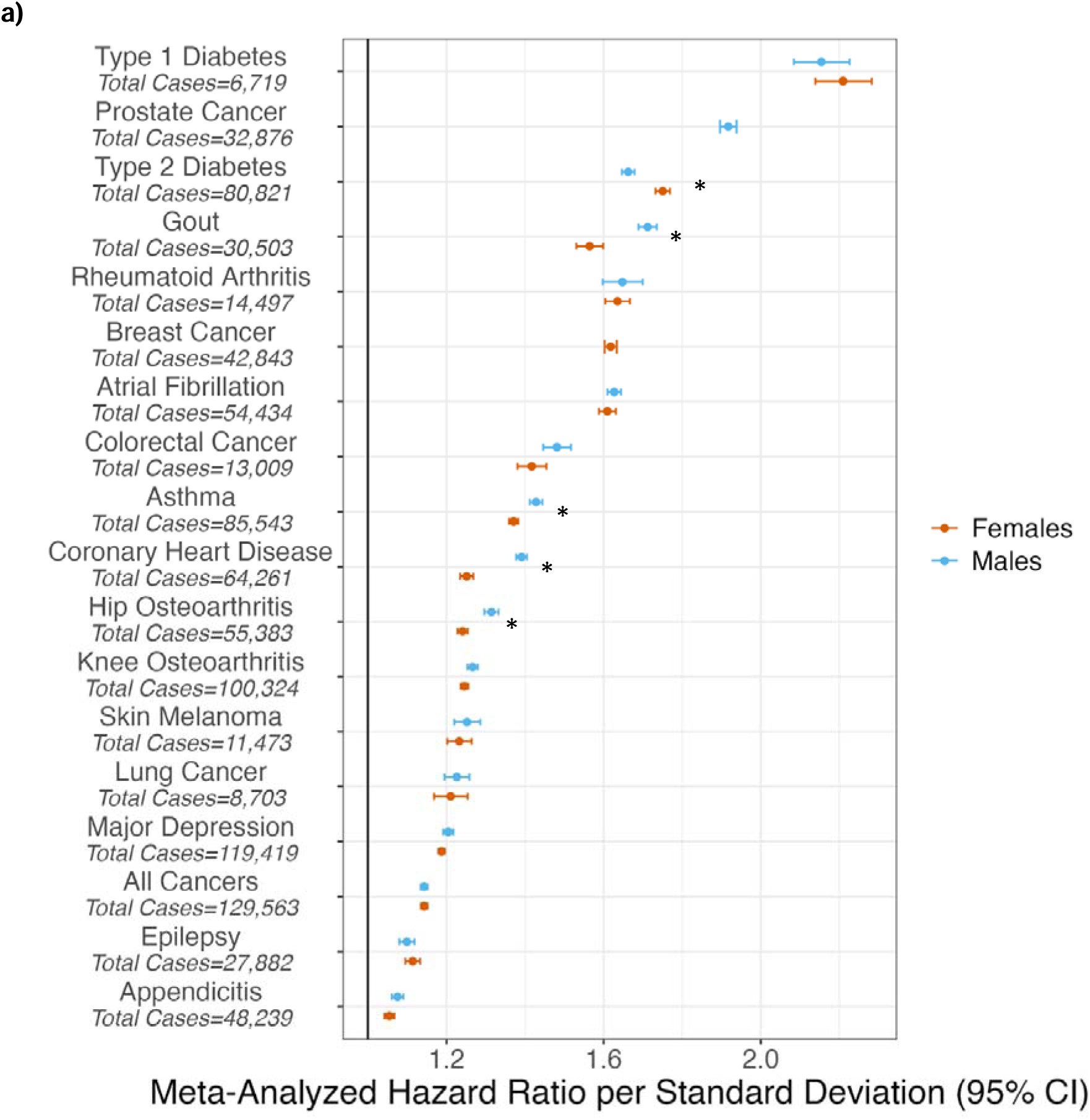
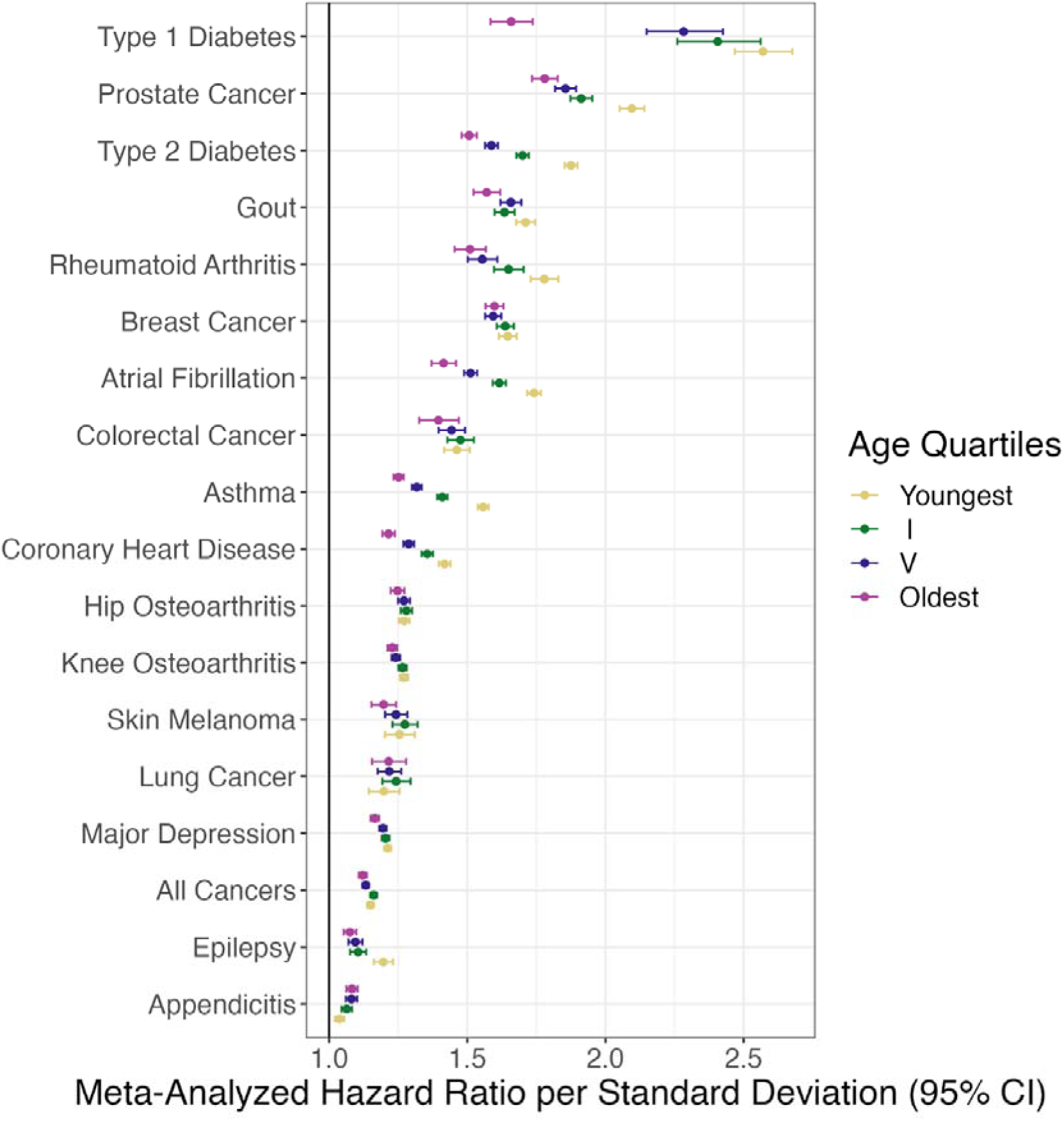
Model selection for each phenotype, stratified by ancestry. **a)** Meta-Analyzed hazard ratios per standard deviation stratified by sex. * indicates a significant interaction between PGS and Sex after Bonferroni correction for multiple testing. **b)** Meta-Analyzed hazard ratios per standard deviation stratified by age.

The large sample size allowed us to further examine the combined effect of both age and sex on PGS associations. One notable example was the association between PGS and CHD which decreased with age only in men, but not in women (P_het_ in men=1.05x10^-44^; P_het_ in women=0.04) (Supplementary Figures 4 and 5; Supplementary Tables 8 and 11).

Further examining the association between PGS and 18 diseases by PGS quantiles (Figure 2) we identified that for some diseases, age-specific effects were larger among individuals belonging to the tails of the PGS distribution. For example, individuals in top 5% of a PGS for prostate cancer vs those in the 40%-60% reference group had significantly higher relative risk for prostate when the disease was diagnosed at younger age (age < 62.6) HR=5.01 (95% CI: 4.65 - 5.39) compared to oldest ages (age > 73.9) HR=3.27 (95% CI: 2.97 - 3.60).

**Figure 2.**
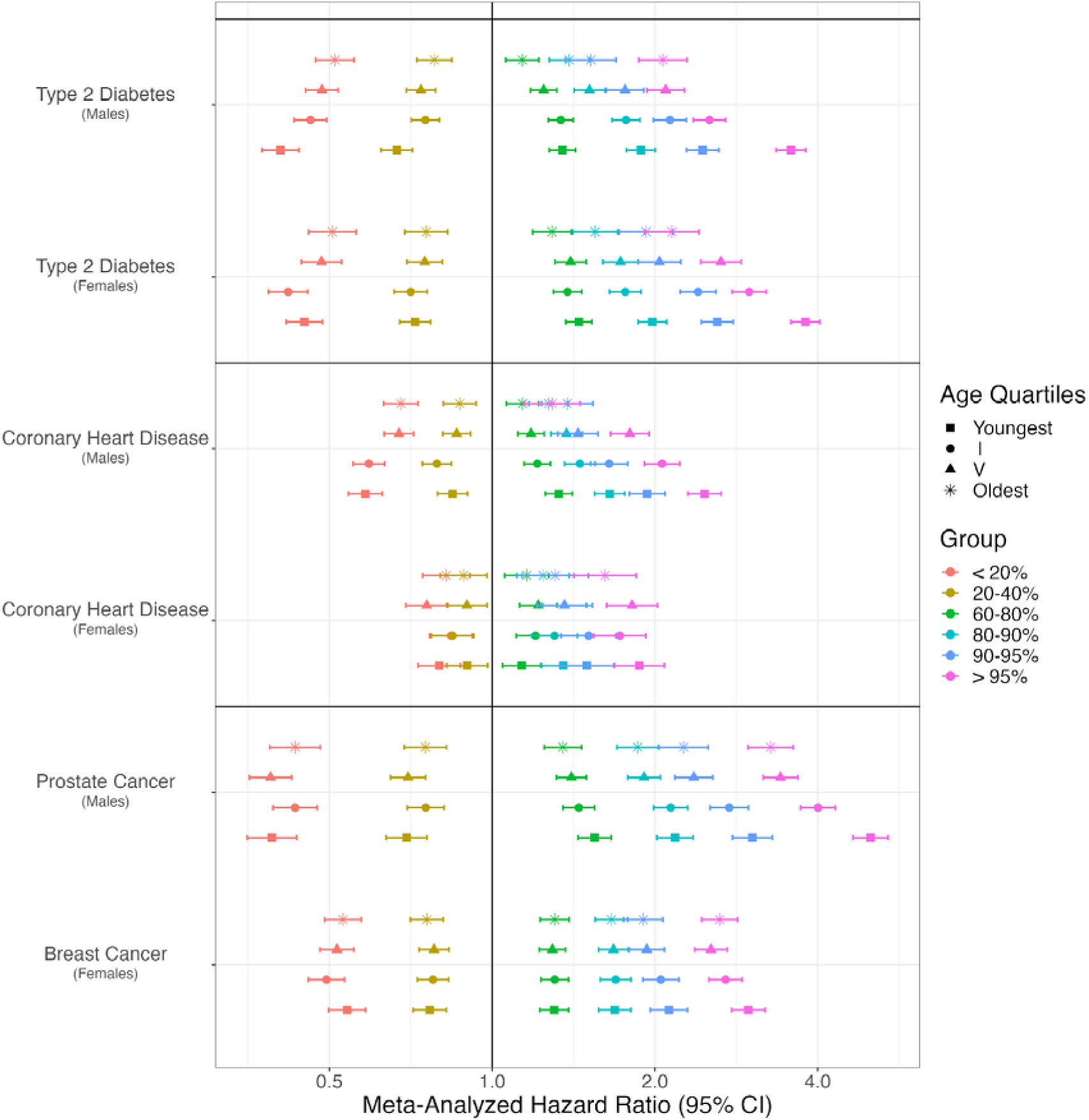
Meta-analyzed hazard ratios stratified by age and sex for Type 2 Diabetes, Coronary Heart Disease, Prostate Cancer and Breast Cancer.

### Country-specific cumulative incidence estimation stratified by PGS

For each disease, we derived country-specific estimates of the cumulative incidence by PGS quantiles, accounting for age and sex-specific effects and calibrating the baseline risk using GBD. Supplementary Table 12 highlights the final models to be employed in the estimation of cumulative incidence.

Variation in cumulative incidence was evident by PGS quantiles, country and sex with the main driver of the difference between country and sex being difference in baseline disease risk (Figure 3; Supplementary Figure 6; Supplementary Table 13). For example, in Massachusetts, the cumulative incidence at age 80 for CHD was significantly greater in men in the top 5% of PGS compared to the bottom 20% quantile (27.2% [95%CI: 23.8% - 31.2%] vs 18.9% [95%CI: 16.7% - 21.2%]). In Estonia, absolute differences between the top 5% and bottom 20% PGS quantiles were larger (72.9% [95%CI: 66.6% - 77.8%] vs 47.5% [95%CI: 41.6% - 53.4%]) due to overall higher incidence. In women, the absolute difference for the same PGS quantiles was lower than in men due to a decreased baseline risk (Massachusetts: 17.7% [95%CI: 15.3% - 20.2%] vs 11.3% [95%CI: 10.0% - 12.7%]); Estonia: 54.3% [95%CI: 47.2% - 60.5%] vs 43.6% [95%CI: 37.5% - 48.8%]). Similarly, for prostate cancer in the UK, the cumulative incidence was greater in men in the top 5% of PGS compared to the bottom 20% quantile (27.5% [95%CI: 23.4% - 31.3%] vs 3.7% [95%CI: 3.0% - 4.4%]). In Norway, the cumulative incidence was greater for the same PGS quantiles (35.4% [95%CI: 28.3% - 42.4%] vs 5.4% [95%CI: 4.0% - 7.2%]).

**Figure 3.**
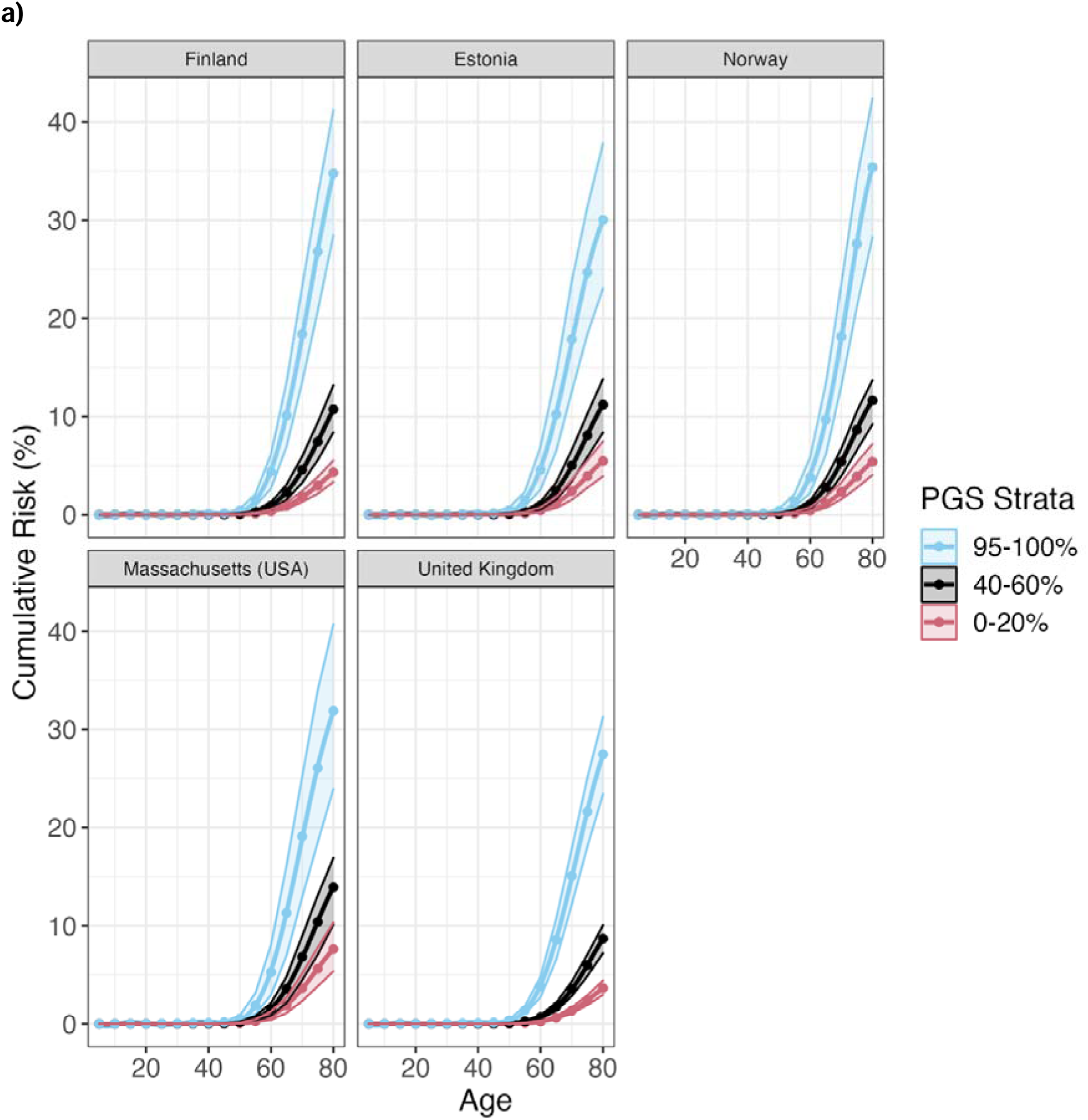
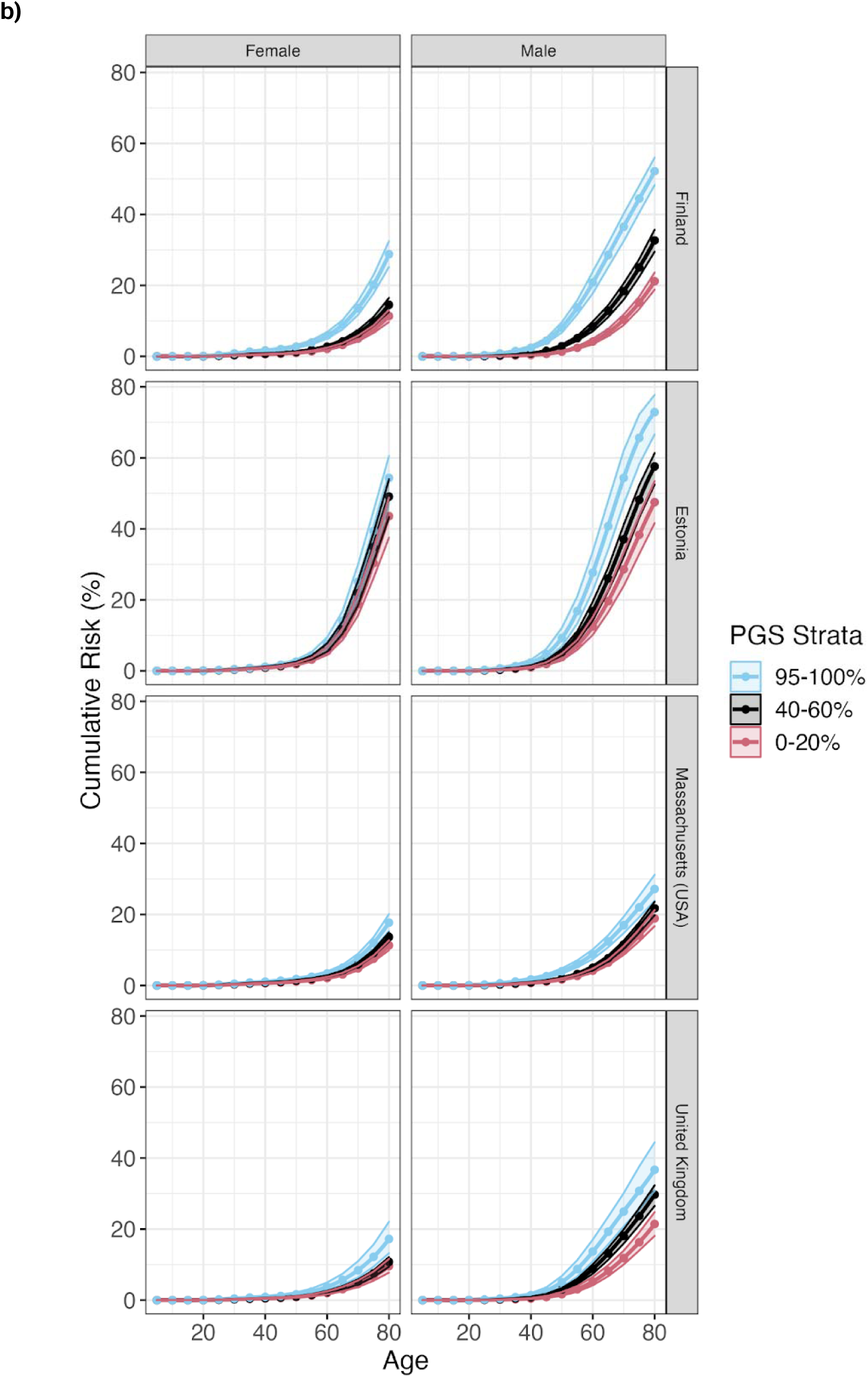
Country and sex-specific cumulative incidence estimates for the tails of the PGS distribution accounting for uncertainty. **a)** Prostate Cancer. **b)** Coronary Heart Disease.

### PGSs and disease screening

Our GBD calibrated country specific cumulative incidence estimates allowed us to illustrate the potential utility of risk based stratified screening for two diseases with existing screening recommendations: T2D and breast cancer. As mentioned earlier we found that the tails of the PGS distribution are particularly impacted by age-specific effects (Figure 2). This has direct relevance for screening strategies given clinical decisions regarding treatment will more likely occur in these groups. We, therefore, further explored the improvement on calibration that results from accounting for age- and sex-specific effects.

The American Diabetes Association recommends universal screening for T2D at age 45 with 3 year check-ups if the results of the screening are normal (43,44). We therefore estimated the country-specific cumulative incidence at age 45 prior to PGS stratification and used this as a clinical threshold (Supplementary Figure 7). Thresholds in cumulative incidence at 45 varied substantially across countries; ranging from 5.6% (Estonia) to 13.0% (UK) in men and from 4.7% (Norway) to 8.9% (UK) in women (Supplementary Table 14).

We estimated at which age the risk thresholds would have been reached as a function of PGS. Across studies, individuals in the bottom 20% of PGS would reach the risk threshold at an average age of 63.1 (95% CI: 58.8 - 67.3) whereas individuals in the top 5% would reach the same risk threshold at an average of 29.3 (95% CI: 26.2 - 32.9); a difference of 33.8 years (Supplementary Figures 6 and 8; Supplementary Table 15).

If age and sex-specific PGS effects were not accounted for in the calculation of the cumulative incidence, the ages at which the risk threshold was attained would be, on average, 2.8 years earlier in the bottom 20% and 1.8 years later in the top 5%.

For breast cancer, in many countries the initial screening is recommended to women at age 50 (45–50). In the countries examined by our study, the average cumulative incidence at age 50 ranged from 1.47% (Norway) to 2.05% (UK) (Supplementary Figure 9, Supplementary Table 16).

Women in the bottom 20% of PGS reached the risk threshold for breast cancer screening, on average, at age 58.7 (95% CI: 55.8 - 62.3) whereas women in the top 5% reached it at an average age of 42.9 (95% CI: 41.6 - 44.9); a mean difference of 15.8 years (Supplementary Figures 6 and 10; Supplementary Table 17).

To further illustrate the effects of the cumulative incidence differences at the tails of the PRS on potential risk based screening, we used the largest biobank study FinnGen to estimate the top/bottom 1% PRS cumulative incidences and calculated the ages at which the cumulative incidences were at the same level as in average person in the Finnish population at the start of the ADA screening for T2D (45 years) and national screening program for breast cancer (50 years). For T2D, Men and women in the top 1% reached the threshold aged 24.8 (95% CI: 22.5 – 27.6) and 22.3 (95% CI: 20.0 – 25.3) respectively. Individuals in the bottom 1% of PGS did not reach the risk threshold by age 80 (Figure 4a). For breast cancer women in the top 1% of PGS reached the threshold aged 42.3 (95% CI: 41 - 44.3) whereas women in the bottom 1% of PGS reached the threshold aged 66.3 (95% CI: 61.3 - 72.4); a difference of 24 years (Figure 4b).

**Figure 4.**
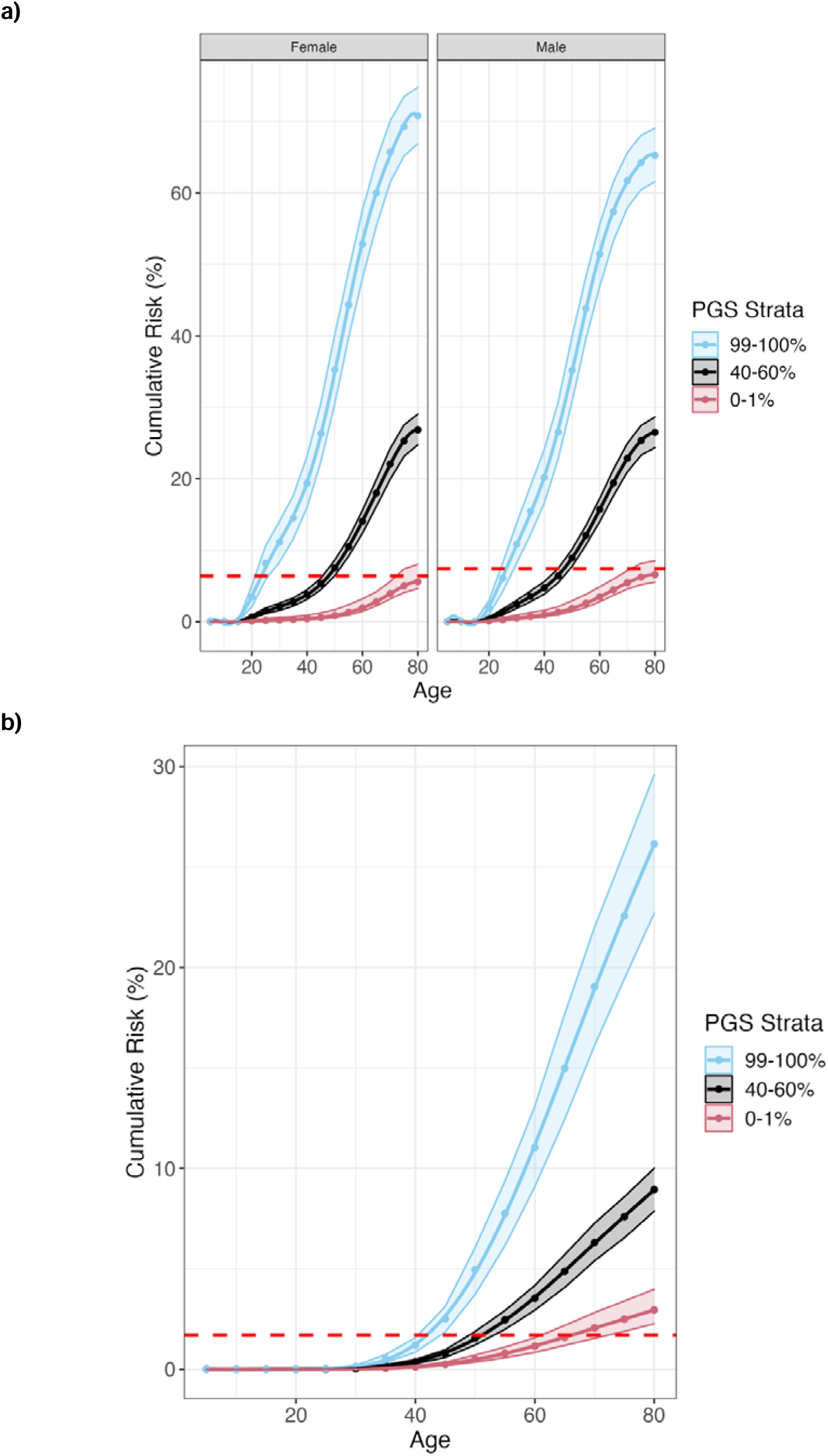
Sex-specific cumulative incidence estimates for Type 2 Diabetes and Breast Cancer in Finland. The red dashed line in each figure represents a country-specific clinically defined risk threshold for screening. **a)** Type 2 Diabetes cumulative incidence. **b)** Breast Cancer cumulative incidence.

### Translating to additional countries

In practice, most countries are unlikely to have studies with sufficient power to obtain robust associations between PGS and diseases. A possible solution is to use a pooled estimate from the meta-analysis across studies, however, heterogeneity in the HR across countries could limit the tools’ utility. Despite such heterogeneity, it appears meta-analyzed HRs are a good substitute. Using T2D and CHD as our examples, we first created a pooled estimate of the HR’s through meta-analyzing all estimates and recalculated cumulative incidence by combining these HRs with country-specific baseline hazards. In general, country-specific cumulative incidences using the meta-analyzed HRs were within the confidence intervals of our original estimates of cumulative incidence (Supplementary Table 18); indicating meta-analyzed HRs are a good substitute in the absence of country-specific data. Where differences did exist cumulative incidence was elevated when using meta-analyzed HRs. For type 2 diabetes, cumulative incidence was increased in the tails of the PGS distribution (top 5% and bottom 20%) in Massachusetts, as well as for Norwegians in the bottom 20% only (Supplementary Figure 11a). Similarly for CHD, cumulative incidence was elevated in Estonian women and men from Massachusetts in the top 5% of polygenic risk (Supplementary Figure 11b).

### Sensitivity analyses

First we determined that inclusion of related individuals did not impact the association between PGSs and diseases (Supplementary Figure 12). Second, we evaluated the robustness of our disease definitions based on primary care data from the UKB. Of the four phenotypes tested in the UKB (rheumatoid arthritis, epilepsy, gout, T1D), disease definitions using primary care data resulted in reduced HRs relative to the secondary care phenotypes. However, this difference was removed when adding in the criterion for each individual to have at least two codes (Supplementary Figure 13) - a common practice within primary care phenotyping to reduce misclassification (51). Combining primary and secondary care data tended to produce an association closer to the primary care only phenotype. Third, survival bias, potential induced by considering cases before study enrolment, does not seem to impact our results. Among six phenotypes considered in the UKB (gout, rheumatoid arthritis, prostate cancer, breast cancer, T1D, epilepsy), only prostate cancer PGS had a reduced relative risk when considering follow-up at baseline - equivalent of testing incident cases only - rather than birth (Supplementary Figure 14; Supplementary Table 19).

## Discussion

In this study we use data on 1.2 million study participants from 7 biobank studies to provide a broad overview on the impact of PGS on cumulative incidence of 18 diseases. We find evidence of considerable heterogeneity in the effect of PGS by both age and sex. We integrate such variation to reflect more accurate estimations of cumulative risk over the life course and highlight how PGS stratifies individuals and can impact risk-based screening practices for breast cancer and type 2 diabetes.

Our findings allow us to draw several conclusions. First, the heterogeneity in PGS effects across age shows that while our genetic profiles and PGS do not change with age, their impact on disease risk changes with age. A decreasing effect of PGS with age has been shown previously for some diseases (20,21,52) and our results confirm and expand those findings. In some diseases, environmental effects become more prevalent and variable with age (21), in effect reducing the heritability which represents the upper-bound of prediction from PGS. Our findings mirror those found in high-susceptibility genes, for example BRCA1 and BRCA2 mutations have been found to be associated with an earlier age at onset for breast cancer (53). Failing to account for the age-specific effects of PGS would underestimate disease risk in younger individuals.

Second, disease incidence is known to vary with sex for many diseases and this is mirrored by the sex differences in PGS effects (54–56). While there is some evidence of sex-specific effects at the genetic variant level (57–59), it is limited, possibly owing to the greater power requirements for an interaction effect. When combining thousands of genetic variants in a PGS, we show significant sex-specific associations for 5 diseases. Among these diseases, CHD and T2D have some previous evidence to support sex-specific PGS associations (60,61). There are many possible explanations for this differential effect. Different biological causal mechanisms could exist by sex. For example, lipids known to influence risk for CHD and T2D have been shown to vary by sex and age (62). Furthermore, the GWAS used to estimate the PGS may have an imbalance in the male:female ratio or sex-differential participation bias can induce differences in the phenotypic composition of males and females recruited in the study (64). Finally, diagnostic bias may mean one sex needs to have a higher disease liability before receiving a diagnosis. Understanding the root cause of these differences is important and can ultimately only be solved through the routine introduction of sex-specific GWAS.

Third, we found that while PGS tends to have a larger effect on diseases at younger ages, this effect might differ between males and females. For CHD, we observed a large PGS-age interaction effect only in males. Age at onset in CHD varies more in males than in females (24,65) and the risk factor profiles are known to change differently over age in males and females. Our results show that cumulative genetic effects captured by PGS are contributing to differences in age-related risks and may also be partly mediated by changes in risk factor profiles. In addition to CHD, we saw similar effects for gout which also has earlier age at onset for males (40,66). Ultimately, each disease will require its own assessment for age- and sex-specific effects. This study provides such a method for selecting the PGS estimate which balances power and accuracy in a systematic fashion, while also accounting for the importance of a harmonized phenotype across studies.

Arguably the best application of our country specific disease incidence and PGS estimates across large-scale biobanks is the development of country specific risk calculators that are based on lifetime risk of diseases which may be used to determine optimal ages for screening. Our framework is flexible in that it can integrate both country-specific and pooled PGS associations, depending on the requirement for specificity or generalizability respectively. This approach has multiple important advantages relative to the short-term prediction of most current models. Lifetime risk can be estimated early in life and overcomes the well documented challenge of short-term risk calculators for diseases not being useful in early life and therefore unable to enable primordial prevention approaches (17). As we illustrate with T2D and Breast Cancer, PGS provides lifetime risk trajectories that can enable stratified screening approaches. When available, other risk factors can easily be added when designing prediction models and screening approaches. Our approach also allows for developing country-specific risk calculators by utilizing baseline risk estimates derived from GBD, producing more accurate estimates for screening and overcoming the ascertainment biases inherent in many biobanks. We here demonstrate the risk estimation framework using 18 diseases, but risks can in principle be calculated for hundreds of diseases over the lifecourse at little additional expense (6).

Our study should be considered in light of the following limitations. First, the PGS used in this study do not consider the risk due to rare genetic variation (67). While this may reduce the accuracy of our risk estimates, at a population level, common genetic variation will be more predictive of the variation in complex disease which we are particularly well-powered to test in this study. As more sequencing studies become available, rare variants will also be included in the genome-wide risk estimation (4). Second, the use of harmonized phenotypes can ignore the country-specific nuances of ICD coding due to billing strategies. We deliberately focused on harmonization of the analysis to reduce bias due to technical variation, a rare feature in projects involving multiple studies. Third, our PGS relative risk are applicable to individuals of European ancestries only. As more studies with non-European individuals become available, similar estimates can be derived for a range of ancestries and admixtures. Fourth, some of the studies included are non-representative of the population due to sample recruitment strategies (volunteer- or hospital-based). While this may bias the cumulative risk estimation, a key strength of this study is the use of GBD to reduce the impact of selection bias in the baseline hazard which will bring our estimates closer to the true cumulative incidence.

In conclusion, we demonstrate the heterogeneity in polygenic score estimates between males and females and across lifespan in many diseases. While accounting for this heterogeneity, we developed a uniform framework to allow for estimation of life-time risk of diseases, stratified by individual’s genetic profiles and provide country-specific estimates. This information, which is already available for major modifiable risk factors (68,69), but was not yet comprehensively available for genetic scores, will allow health policy makers to better design screening tools with the goal to assist in the early prevention of common disease.

## Data Availability

Individual level data in this study are not publicly available due to legal and privacy limitations, but they can be accessed through individual participating biobanks. The FinnGen data may be accessed through Finnish Biobanks' FinBB portal (www.finbb.fi). The Trøndelag Health Study data may be accessed by application to the HUNT Research Centre. Researchers interested in Estonian Biobank can request the access at https://www.geenivaramu.ee/en/access-biobank. De-identified data of the MGB Biobank that supports this study is available from the MGB Biobank portal. Restrictions apply to the availability of these data, which are available to MGB-affiliated researchers via a formal application. UK Biobank data are available through a procedure described at http://www.ukbiobank.ac.uk/using-the-resource/. Genomics England data is available through an application process described here: https://www.genomicsengland.co.uk/research/academic/join-gecip. Generation Scotland data may be accessed through an application process described here: https://www.ed.ac.uk/generation-scotland/for-researchers/access
Summary level data are reported in tables and supplementary tables. The GWAS summary statistics used are publicly available and listed in Supplementary Table 4. The Global Burden of Disease data used here is available from http://ghdx.healthdata.org/.

## Acknowledgements

We want to acknowledge the participants and investigators of FinnGen study. The FinnGen project is funded by two grants from Business Finland (HUS 4685/31/2016 and UH 4386/31/2016) and the following industry partners: AbbVie Inc., AstraZeneca UK Ltd, Biogen MA Inc., Bristol Myers Squibb (and Celgene Corporation & Celgene International II Sàrl), Genentech Inc., Merck Sharp & Dohme LCC, Pfizer Inc., GlaxoSmithKline Intellectual Property Development Ltd., Sanofi US Services Inc., Maze Therapeutics Inc., Janssen Biotech Inc, Novartis AG, and Boehringer Ingelheim International GmbH. Following biobanks are acknowledged for delivering biobank samples to FinnGen: Auria Biobank (www.auria.fi/biopankki), THL Biobank (www.thl.fi/biobank), Helsinki Biobank (www.helsinginbiopankki.fi), Biobank Borealis of Northern Finland (https://www.ppshp.fi/Tutkimus-ja-opetus/Biopankki/Pages/Biobank-Borealis-briefly-in-English.aspx), Finnish Clinical Biobank Tampere (www.tays.fi/en-US/Research_and_development/Finnish_Clinical_Biobank_Tampere), Biobank of Eastern Finland (www.ita-suomenbiopankki.fi/en), Central Finland Biobank (www.ksshp.fi/fi-FI/Potilaalle/Biopankki), Finnish Red Cross Blood Service Biobank (www.veripalvelu.fi/verenluovutus/biopankkitoiminta), Terveystalo Biobank (www.terveystalo.com/fi/Yritystietoa/Terveystalo-Biopankki/Biopankki/) and Arctic Biobank (https://www.oulu.fi/en/university/faculties-and-units/faculty-medicine/northern-finland-birth-cohorts-and-arctic-biobank). All Finnish Biobanks are members of BBMRI.fi infrastructure (www.bbmri.fi). Finnish Biobank Cooperative -FINBB (https://finbb.fi/) is the coordinator of BBMRI-ERIC operations in Finland. The Finnish biobank data can be accessed through the Fingenious^®^ services (https://site.fingenious.fi/en/) managed by FINBB.

Generation Scotland received core support from the Chief Scientist Office of the Scottish Government Health Directorates [CZD/16/6] and the Scottish Funding Council [HR03006] and is currently supported by the Wellcome Trust [216767/Z/19/Z]. Genotyping of the GS:SFHS samples was carried out by the Genetics Core Laboratory at the Edinburgh Clinical Research Facility, University of Edinburgh, Scotland and was funded by the Medical Research Council UK and the Wellcome Trust (Wellcome Trust Strategic Award “STratifying Resilience and Depression Longitudinally” (STRADL) Reference 104036/Z/14/Z).We thank participants and scientists involved in making the UK Biobank resource available (http://www.ukbiobank.ac.uk/).

The Massachusetts General Brigham Biobank is described further here: https://www.massgeneralbrigham.org/en/research-and-innovation/participate-in-research/biobank.

The Trøndelag Health Study (HUNT) is a collaboration between HUNT Research Center (Faculty of Medicine and Health Sciences, Norwegian University of Science and Technology NTNU), Trøndelag County Council, Central Norway Regional Health Authority, and the Norwegian Institute of Public Health.

Estonian biobank was funded by European Union through the European Regional Development Fund Project No. 2014-2020.4.01.15-0012 GENTRANSMED. Data analysis was carried out in part in the High-Performance Computing Center of University of Tartu.

Estonian Biobank research team received funding from Estonian Research Council grant TT17 "Estonian Centre for Genomics".

This research was made possible through access to the data and findings generated by the 100,000 Genomes Project. The 100,000 Genomes Project is managed by Genomics England Limited (a wholly-owned company of the Department of Health and Social Care). The 100,000 Genomes Project is funded by the National Institute for Health Research and NHS England. The Wellcome Trust, Cancer Research UK and the Medical Research Council have also funded research infrastructure. The 100,000 Genomes Project uses data provided by patients and collected by the National Health Service (NHS) as part of their care and support. We acknowledge the contribution of the Genomics England Research Consortium to the 100,000 Genomes Project. The complete list of members of this Consortium can be found in the Supplementary Materials.

## Data availability

Individual level data in this study are not publicly available due to legal and privacy limitations, but they can be accessed through individual participating biobanks. The FinnGen data may be accessed through Finnish Biobanks’ FinBB portal (www.finbb.fi). The Trøndelag Health Study (HUNT). The HUNT data may be accessed by application to the HUNT Research Centre. Estonian Biobank. Researchers interested in Estonian Biobank can request the access at https://www.geenivaramu.ee/en/access-biobank. De-identified data of the MGB Biobank that supports this study is available from the MGB Biobank portal. Restrictions apply to the availability of these data, which are available to MGB-affiliated researchers via a formal application. UK Biobank data are available through a procedure described at http://www.ukbiobank.ac.uk/using-the-resource/. Genomics England data is available through an application process described here: https://www.genomicsengland.co.uk/research/academic/join-gecip. Generation Scotland data may be accessed through an application process described here: https://www.ed.ac.uk/generation-scotland/for-researchers/access

Summary level data are reported in tables and supplementary tables. The GWAS summary statistics used are publicly available and listed in Supplementary Table 4. The Global Burden of Disease data used here is available from http://ghdx.healthdata.org/.

## Contributor information

SR and AG conceived and designed the study. BJ, KL, and BW led meta-analysis, contributed to study design and planning, and wrote the manuscript with comments from all co-authors. BJ, KL, BW, YW, JZ, YC, MK, SK performed statistical and computational analyses. All authors provided critical inputs to interpretation of the data and approved the final version of the manuscript.

