## Supplementary Materials for "A unified framework for estimating country-specific cumulative incidence for 18 diseases stratified by polygenic risk"

### Supplementary Methods

**Study Specific Quality Control**

**UK Biobank**

***Registry data***

The relevant columns used to define the phenotypes were:

- Cause of Death Primary (Column ID: 40001)
- Cause of Death Secondary (Column ID: 40002)
- Summary ICD10 Diagnoses (Column ID: 41270)
- Summary ICD10 Diagnoses Date (Column ID: 41280)
- Summary ICD9 Diagnoses (Column ID: 41271)
- Summary ICD9 Diagnoses Date (Column ID: 41281)

Such data are taken from the hospital episode statistics which relate to hospital inpatient data. For more information, please see [here](https://biobank.ndph.ox.ac.uk/showcase/showcase/docs/HospitalEpisodeStatistics.pdf). Registry coverage depends on the country with follow-up beginning in 1997, 1998 and 1981 for England, Wales and Scotland respectively. End of follow-up was stated as 31st January 2021.

***Genotyping and quality control***

Two arrays were used to genotype UK Biobank participants. The UK Biobank Lung Exome Variant Evaluation (UKBiLEVE) Axiom array was used to genotype 49,950 participants. The remaining 438,427 participants were genotypes using the Applied Biosystems UK Biobank Axiom Array.

Principal Component Analysis (PCA) was performed on the genetic data and centralised quality control (QC) on variants was performed on individuals identified to belong to the largest cluster (N=463,844) according to Aberrant - an unsupervised clustering algorithm (1). Variants were assessed for evidence of allele frequency variation across batch, plate, sex or array and that genotypes were largely consistent with Hardy-Weinberg Equilibrium expectations (all p-value thresholds < 10^-12^). If a variant failed one or more tests within a given batch it was set to missing. See (2) for more detailed information on testing.

***Imputation***

For 487,442 individuals, imputation was performed using the IMPUTE4 (3) software. Genetic variation from the Haplotype Reference Consortium (HRC) (4) and merged UK10K+1000 Genomes (5) were used as a reference panel. Single Nucleotide Polymorphisms (SNPs) were only included in the final imputation if they were present in both reference panels, giving a total of 96,959,328 SNPs.

***Ancestry assignment***

Ancestry assignment uses methodology and scripts from GenoPred (<https://opain.github.io/GenoPred/DiverseAncestry.html>). Individuals were stratified into one of five super populations African (AFR), American (AMR), South Asian (SAS), East Asian (EAS) and European (EUR). The 1000 Genomes data (6) acted as a reference given the individuals are known to belong to one of the 5 super populations. Only unambiguous SNPs also present in both the HapMap3 consortium (7) and the imputed UK Biobank data were retained for PCA. SNPs within both the reference (1000 Genomes) and target (UK Biobank) samples underwent quality control such that the minor allele frequency (MAF) > 5%, variant missingness > 2% and Hardy-Weinberg Equilibrium p-value > 1e^-6^. 467,970 autosomal SNPs remained following QC and were in the intersection of the reference and target samples. Regions with long range linkage disequilibrium were excluded and independent SNPs (SNPs greater than 1000kb apart and r^2^ < 0.2) retained. PCA was then performed in the reference sample using PLINK v2 (8) and a multinomial elastic-net regression was trained using 5-fold cross validation, super population as the outcome and the first 10 PCs as covariates. PCs from the target sample were then projected into the reference space and prediction on super population made. Classifications were made according to the super population with the greatest probability. To be classified the max probability must be over 0.5, otherwise it was set to missing.

PCA was performed using a random subset of 1000 individuals per super population and PC’s from the rest of the super population sample projected onto this space. Distances from the centroid were calculated and outliers removed. Outliers were classified as having a distance > 75 percentile + 30*Interquartile Range. Following within-ancestry QC, 8,381, 1,063, 2,393, 447,332 and 9,435 individuals were allocated to AFR, AMR, EAS, EUR and SAS super populations respectively.

**FinnGen**

***Registry data***

Phenotype data within FinnGen is constructed from the collection of nationwide electronic health registers. This gives a comprehensive coverage of almost all of a patient's interactions with the health service including hospitalizations, medications, procedures and deaths. The 18 different registers used by the project are listed below in order of their follow-up times:

- [Finnish Cancer Registry](https://cancerregistry.fi/) - From 1953
- [Register of Congenital Malformations](https://thl.fi/en/web/thlfi-en/statistics-and-data/data-and-services/register-descriptions/register-of-congenital-malformations) - From 1963
- [Reimbursement](https://raportit.kela.fi/ibi_apps/WFServlet?IBIF_ex=NIT137AL&YKIELI=E) - From 1964
- [Population Register](https://dvv.fi/en/population-information-system) - From 1964
- [Finnish Registry for Kidney Diseases](https://www.muma.fi/liitto/suomen_munuaistautirekisteri/finnish_registry_for_kidney_diseases) - From 1964
- [Causes of Death](https://www.stat.fi/til/ksyyt/index_en.html) - From 1969
- [Care Register for Health Care Inpatient Visits, HILMO](https://thl.fi/en/web/thlfi-en/statistics-and-data/data-and-services/register-descriptions/care-register-for-health-care) - From 1969
- [Socio-economic data](https://taika.stat.fi/en/) - From 1970
- [The Finnish Registry of Visual Impairment](https://www.nkl.fi/en) - From 1983
- [Medical Birth Register](https://thl.fi/en/web/thlfi-en/statistics-and-data/data-and-services/register-descriptions/newborns) - From 1987
- [Finnish National Infectious Disease Register](https://thl.fi/en/web/infectious-diseases-and-vaccinations/surveillance-and-registers/finnish-national-infectious-diseases-register) - From 1989
- [Cervical Cancer Screening](https://cancerregistry.fi/screening/cervical-cancer-screening/) - From 1991
- [Breast Cancer Screening](https://cancerregistry.fi/screening/breast-cancer-screening/) - From 1992
- [Drug Purchases](https://www.kela.fi/kelas-research-and-statistics) - From 1995
- [The Care Register for Social Welfare](https://www.julkari.fi/bitstream/handle/10024/127104/Tr21_15.pdf?sequence=4&isAllowed=y) - From 1995
- [Care Register for Health Care, specialist outpatient visits, HILMO](https://thl.fi/en/web/thlfi-en/statistics-and-data/data-and-services/register-descriptions/care-register-for-health-care) - From 1998
- [Register of Primary Health Care Visits, Avohilmo](https://thl.fi/fi/tilastot-ja-data/ohjeet-tietojen-toimittamiseen/perusterveydenhuollon-avohoidon-hoitoilmoitus-avohilmo) - From 2011
- [The Finnish Vaccination Register](https://thl.fi/en/web/infectious-diseases-and-vaccinations/surveillance-and-registers/finnish-national-vaccination-register-and-monitoring-of-the-vaccination-programme) - From 2011

Note: while primary health care visits are included within FinnGen, by default these cases are excluded from the endpoints. As such, we only consider secondary care data for our disease endpoints.

***Genotyping and quality control***

FinnGen consists of prospectively recruited samples and a series of legacy cohorts with genotypes already available (9). Prospective samples were genotyped using the ThermoFisher Axiom custom array which tags a total of 655,973 variants. Genotype calling was performed using the Array Power Tools software. Legacy cohorts were genotyped using various Illumina arrays and genotype calling was performed using either GenCall or zCall algorithms.

For both prospective and legacy cohorts the following quality control metrics were used.

Samples were removed if:

- Pihat was > 0.9 and the samples were not monozygotic or replicates
- There was a discrepancy between reported sex and genetically determined sex (F-value ≤ 0.3 for females and ≥ 0.8 for males)
- Missingness was ≥ 5%
- Heterozygosity was ±4 standard deviations from the population average
- Pihat was > 0.1 with 14 or more samples
- Samples were ±4 standard deviations away from the population average according to the first two genetic principal components.

Samples were tagged should there be evidence of a mendelian error or contain replicate samples with over 50,000 discrepancies.

Variants were removed if:

- The variant failed the Hardy-Weinberg Equilibrium test (p-value < 10^-6^)
- The variant had a call rate < 98%

***Imputation***

Pre-phasing was performed using Eagle 2.3.5 (10) and samples were imputed using the SiSu v3 imputation reference panel. This reference panel is specific to the Finnish population, containing high-coverage (25-30x) whole-genome sequencing data from 3,775 Finns and 16,962,023 variants with minor allele count ≥ 3. After imputation, 16,387,711 variants were imputed with high quality (INFO > 0.6).

***Ancestry assignment***

Firstly, the FinnGen samples were combined with the 1000 genomes phase 3 dataset (6). Genetic principal components were calculated using a subset of 49,451 pruned SNPs. Aberrant (1) was used to identify and remove samples that deviated from the main cluster. A probability of belonging to either a North-Western European or Finnish population was calculated by firstly performing PCA with individuals belonging to these ancestries from 1000 genomes data. FinnGen samples were then projected onto this PCA space and Mahalanobis distances calculated for each sample against each of the two ancestries. Samples were retained if there was ≥ 95% probability of belonging to the Finnish ancestry cluster.

**Trøndelag Health Study**

***Registry data***

The periodic population-based health survey design includes three recruitment waves—HUNT1 (1984-1986), HUNT2 (1995-1997), and HUNT3 (2006-2008)—concentrated in the North-Trøndelag area, where all adults > 20 years of age were invited to participate. Electronic health records from the Trøndelag county hospitals (Nord-Trøndelag Hospital Trust, including St. Olavs, Namsos, and Levanger Hospitals) hold International Classification of Diseases and Related Health Problems (ICD) codes back to 1987 and were last accessed August 8, 2021. There is likely under-ascertainment of less-serious common conditions with only hospital records. Main and secondary diagnoses were used.

***Genotyping and quality control***

DNA from 71,860 HUNT samples was genotyped using one of three different Illumina HumanCoreExome arrays (HumanCoreExome12 v1.0, HumanCoreExome12 v1.1 and UM HUNT Biobank v1.0). These chips included custom content to directly genotyped missense and loss of function variants and lipid associated variants from low pass sequencing, among other potentially actionable protein altering variants. Samples that failed to reach a 99% call rate, had contamination > 2.5% as estimated with BAF Regress (11), large chromosomal copy number variants, lower call rate of a technical duplicate pair and twins, gonosomal constellations other than XX and XY, or whose inferred sex contradicted the reported gender, were excluded. Samples that passed quality control were analysed in a second round of genotype calling following the Genome Studio quality control protocol described elsewhere (12). Genomic position, strand orientation and the reference allele of genotyped variants were determined by aligning their probe sequences against the human genome (Genome Reference Consortium Human genome build 37 and revised Cambridge Reference Sequence of the human mitochondrial DNA; [http://genome.ucsc.edu](http://genome.ucsc.edu/)) using BLAT. Variants were excluded if their probe sequences could not be perfectly mapped to the reference genome, cluster separation was < 0.3, Gentrain score was < 0.15, showed deviations from Hardy Weinberg equilibrium in unrelated samples of European ancestry with p-value < 0.0001), their call rate was < 99%, or another assay with higher call rate genotyped the same variant.

***Imputation***

Imputation was performed on the 69,716 samples of recent European ancestry using Minimac3 (v2.0.1, <http://genome.sph.umich.edu/wiki/Minimac3>) (13) with default settings (2.5 Mb reference based chunking with 500kb windows) and a customized Haplotype Reference consortium release 1.1 (HRC v1.1) for autosomal variants and HRC v1.1 for chromosome X variants (4). The customized reference panel represented the merged panel of two reciprocally imputed reference panels: (1) 2,201 low-coverage whole-genome sequences samples from the HUNT study and (2) HRC v1.1 with 1,023 HUNT WGS samples removed before merging. We excluded imputed variants with Rsq < 0.3 resulting in over 24.9 million well-imputed variants.

***Ancestry assignment***

Ancestry of all samples was inferred by projecting all genotyped samples into the space of the principal components of the Human Genome Diversity Project (HGDP) reference panel (938 unrelated individuals; downloaded from <http://csg.sph.umich.edu/chaolong/LASER/>) (14), using PLINK v1.90 (8). Recent European ancestry was defined as samples that fell into an ellipsoid spanning exclusively European populations of the HGDP panel. The different arrays were harmonized by reducing to a set of overlapping variants and excluding variants that showed frequency differences > 15% between data sets, or that were monomorphic in one and had MAF > 1% in another data set. The resulting genotype data were phased using Eagle2 v2.3 (10).

**Estonian Biobank**

***Registry data***

Phenotype data within EstBB is put together from the collection of electronic health registers, including from two largest hospitals in Estonia. We include both primary and secondary care data as well as self-reported diagnoses. Estonia has a solidary health insurance system and national public health insurance covers ∼94% of the population (https://eurohealthobservatory.who.int/countries/estonia). Causes of death and Cancer registry record all cases despite the health insurance status in Estonia.  Following registries were included in phenotype definition process:

- [Causes of Death Registry](https://www.tai.ee/et/statistika-ja-registrid/surma-pohjuste-register)- diagnoses from 2003 until 2020
- [National Cancer Registry](https://www.tai.ee/et/statistika-ja-registrid/vahiregister)- diagnoses from 1955 until 2017
- Estonian Health Insurance Fund - From 2001 until 2020

### [The North Estonia Medical Centre](https://www.regionaalhaigla.ee/en) from 1993 until 2017

- Tartu University hospital from 2006 until 2017
- E-Health system from 1998 to 2020

Self-reported diagnoses’ dates ranged from 1920 - 2018.

***Genotyping and quality control***

Estonian BioBank (EstBB) samples were genotyped with 4 sub-versions of Infinium Global Screening Array-24. Samples with less than 95% call-rate were excluded. Sample sex recorded in EstBB database was compared with genetic sex. Samples with sex mismatch were further inspected for sex chromosome abnormalities (X0, XXY, etc.), and samples with confirmed database vs genetic sex mismatch were excluded. In total, 202 910 individuals passed sample quality control. SNP quality control was performed by excluding: (a) all SNPs with less than 95% call-rate, (b) SNPs showing more than 5% AF difference from the AF mean estimated using all genotyping batches with more than 10 000 samples per batch, (c) SNPs with Illumina GenTrain score < 0.6 or cluster separation score < 0.4 in any genotyping batch, (d) autosomal SNPs with HWE exact test p-value < 1e-4. In total, approximately 328K autosomal and X-chromosome SNPs with MAF > 1% passed quality control and were used in the imputation. All the variants were processed on the human genome assembly GRCh37.

***Imputation***

Imputation was performed using local Estonian imputation reference panel made of 2056 WGS samples. Genotypes were pre-phased with Eagle v2.4.1 (10) and imputed with Beagle 5.1 using default parameters (15). Multiallelic positions were excluded from imputation output. In total, 39 546 641 variants were used in the study.

***Ancestry assignment***

EstBB samples were combined with the 1000 genomes phase 3 dataset for ancestry analysis (6). Genetic principal components were calculated using a subset of quality controlled and pruned genotyped SNPs. This was further used to identify and remove samples that deviated from the main cluster via visual inspection. In total, 481 non-european ancestry individuals based on principal components were excluded from the analysis.

**Mass General Brigham Biobank**

***Registry data***

Patients and employers of multiple health centers at Mass General Brigham (MGB) in Eastern Massachusetts are enrolled in the MGB Biobank. The MGB Biobank was founded in 2008 and the research protocol was approved by the Human Research Committee of MGB. The EHR data were retrieved from the MGB Patient Data Registry (RPDR). A biobank portal which is an i2b2-based data repository linking disparate and high-dimensional patient data was implemented (16). The weekly updated repository integrates Information from primary and curated data resources. Data stored on the observational medical outcomes partnership (OMOP) can also be queried through i2b2.

***Genotyping and quality control***

To date, ~65,000 individuals with informed consent have generated and provided genomic information. In this study, we were focused on those individuals genotyped on the Illumina Global Screening Array. We retained genotyped SNPs that: i) minor allele count >= 2, ii) missingness <= 2% iii) Hardy-Weinberg equilibrium  *p*-value >= 1e^-6^, and iv) concordant allele frequency with gnomAD (chi-squared value >= 300). For individual-level quality controls, we removed those individuals that i) heterozygosity > 3 standard deviation of population mean, ii) missingness > 1% and ii) discordant sex between self-reported and genetic inferred sex. Finally, 563,449 genotyped variants for 52,459 individuals were subsequently imputed.

***Imputation***

The genotypes after quality controls were imputed at the Michigan imputation server using the TOPMed r^2^ imputation panel (17) using Minimac (18). Eagle v2.4 was used for haplotype phasing (10). Variants with imputation INFO scores > 0.3 were further retained for follow-up analyses.

***Ancestry assignment***

We projected all individuals onto the first 6 PCs based on 168,898 variants in the combined reference dataset from 1000 Genomes Project Phase 3 and Human Genome Diversity Project. We applied a random forest classifier and assigned ancestry to 6 continental groups (including European, Central and South-Asian, East-Asian, African, Middle-Eastern and American) if the probability was larger than 0.8.

**Genomics England**

***Registry data***

Available clinical data in the Genomics England research environment are divided into primary, sourced from the Genomic Medicine Centres for all participants upon enrollment in the program, and secondary clinical data come from third parties such as Public Health England or NHSD which complement the primary clinical data with additional information.

The following table was included in the phenotype definition process:

- **Hospital Episode Statistics admitted patient care** (hes_apc) - contains historic records of admissions into secondary care of Genomics England main programme participants. The period covered by the registry is from September 13, 1992, to January 31, 2022. The HES records are based on the ICD-10 coding system, which has been in use since April 1995. For more information please see here <https://re-docs.genomicsengland.co.uk/release15/>.

***Genotyping and quality control***

Genome sequencing was performed in DNA samples from 78,195 individuals using Illumina HiSeq X systems (150bp  paired-end format). Reads were aligned using the iSAAC Aligner (version 03.16.02.19) and small variants were called using Starling Small Variant Caller (version 2.4.7). Samples were aligned to the Homo Sapiens NCBI GRCh38 assembly with decoys.

Aggregation of single-sample gVCFs was performed using the Illumina software gVCF genotyper (version 2019). Variant normalisation and decomposition were implemented by vt (version 0.57721). Genomic annotation and calculation of allele statistics were performed using Ensembl VEP and bcftools respectively. The multi-sample VCF dataset (aggV2) was then split into 1,371 roughly equal chunks to allow faster processing. Only variants that passed all provided site quality control criteria were processed.

***Imputation***

The WGS genotypes (~722M variants) were filtered to a variant base list used for PGS model generation, which includes 18,421,839 variants. (For further information on how the variant list was derived see: https://research-help.genomicsengland.co.uk/pages/viewpage.action?pageId=72351761)

Genotypes were phased and imputed using the 1000G reference panel (v5a) which was lifted-over from GRCh37 to GRCh38 using cross-map.

***Ancestry assignment***

The genetic ancestry of the patients was estimated using a random forest classifier and data from 1000 genomes project phase 3 (1KGP3) dataset. Firstly, all unrelated samples from the 1KGP3 were selected and 188,382 HQ SNPs were subsetted. After filtering for MAF > 0.05 in 1KGP3 (and GE data), the first 20 PCs were calculated using GCTA and the aggV2 data were projected onto the 1KGP3 PC loadings. The random forest model to predict ancestries was trained based on:

a. First 8 1KGP3 PCs
a. set Ntrees = 400
a. Train and predict on 1KGP3 Admixed American, African, East Asian, European, and South Asian super-populations.

Individuals were assigned for any one ancestry with a probability of > 0.8.

**Generation Scotland**

***Registry data***

Disease outcomes were ascertained through linkage to primary (GP) and secondary (hospital) healthcare records. Individuals were subsetted to those registered at a GP that consented to sharing of primary records. GP records consisted of Read2 codes, which were mapped to ICD-10. Hospital data were obtained from Scottish Morbidity Records (SMR) where disease outcomes were coded using ICD-9 (pre March 1997) or ICD-10 (post March 1997).

***Genotyping and quality control***

Generation Scotland (GS) consists of ~24,000 individuals from across Scotland aged between 18-99 years. Phenotypic data were obtained at baseline along with whole blood samples for DNA quantification.

Genotype data was assayed for 20,195 participants in two batches with 9,863 participants in the first batch and the remainder in the second. The genotyping was performed using the Illumina HumanOmniExpressExome-8 v1.0 BeadChip and the Illumina HumanOmniExpressExome-8 v1.2 BeadChip, respectively. Individuals or SNPs with a low call rate (<98%) and SNPs with Hardy-Weinberg p-value<1x10^-6^ were removed. Mendelian errors were removed by setting the individual-level genotypes at erroneous SNPs to missing.

***Imputation***

Genotyped data were imputed using the HRC panel v1.1 (4). Autosomal haplotypes were checked to ensure consistency with the reference panel (strand orientation, reference allele, position. Pre-phasing was performed using Shapeit2 v2r837 (19,20) using the Shapeit2 duohmm option11 (21) and cohort family structure in order to improve imputation quality (22). Variants with low imputation quality (INFO<0.4) as well as monogenic variants were removed from the imputed set resulting in 24,111,857 variants for downstream analysis.

***Ancestry assignment***

Ancestry outliers were removed from the dataset. These were defined as individuals who were more than six standard deviations away from the mean in a principal component analysis of GS merged with 1092 participants from the 1000 Genomes Project (6).

**Comparing two Hazard Ratios**

To determine whether the hazard ratios (HRs) differ by sex, we compared the HRs directly as well as more formally in an interaction. When comparing the HRs directly we employed the following method:

Firstly the difference between the log(HRs) for men and women is calculated:

Delta = log(HR_men_) - log(HR_women_)

The standard error of the difference is calculated as:

Standard error of difference = (SE_men_)^2^ + (SE_women_)^2^

Finally, a Z-score is created to derive a p-value:

Z-score = Delta / Standard error of difference

### Supplementary Figures

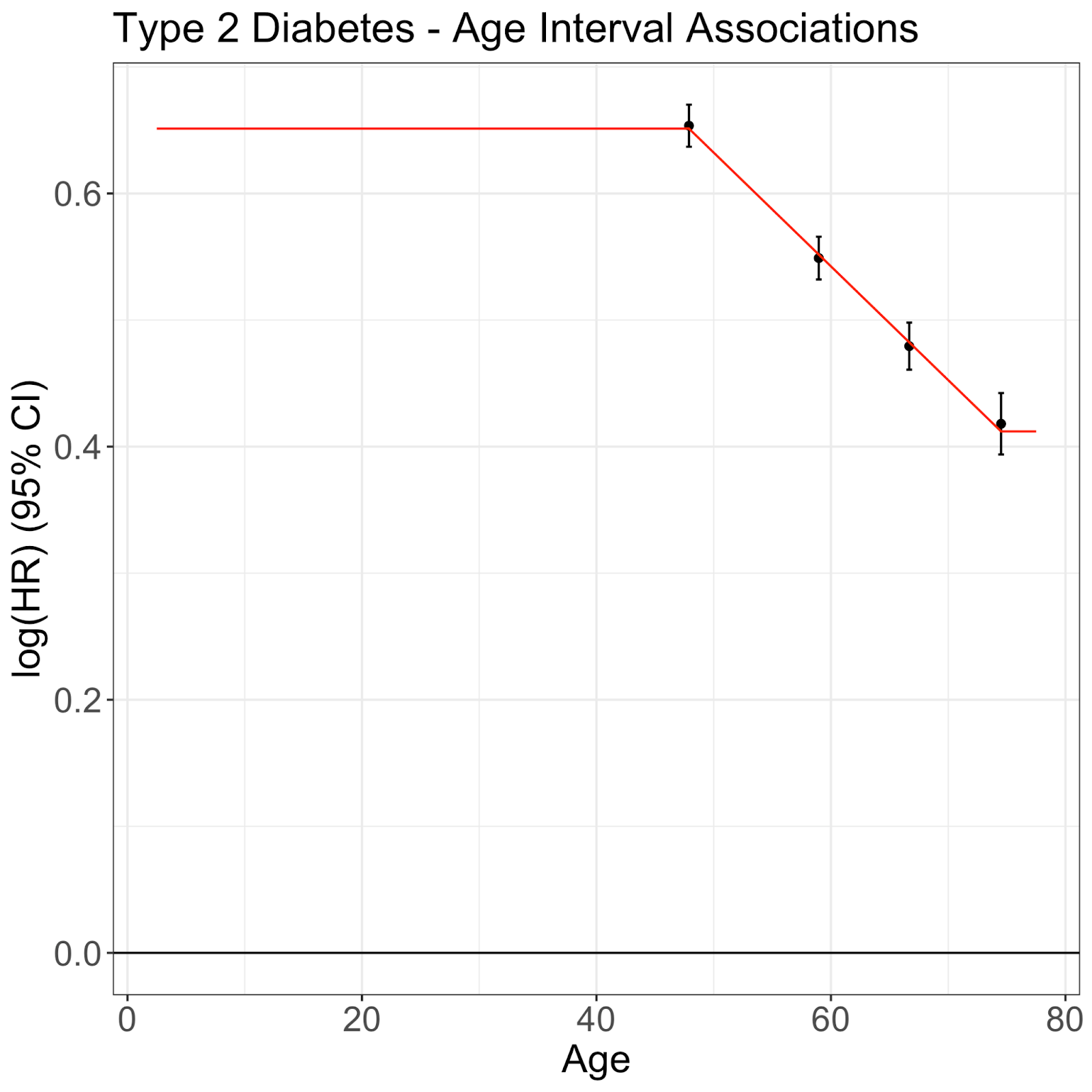

**Supplementary Figure 1.** Estimating age-specific hazard ratios. This schematic describes how age-specific hazard ratios were estimated. Using Type 2 Diabetes and FinnGen data as an example, Cox Proportional Hazard models were first performed on four intervals. The log hazard ratios plotted above are placed at the median age at onset within each interval and a weighted linear regression was fit to the data. The predicted age specific log hazard ratios from the weighted linear regression are plotted in red. For any age outside of the range, the log hazard ratio is assumed to be equal to the nearest age in which a prediction was in range.

**a)**

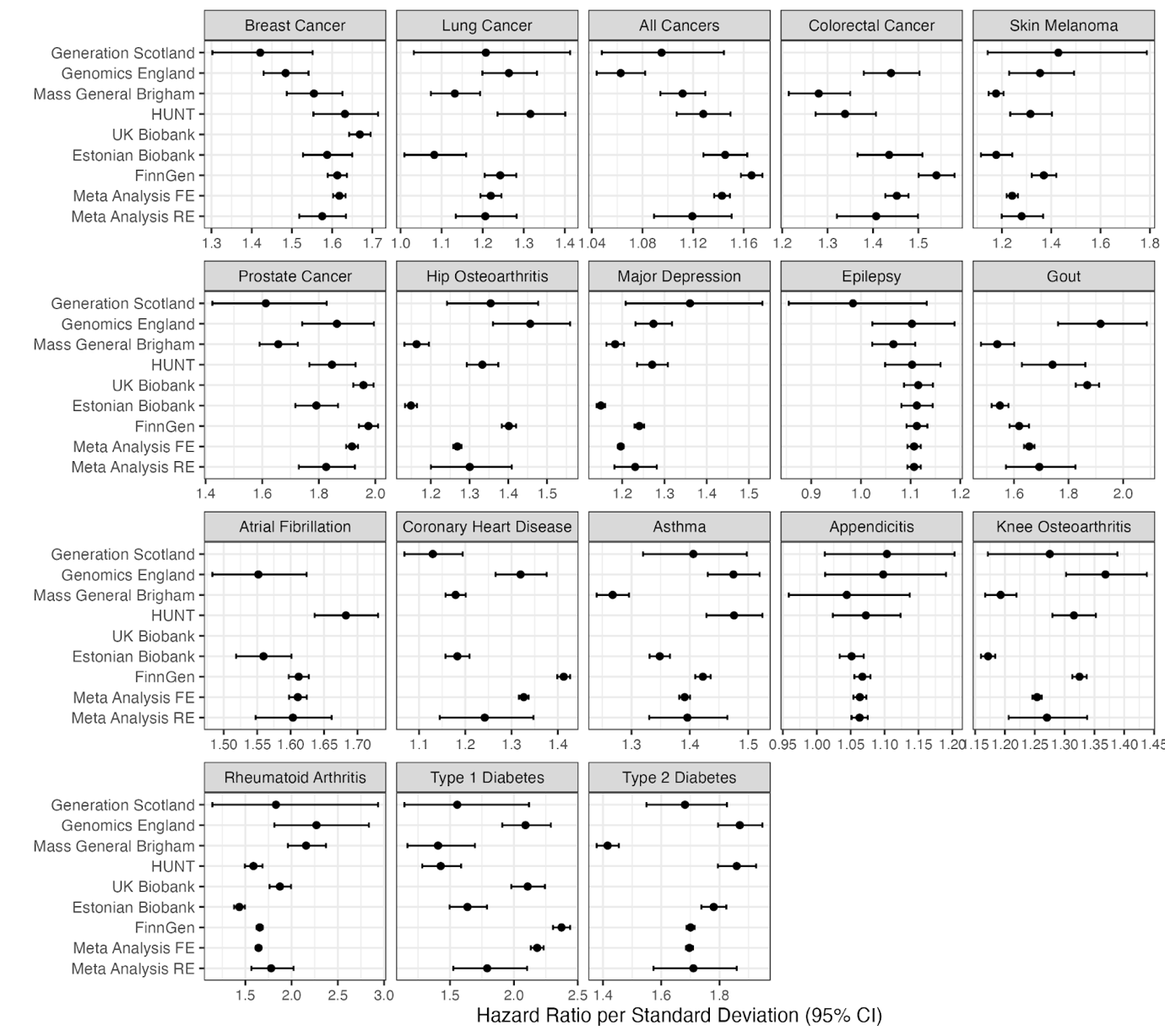

**b)**

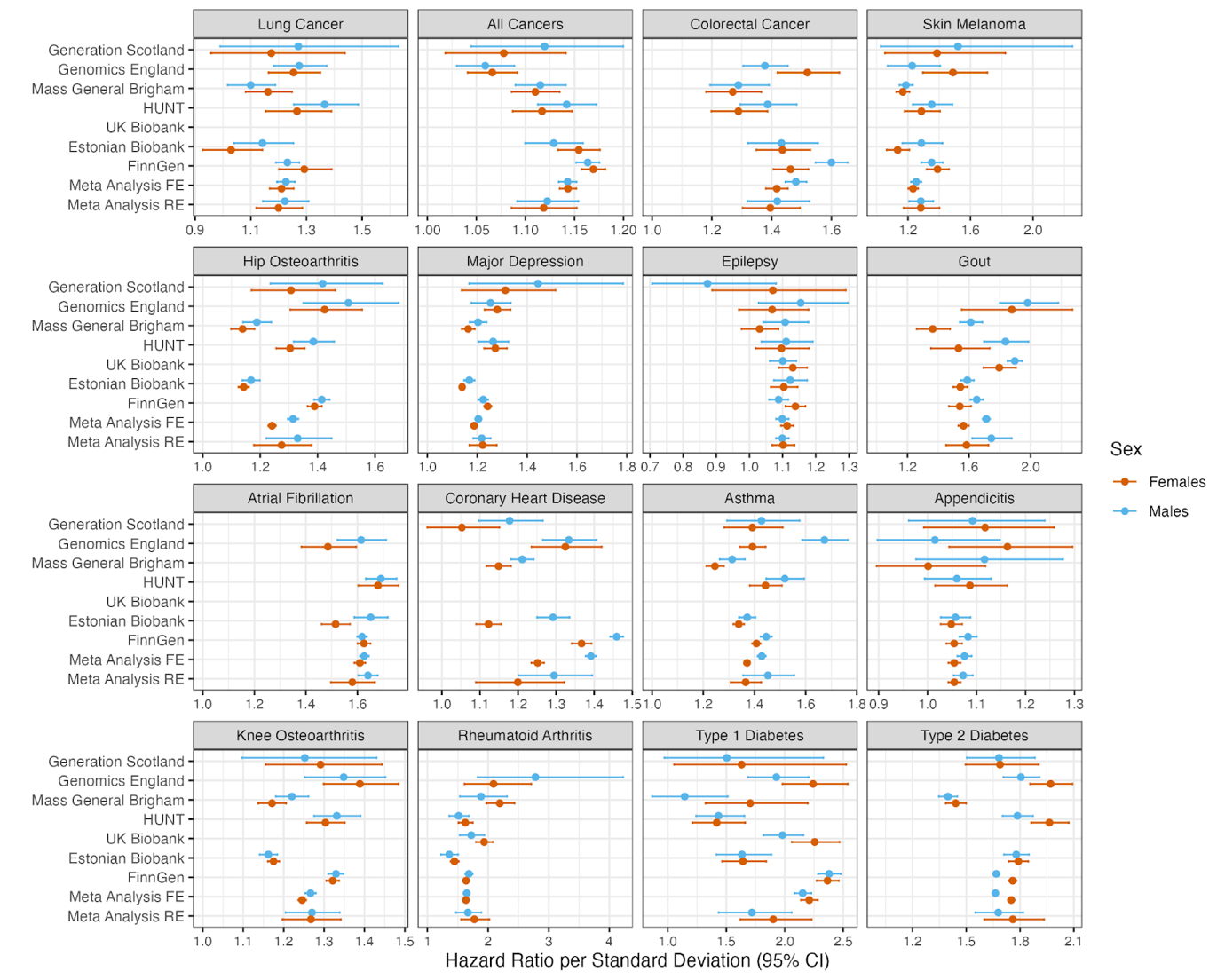

**c)**

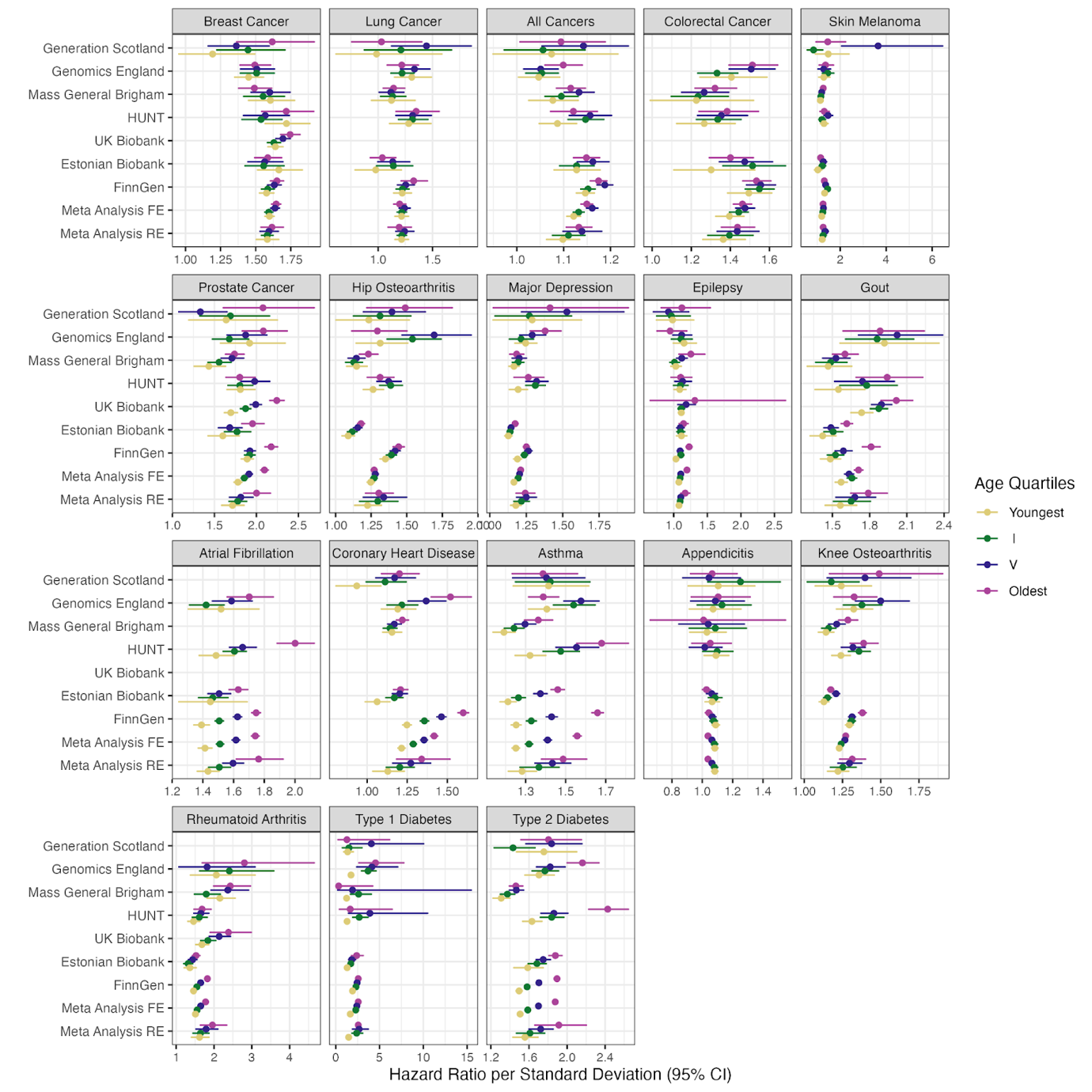

**d)**

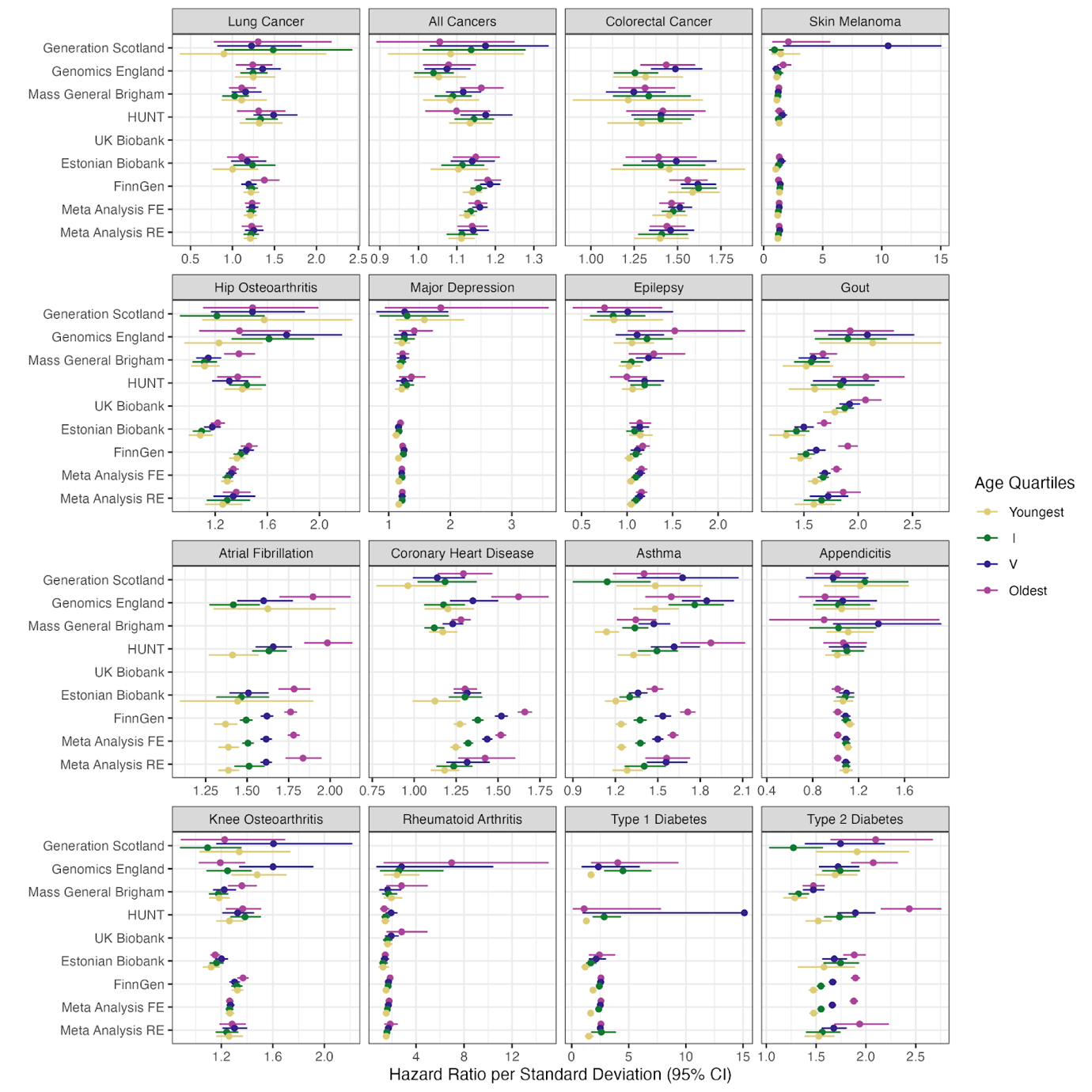

**e)**

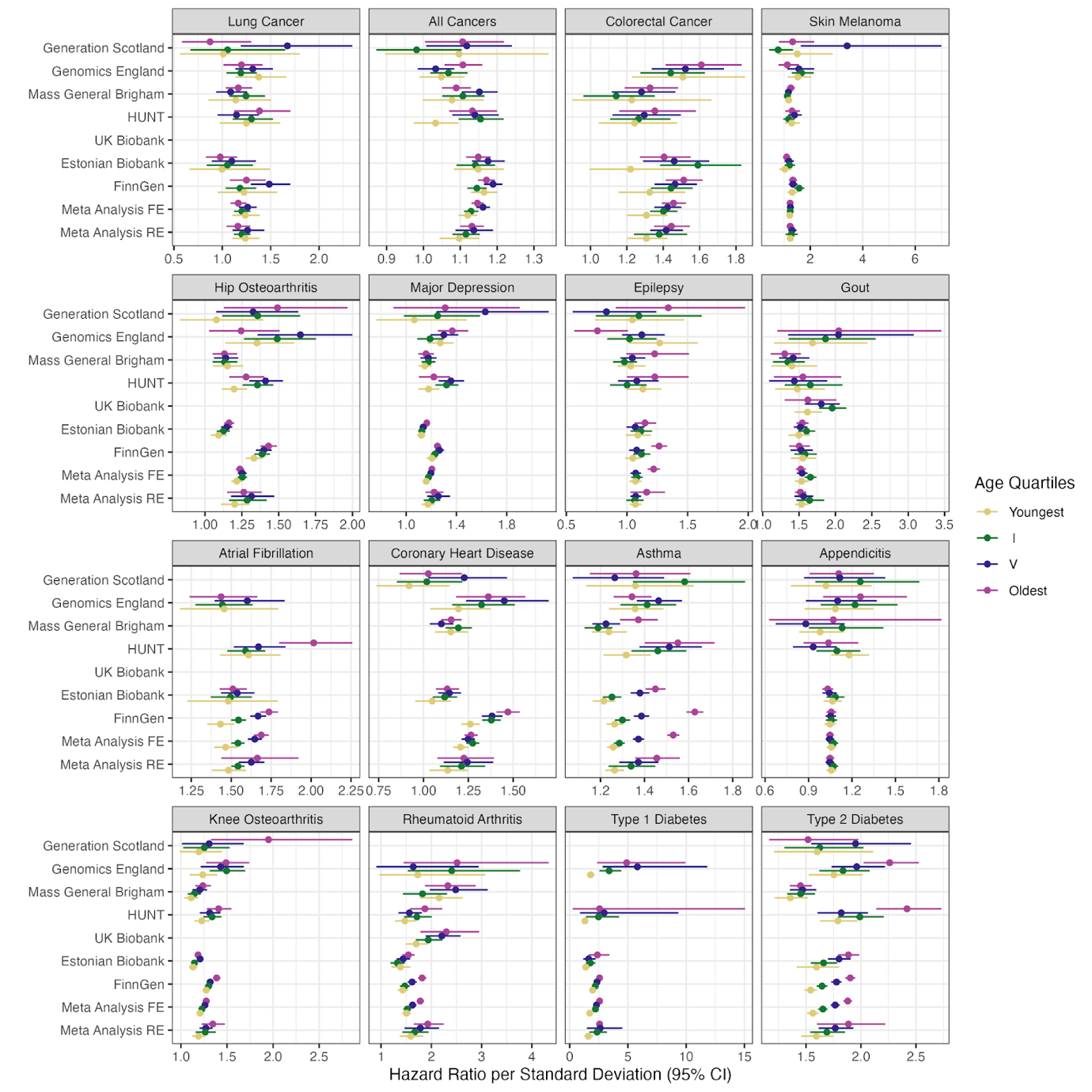

**Supplementary Figure 2.** Forest plots of each phenotype. a) No stratification. b) Sex stratification. c) Age stratification. d) Age and sex stratification - Males. e) Age and sex stratification - Females. Note: for figures d and e the limit has been restricted to 15 for presentability.

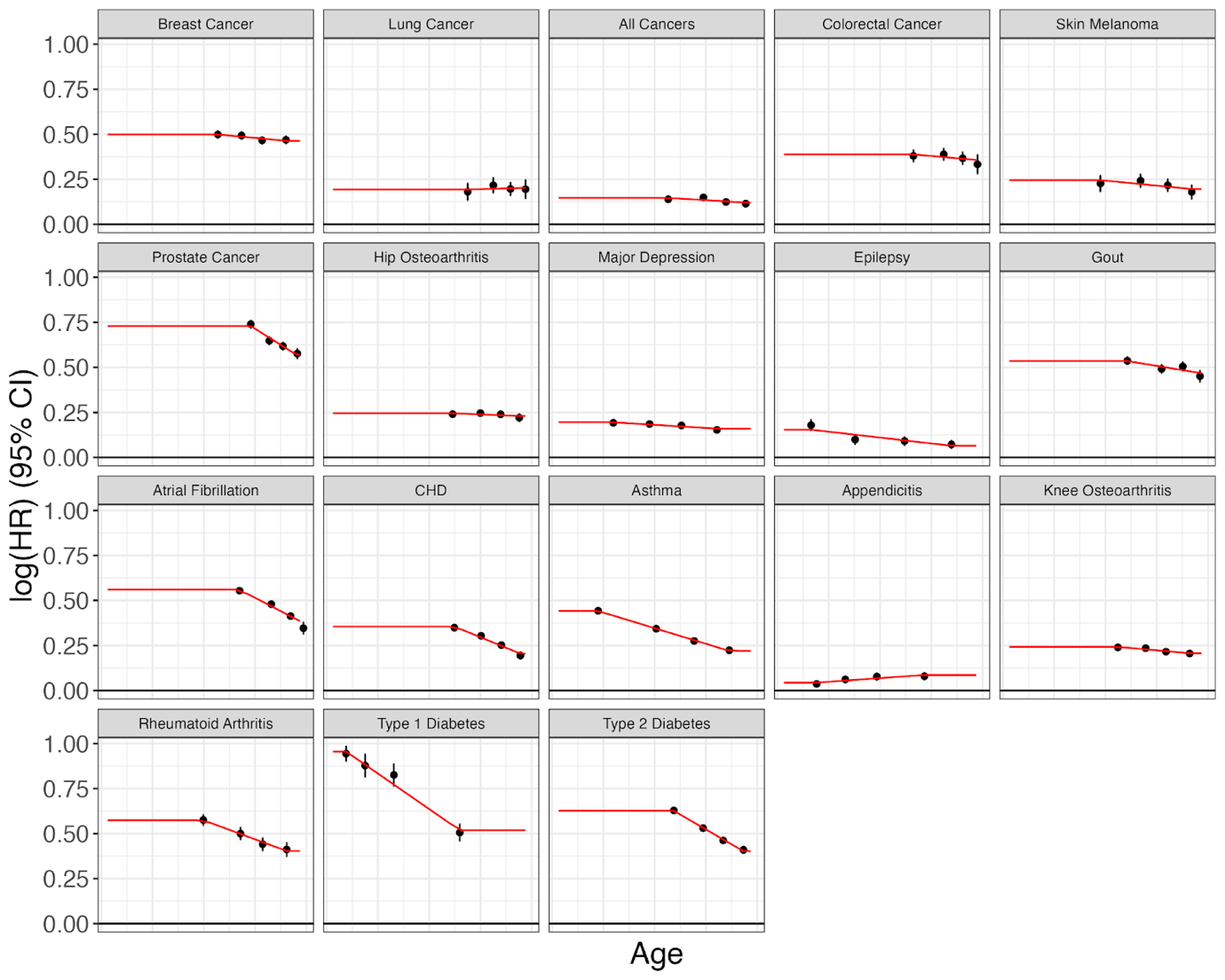

**Supplementary Figure 3.** Age specific effects of meta-analyzed log(Hazard Ratios)

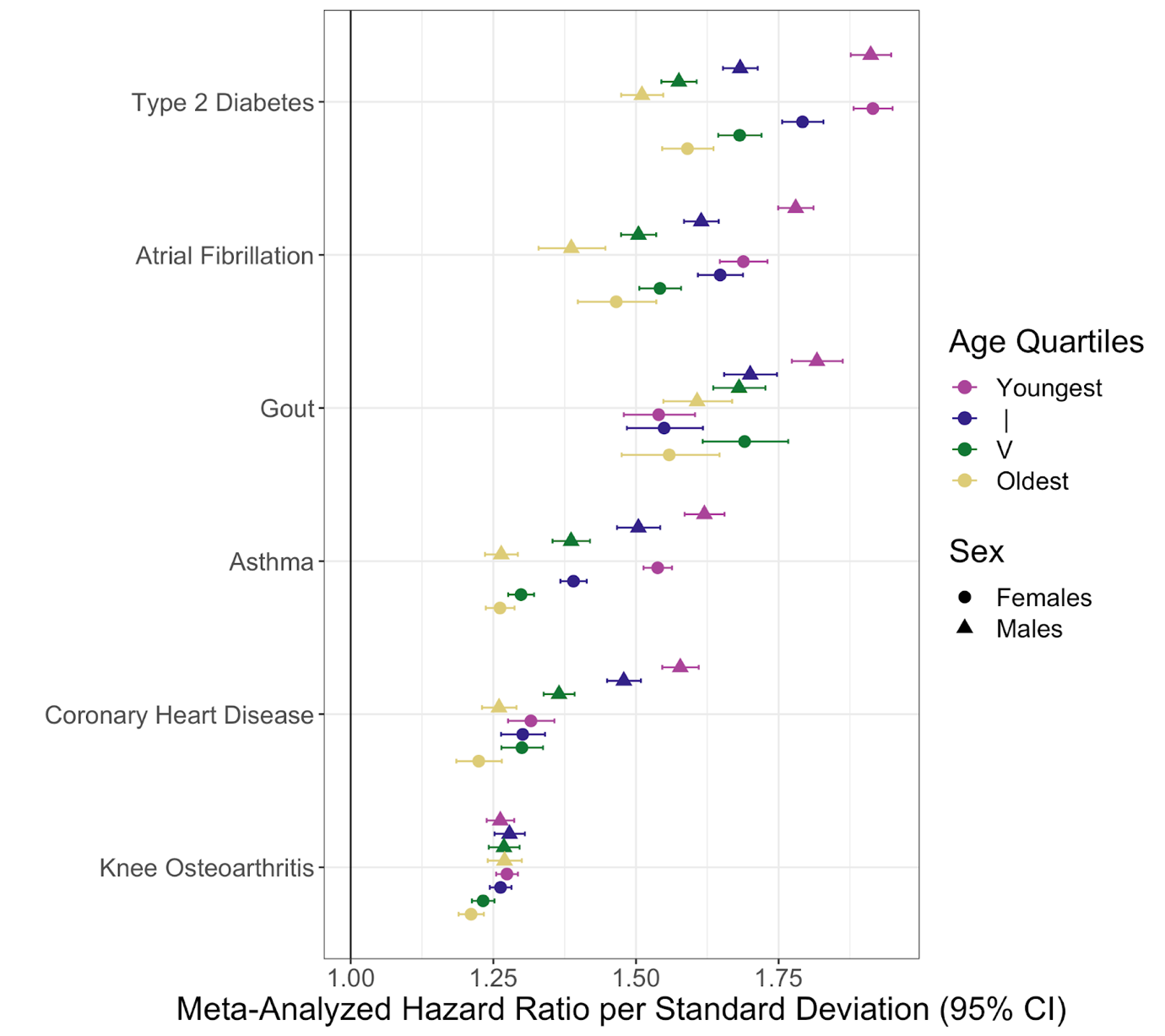

**Supplementary Figure 4.** Hazard ratios per standard deviation stratified by age and sex. Note: phenotypes were only included if the model selected was age and sex stratified.

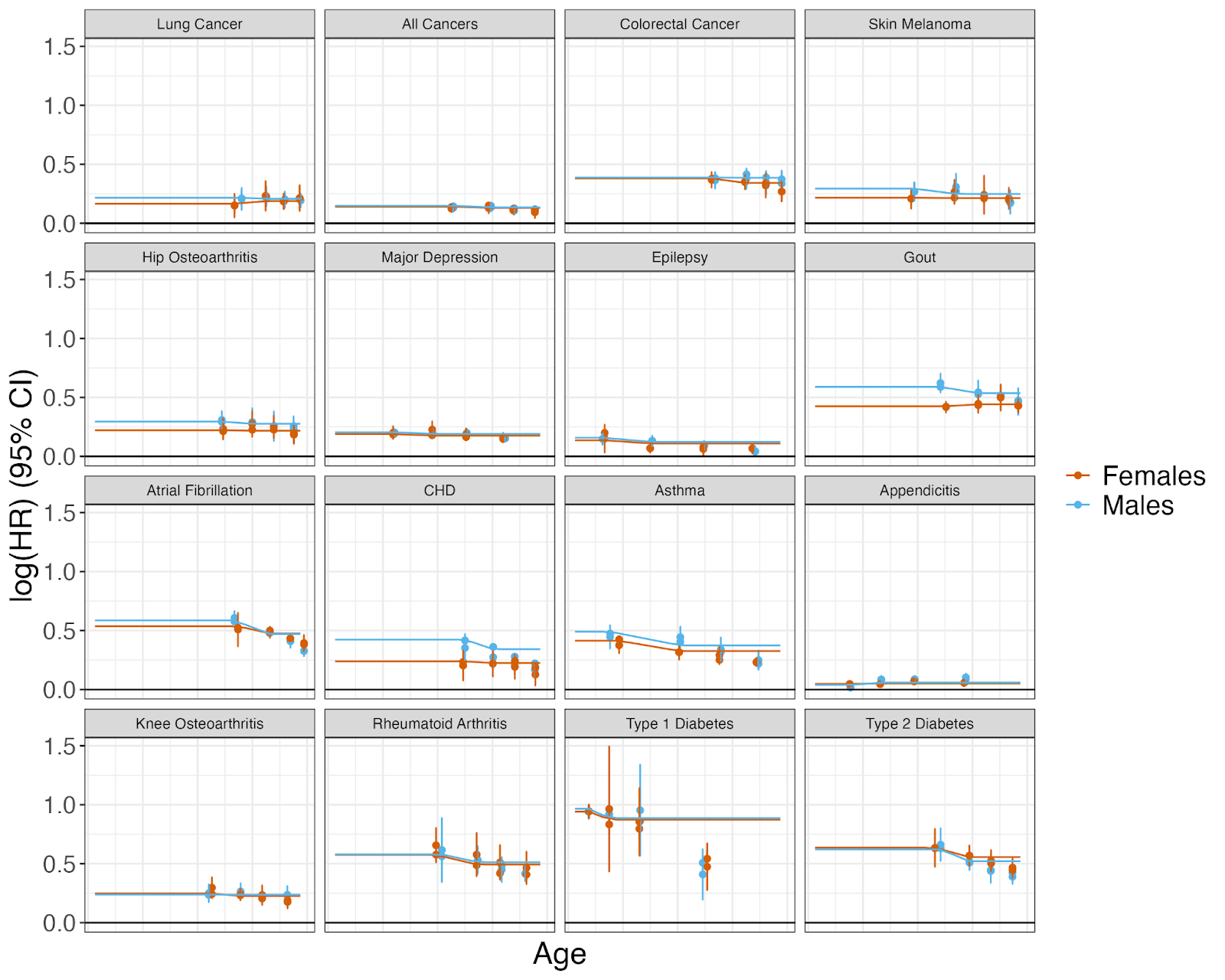

**Supplementary Figure 5.** Age specific effects of meta-analyzed log(Hazard Ratios) stratified by sex

**a. All Cancers**

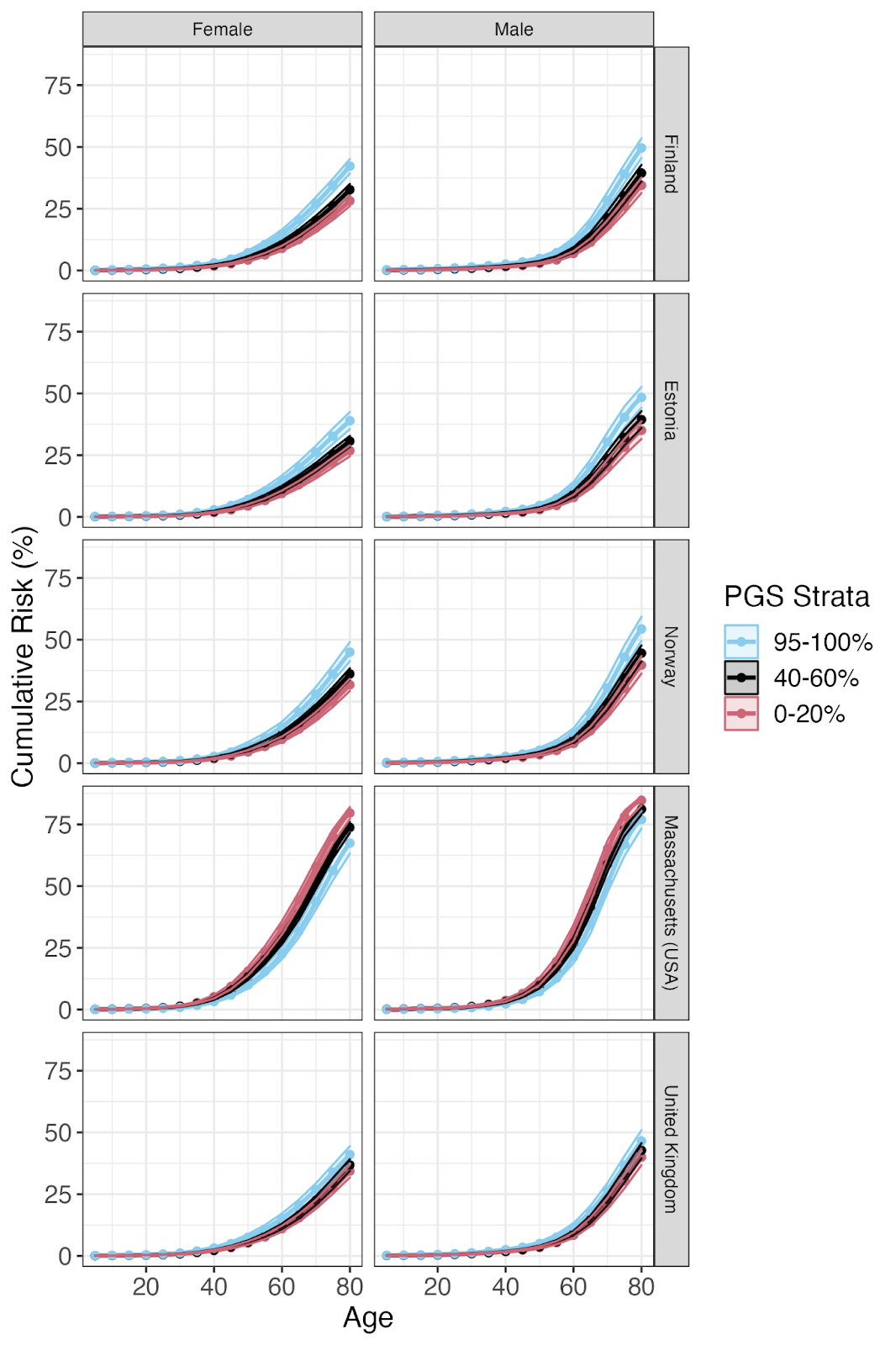

**b.** **Appendicitis**

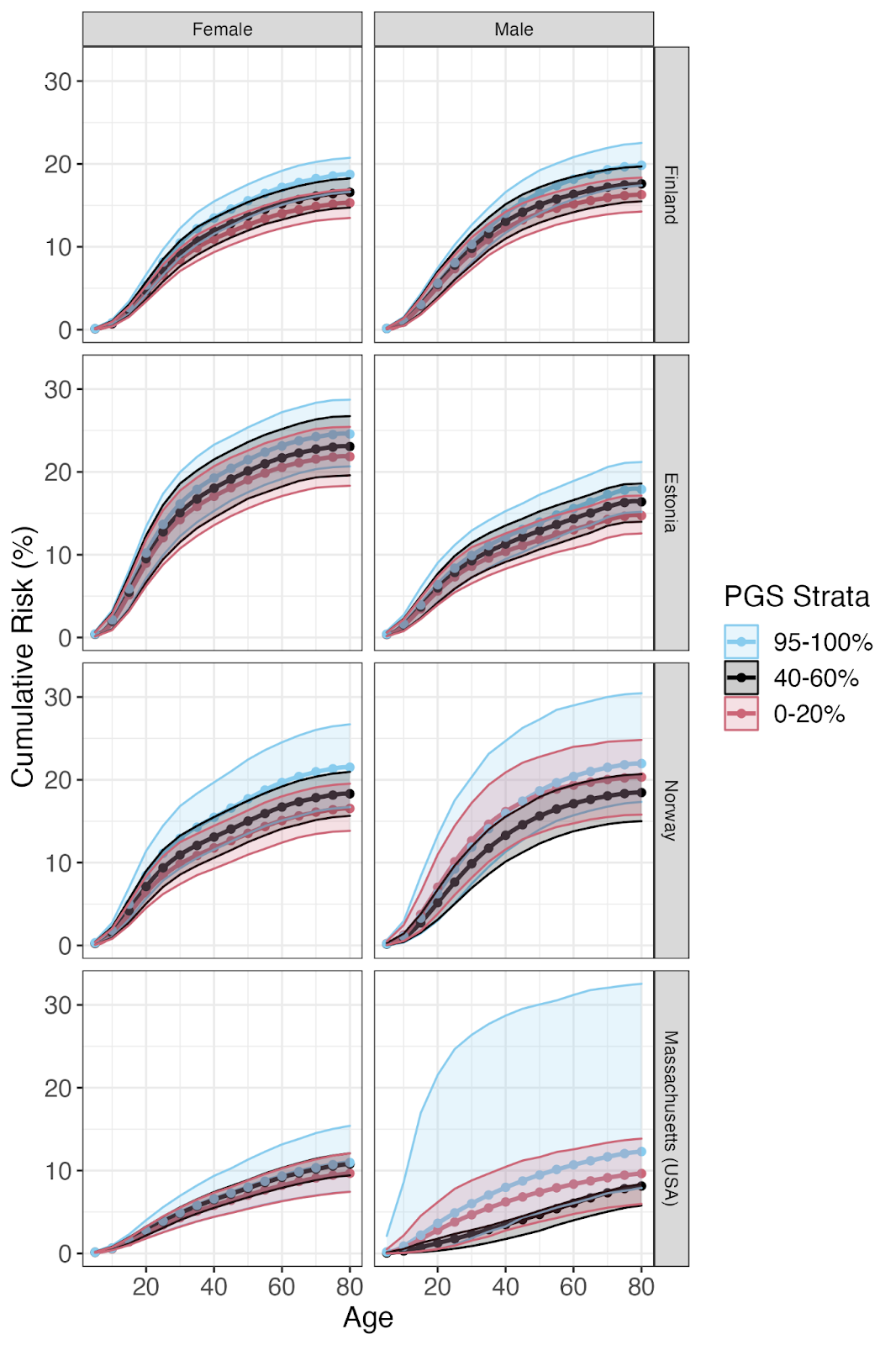

c. **Asthma**

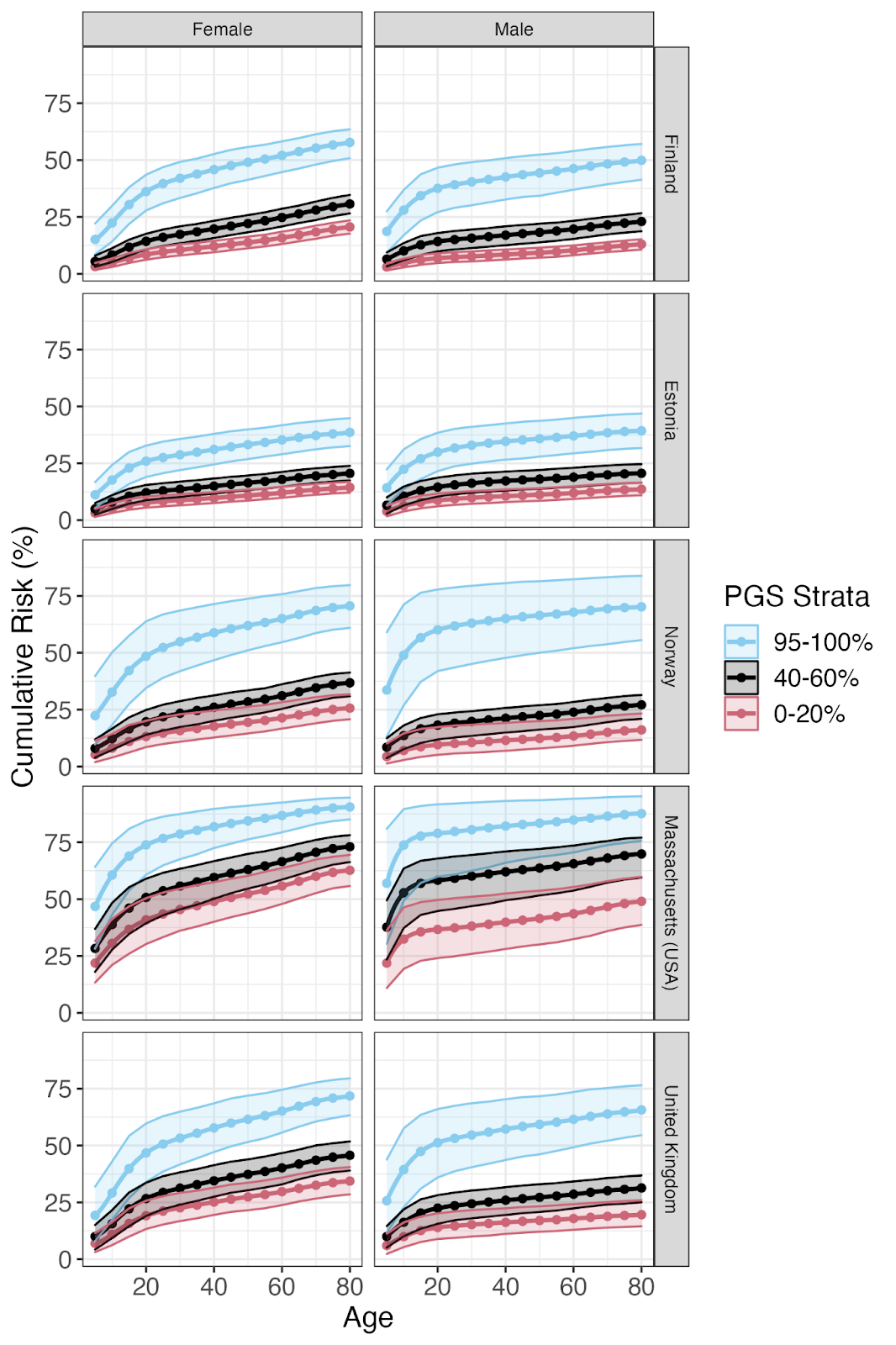

**d. Atrial Fibrillation**

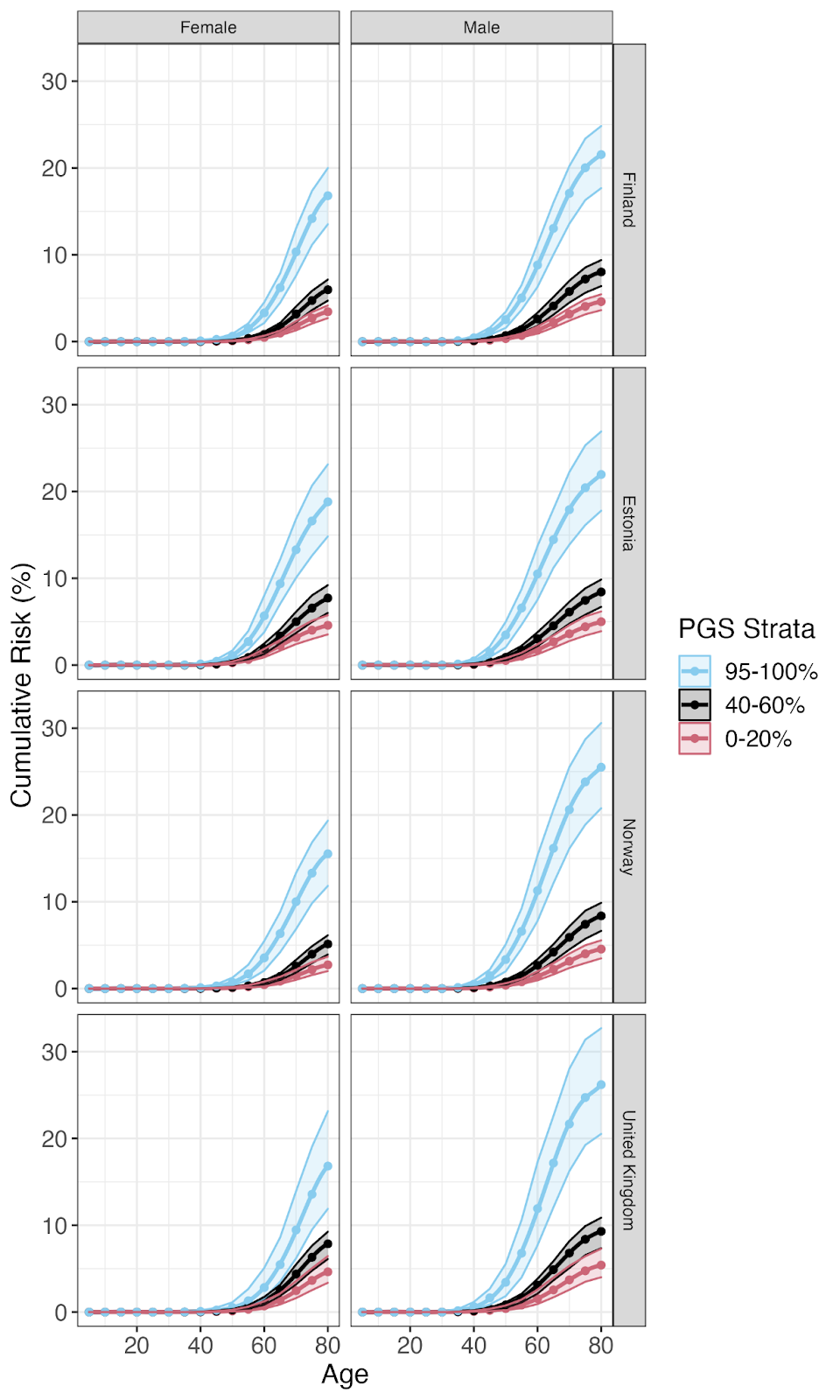

e. **Breast Cancer**

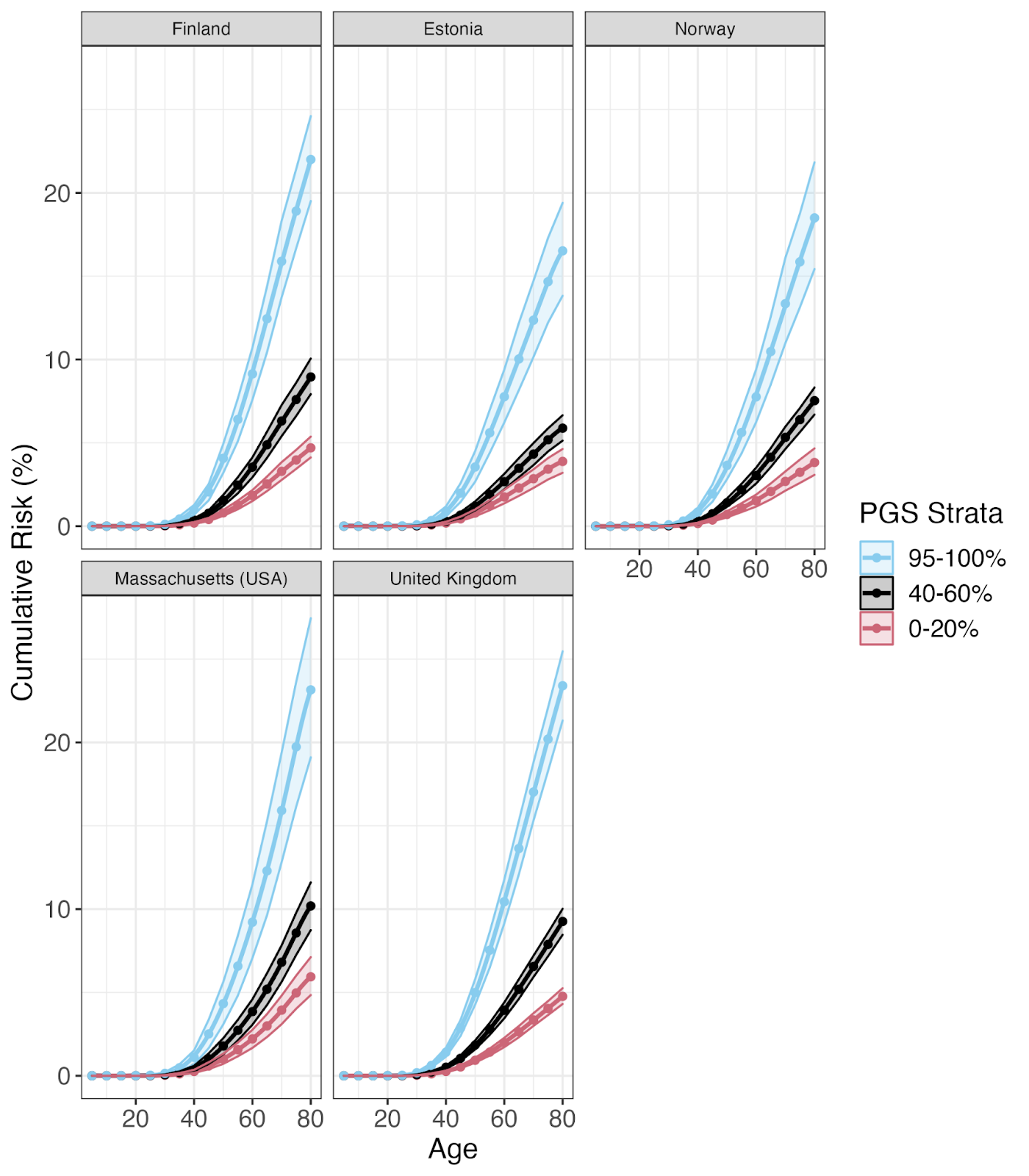

f. **Colorectal Cancer**

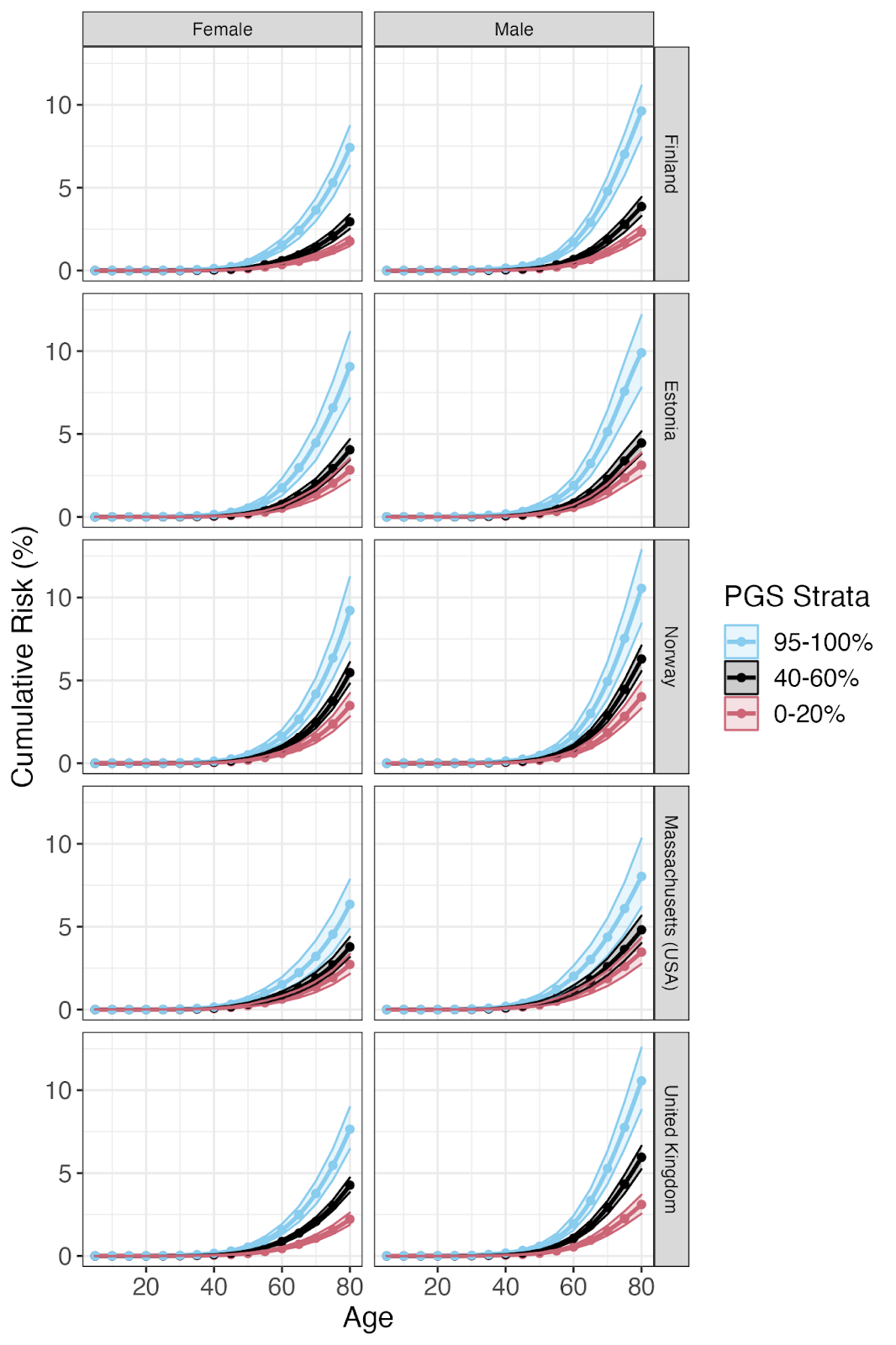

g. **Epilepsy**

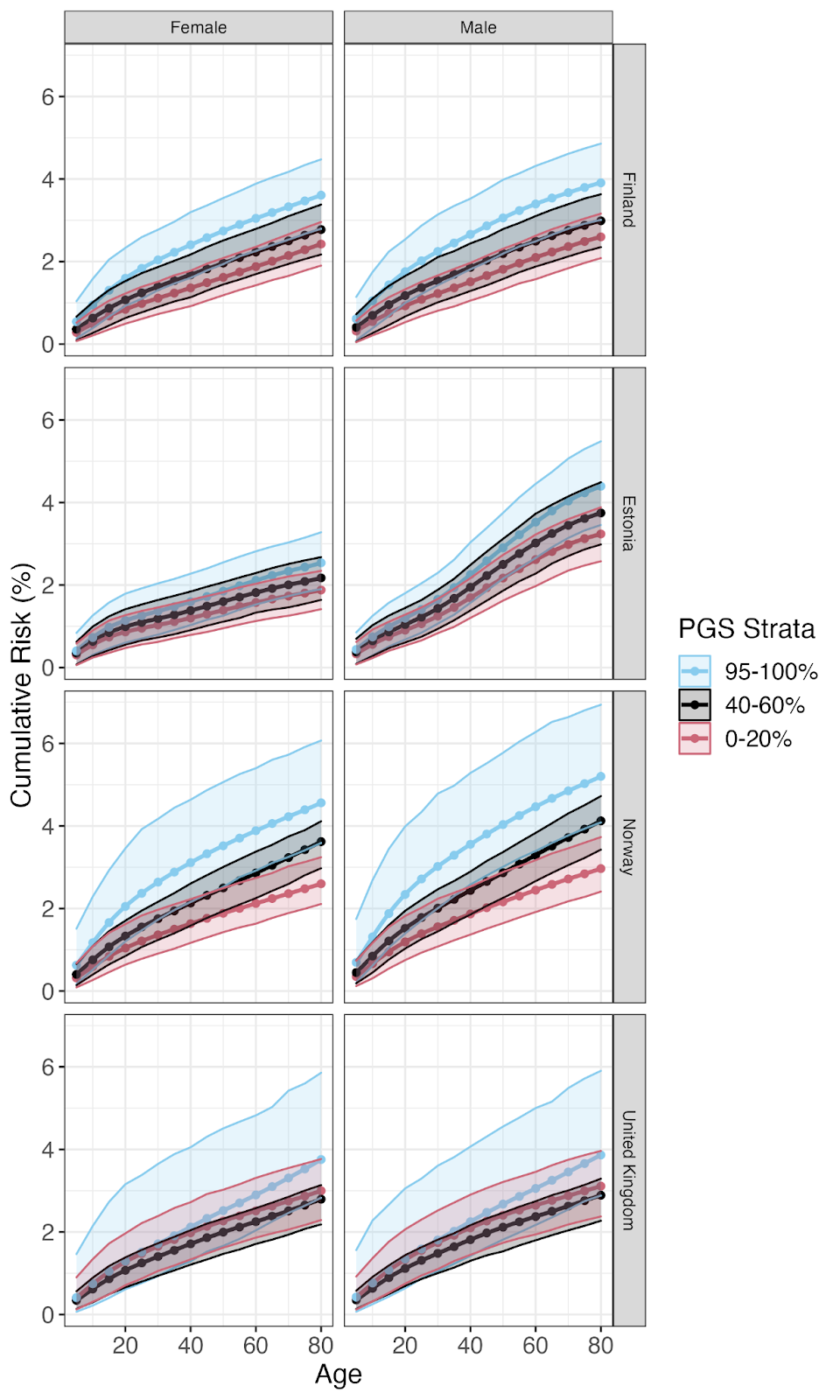

h. **Gout**

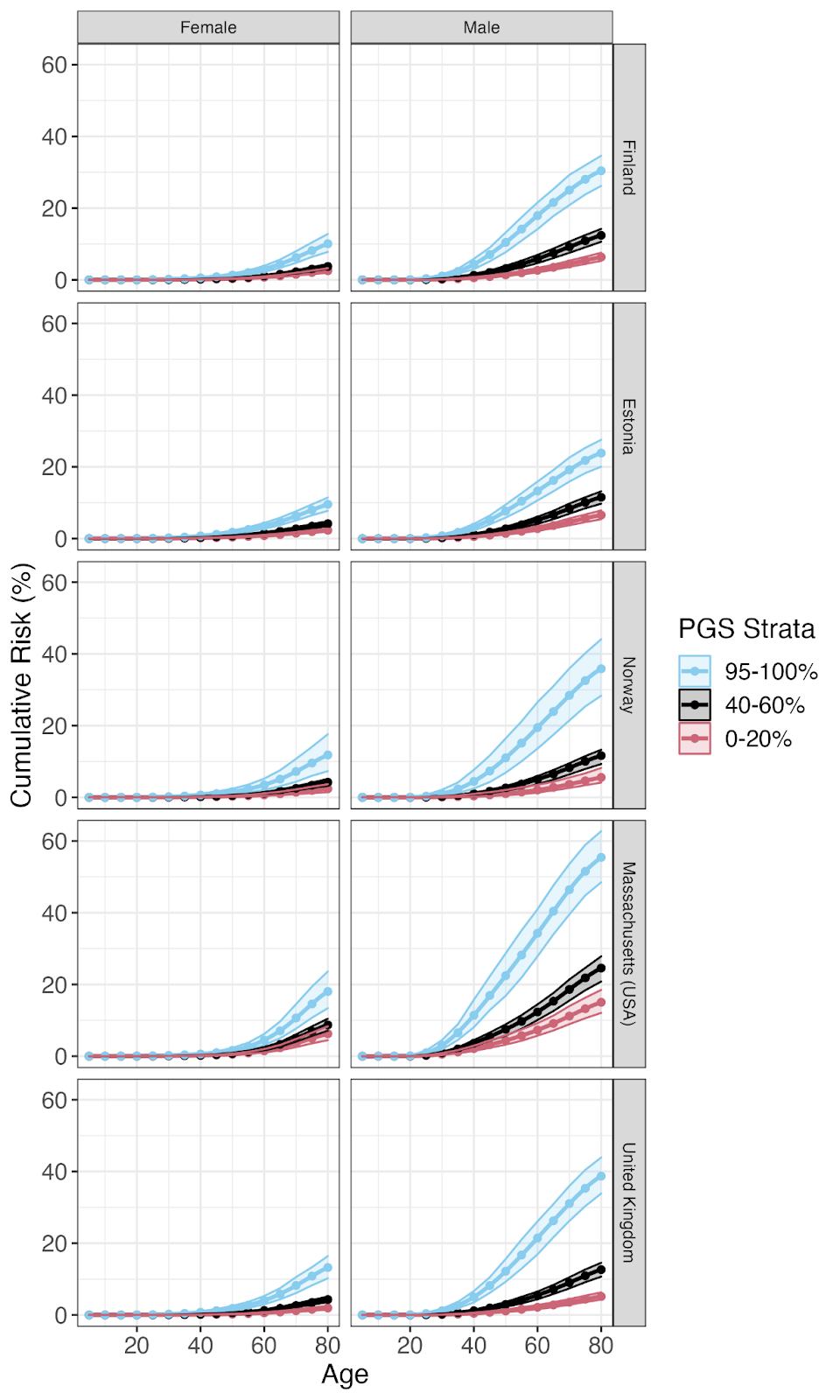

i. **Hip Osteoarthritis**

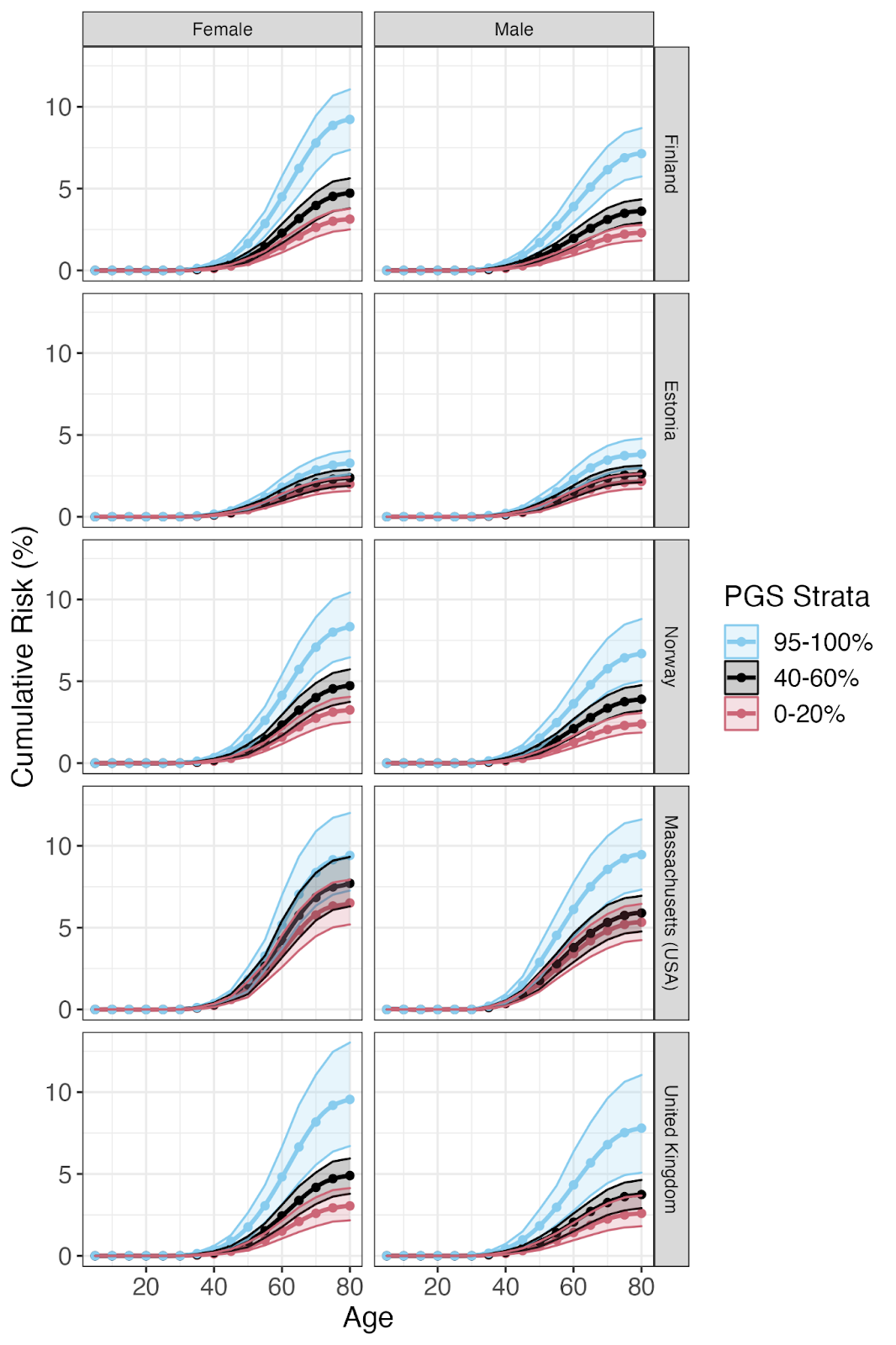

j. **Knee Osteoarthritis**

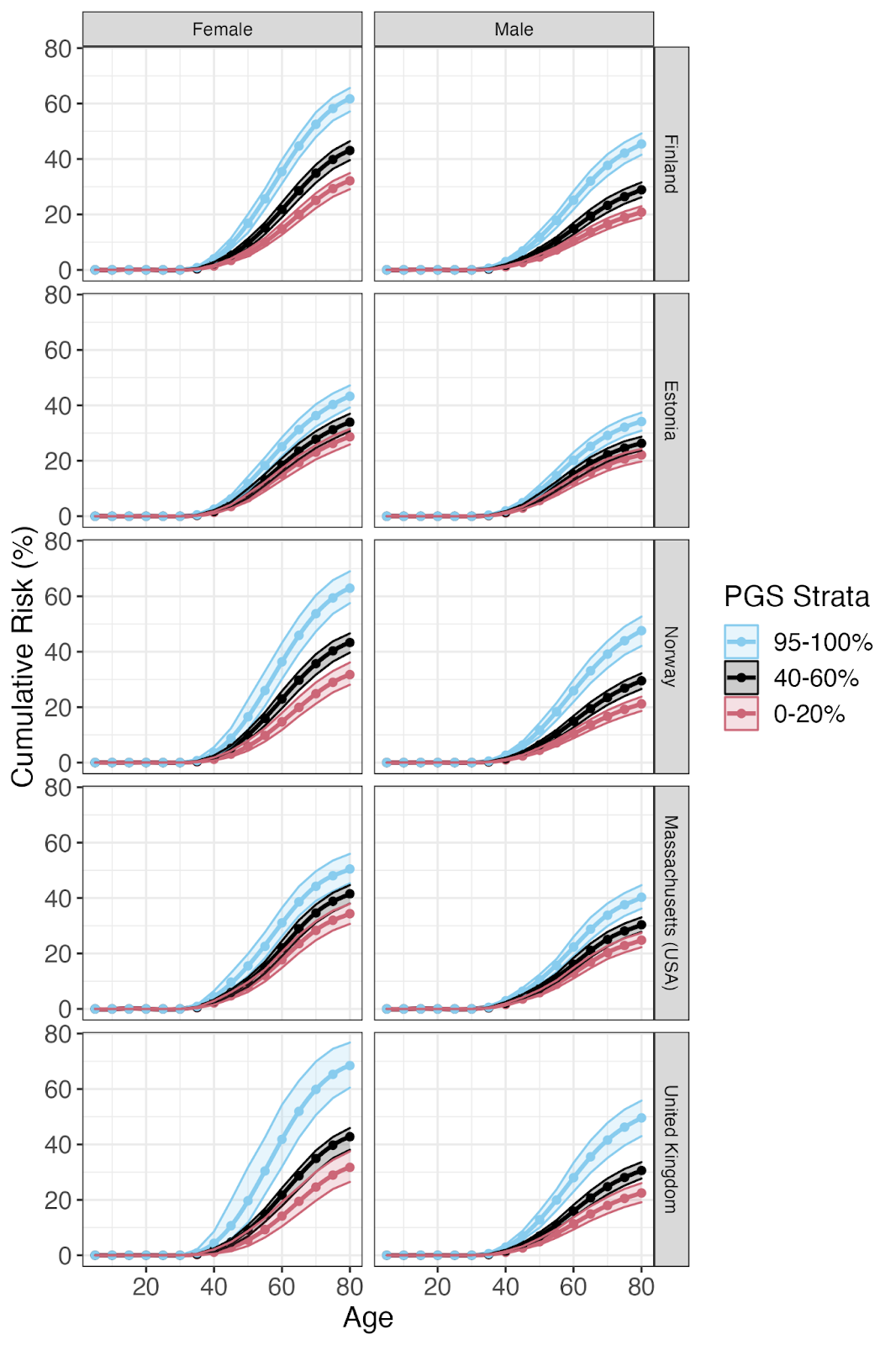

k. **Lung Cancer**

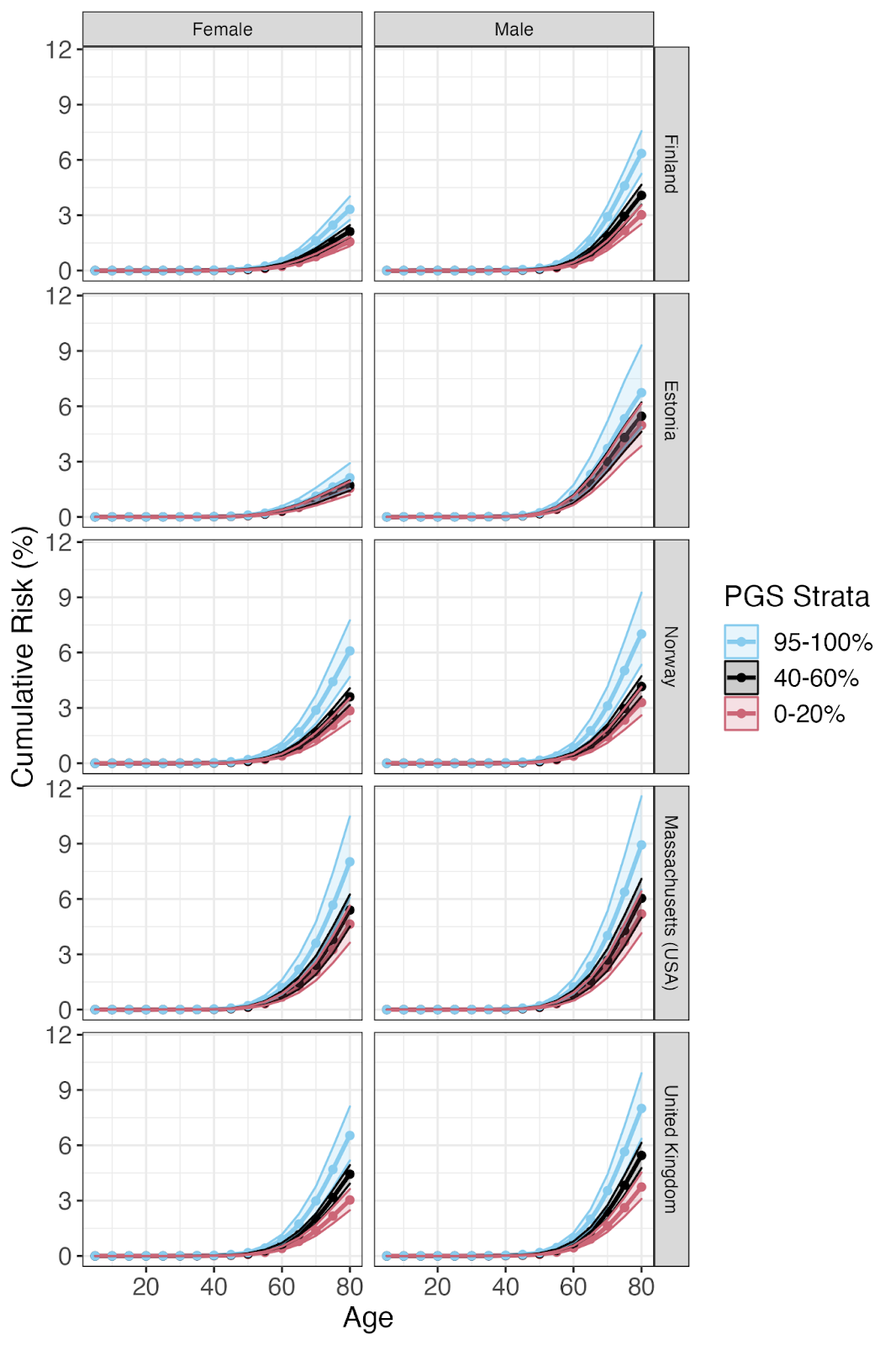

l. **Depression**

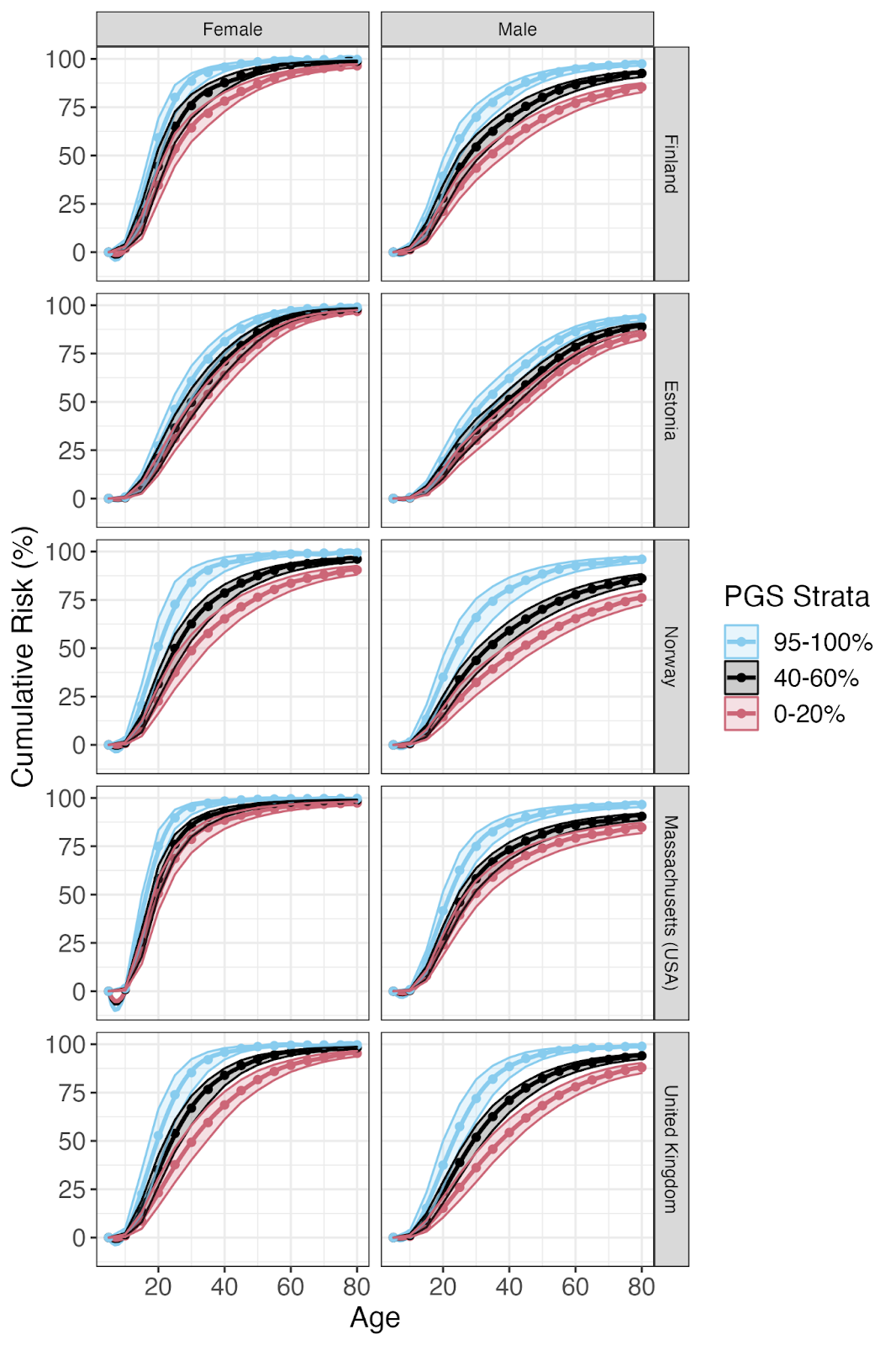

m. **Skin Melanoma**

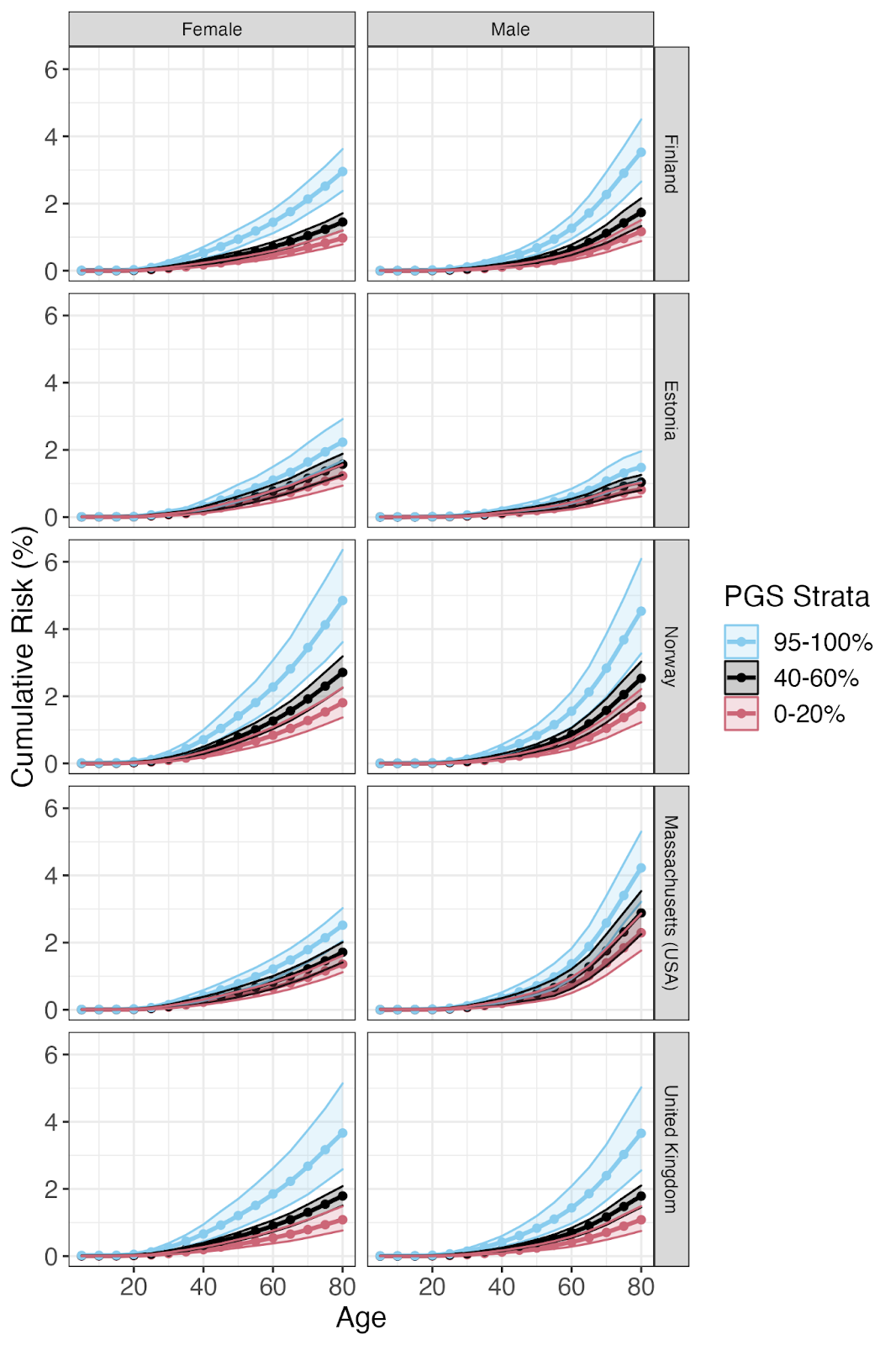

n. **Rheumatoid Arthritis**

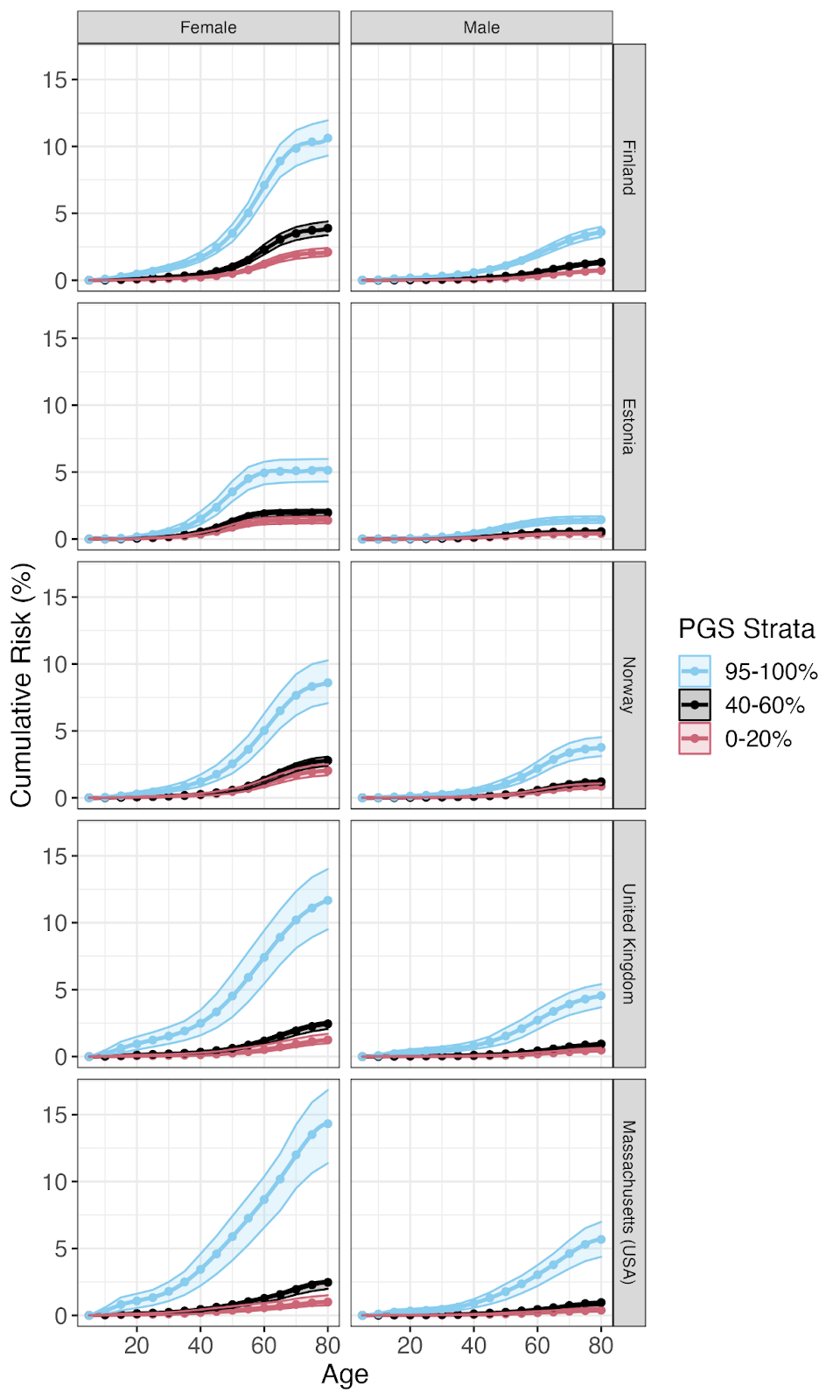

o. **Type 1 Diabetes**

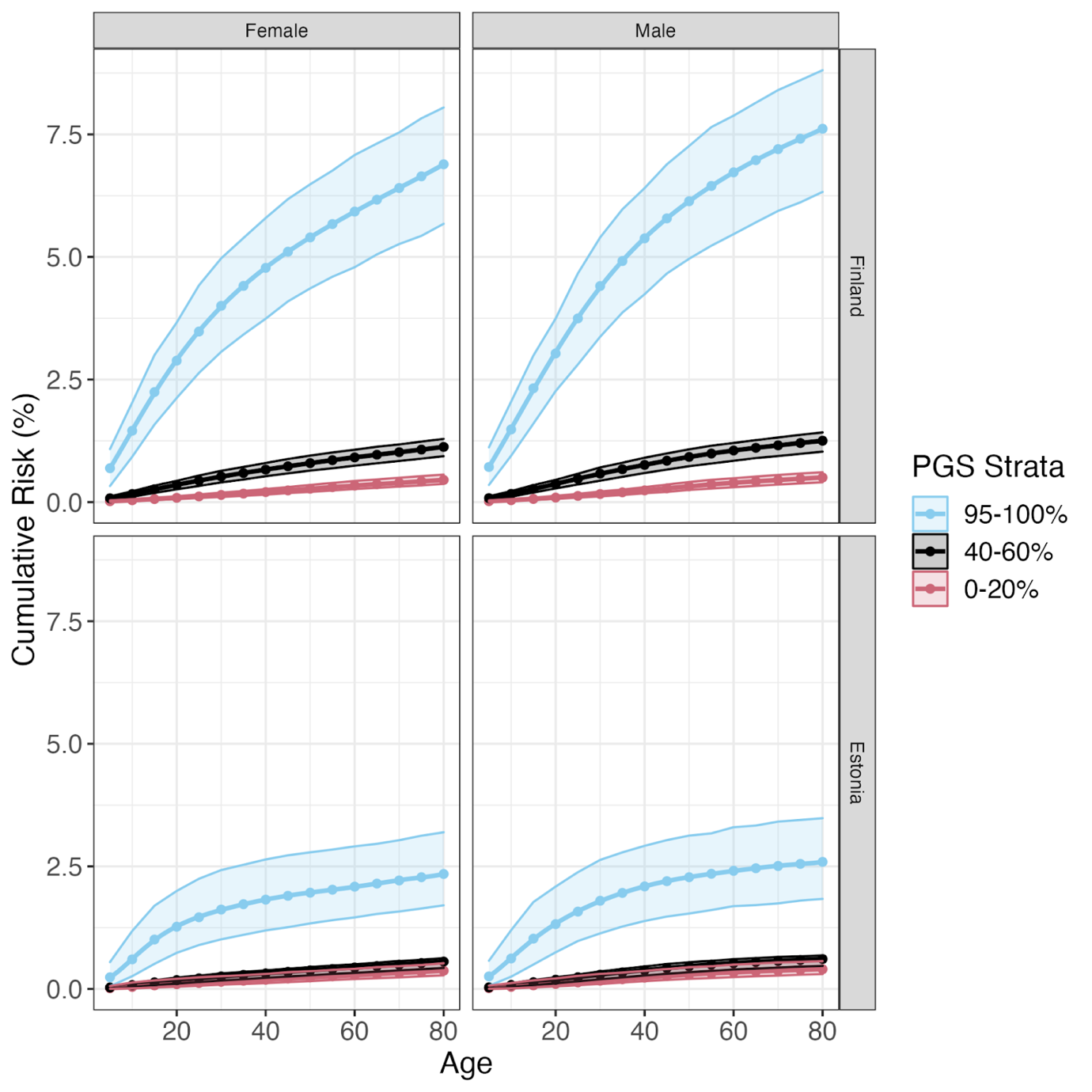

p. **Type 2 Diabetes**

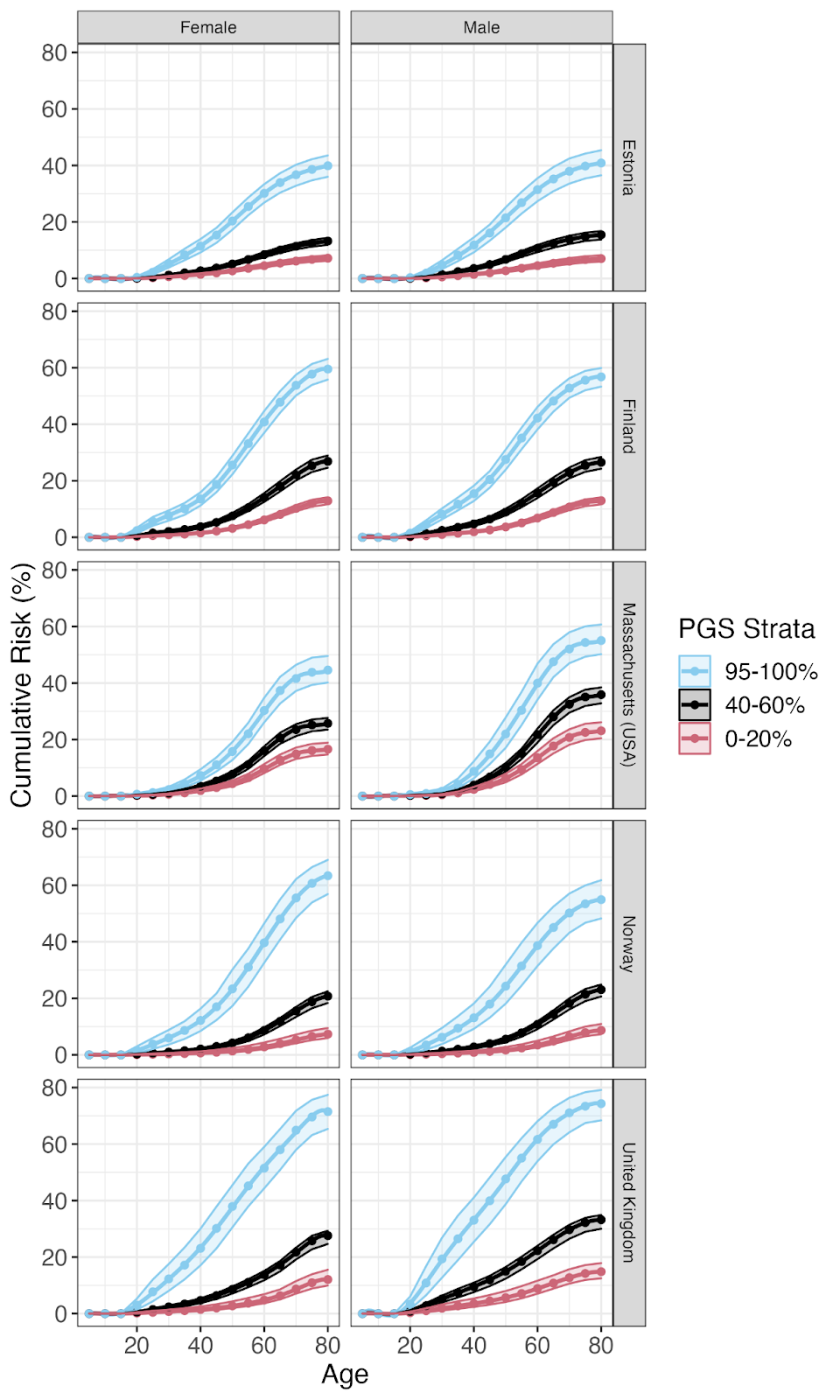

**Supplementary Figure 6.** Country and sex-specific cumulative absolute risk estimates in the top, bottom and reference percentile groupings also including uncertainty measures (95% confidence intervals). **a)** All Cancers. **b)** Appendicitis. **c)** Asthma. **d)** Atrial Fibrillation. **e)** Breast Cancer **f)** Colorectal Cancer. **g)** Epilepsy. **h)** Gout. **i)** Hip Osteoarthritis. **j)** Knee Osteoarthritis. **k)** Lung Cancer. **l)** Depression. **m)** Skin Melanoma. **n)** Rheumatoid Arthritis. **o)** Type 1 Diabetes. **p)** Type 2 Diabetes

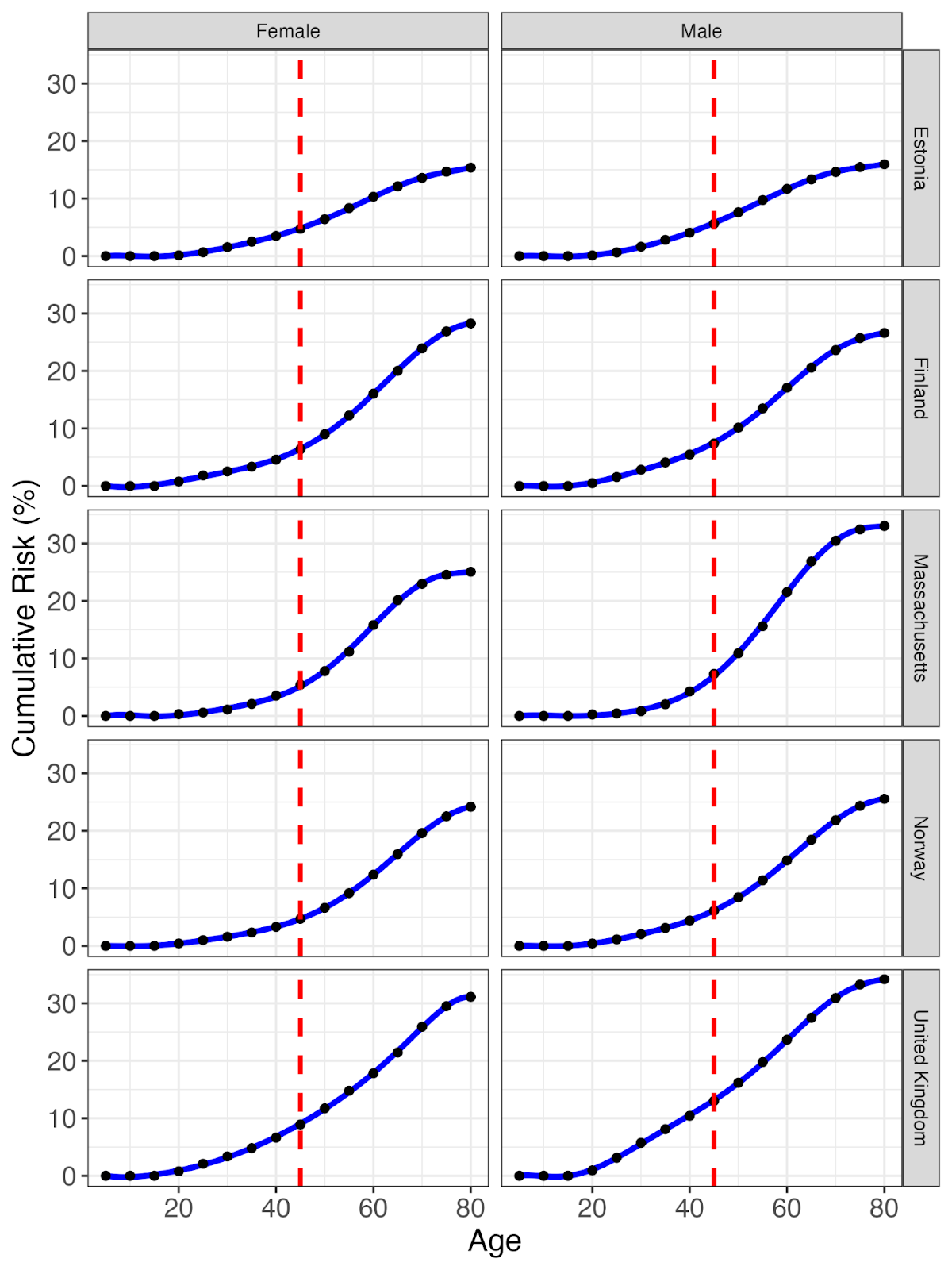

**Supplementary Figure 7.** Country-specific absolute risks for Type 2 Diabetes. Red dashed lines highlight age 45, the point at which the American Diabetes association recommends screening. The intersection with the blue cumulative risks were taken as the country's clinical threshold.

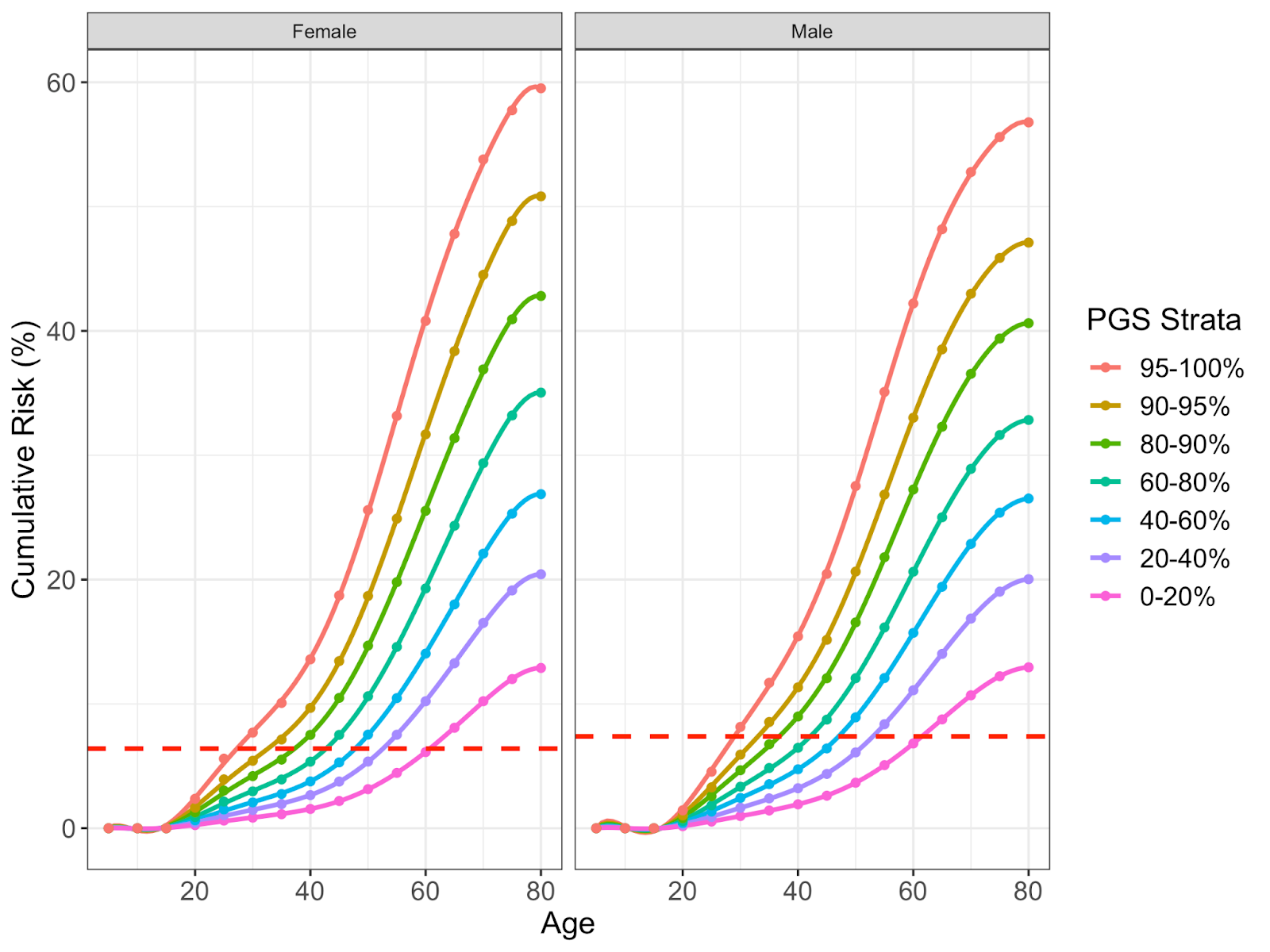

**Supplementary Figure 8.** Type 2 Diabetes cumulative incidence by PGS strata in the top and bottom percentile of the distribution and the reference category (40-60%) inclusive of confidence intervals.

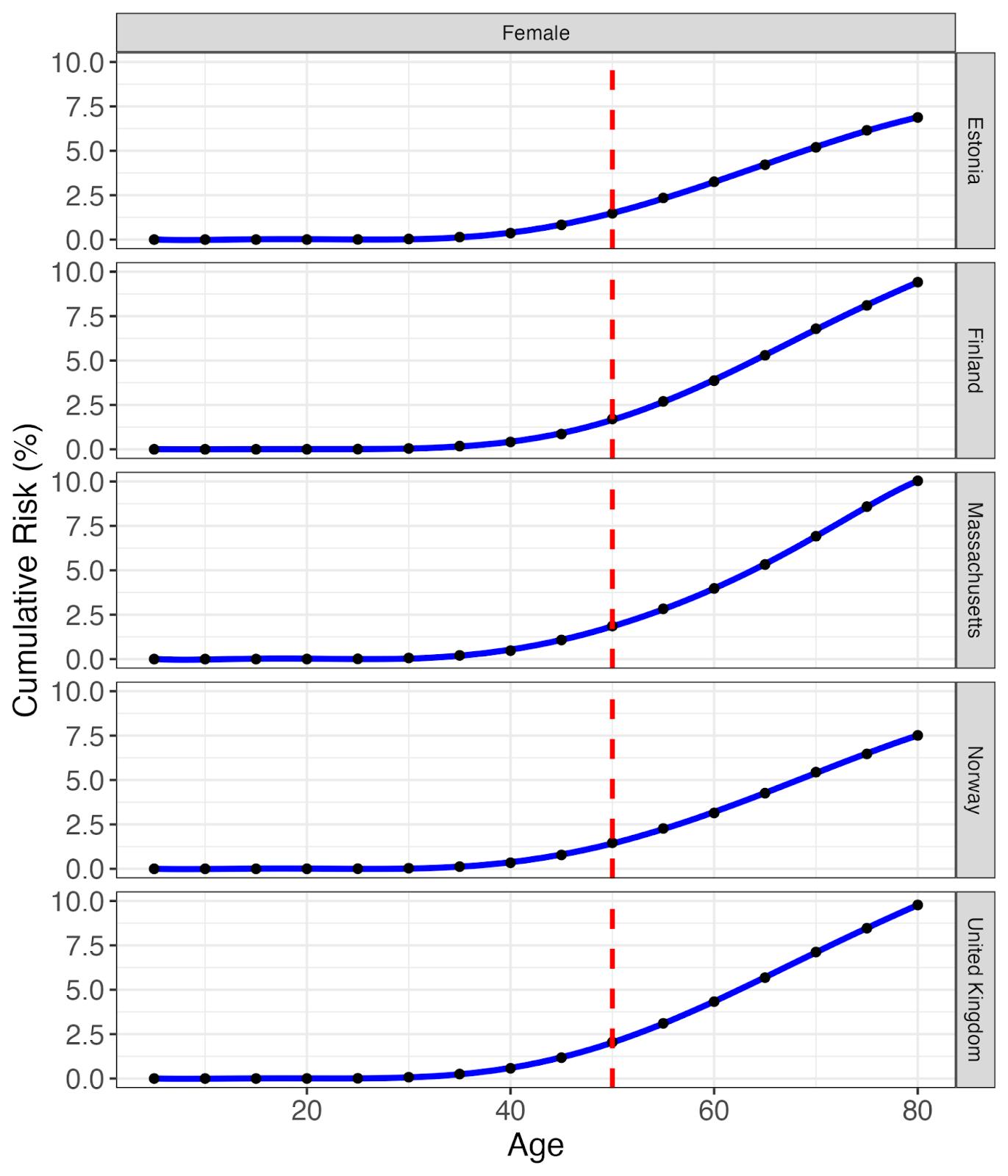

**Supplementary Figure 9.** Country-specific absolute risks for Breast Cancer. Red dashed lines highlight age 50, the age at which screening is recommended. The intersection with the blue cumulative risks were taken as the country's clinical threshold.

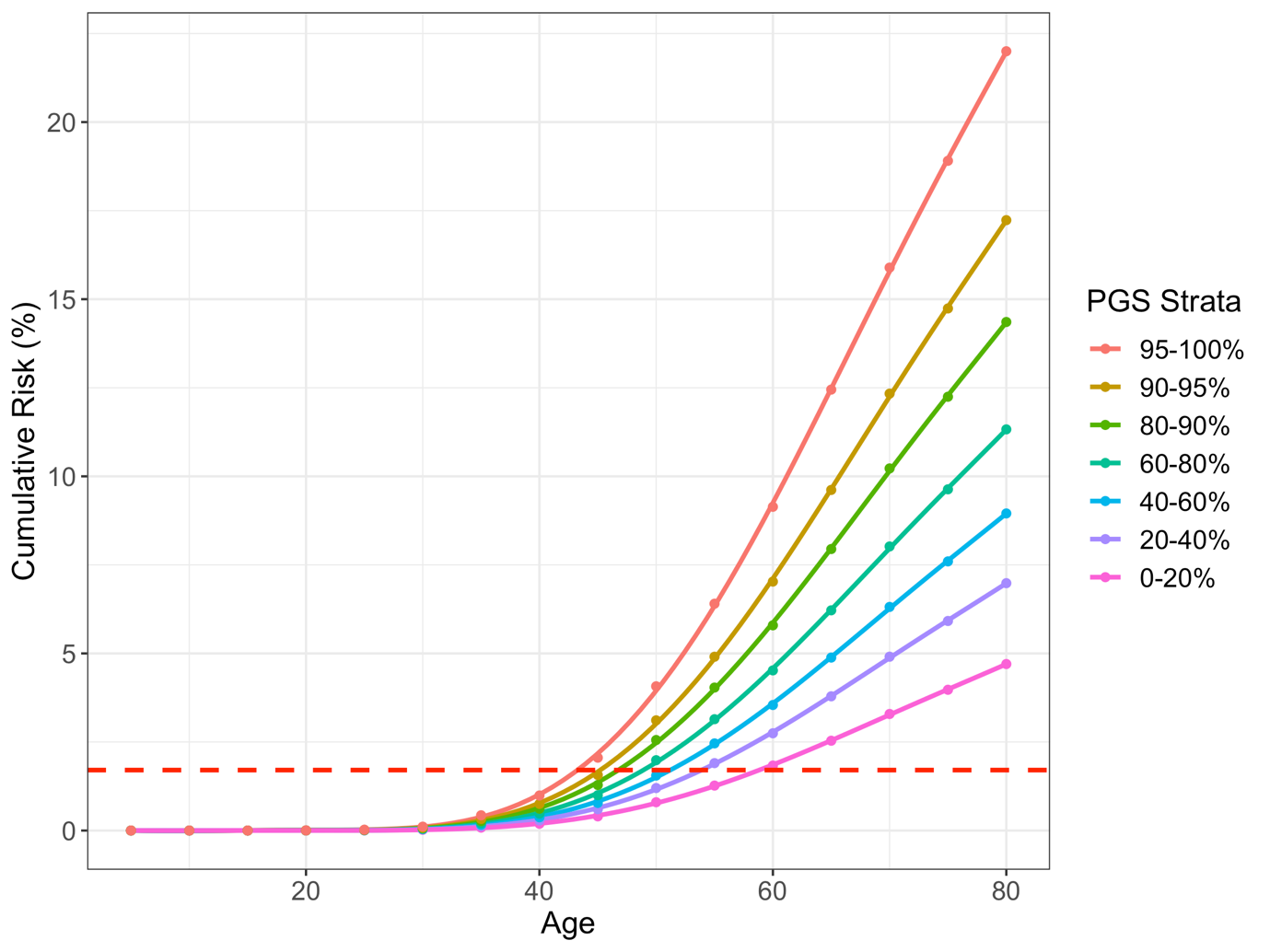

**Supplementary Figure 10.** Breast Cancer cumulative incidence by PGS strata in Finland inclusive of the top and bottom percentiles of risk for breast cancer.

**a)**

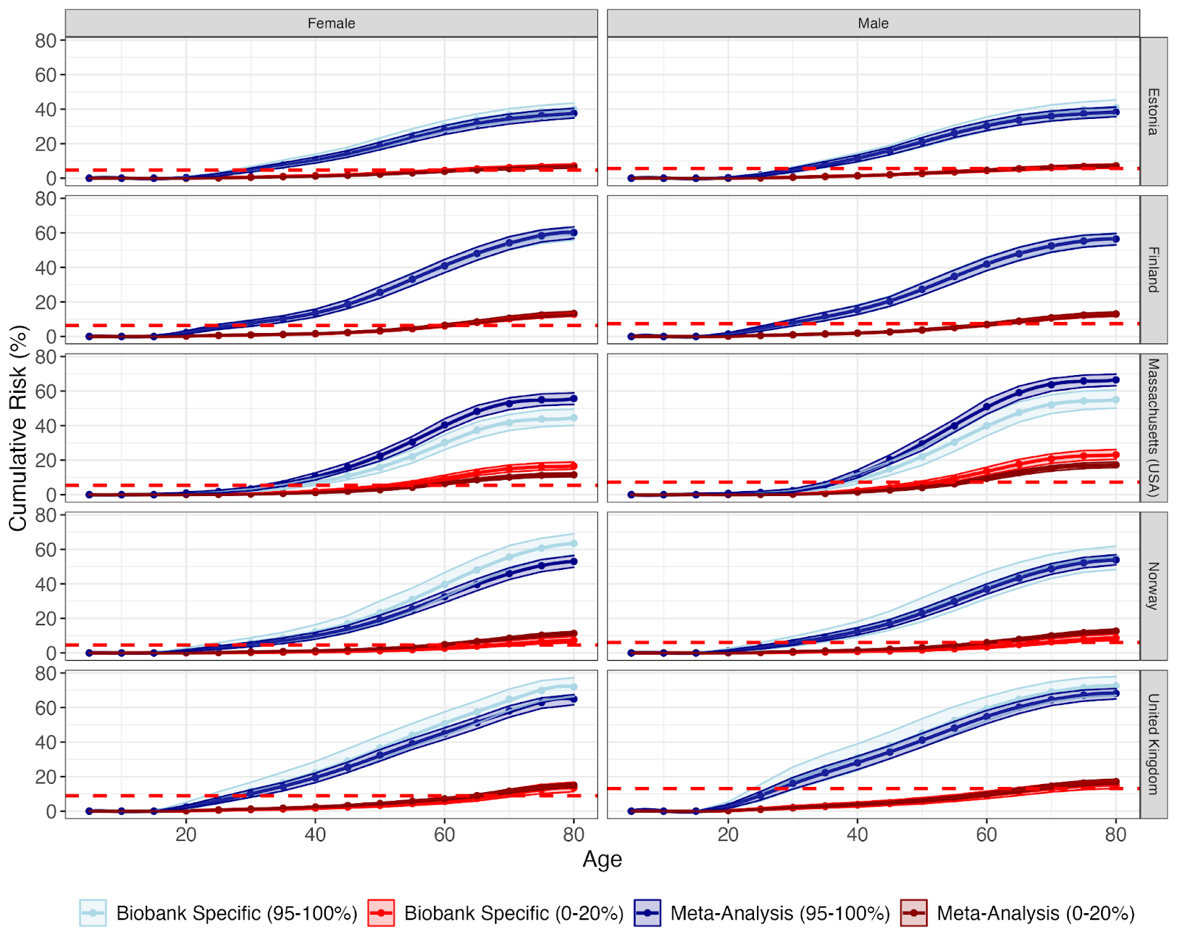

**b)**

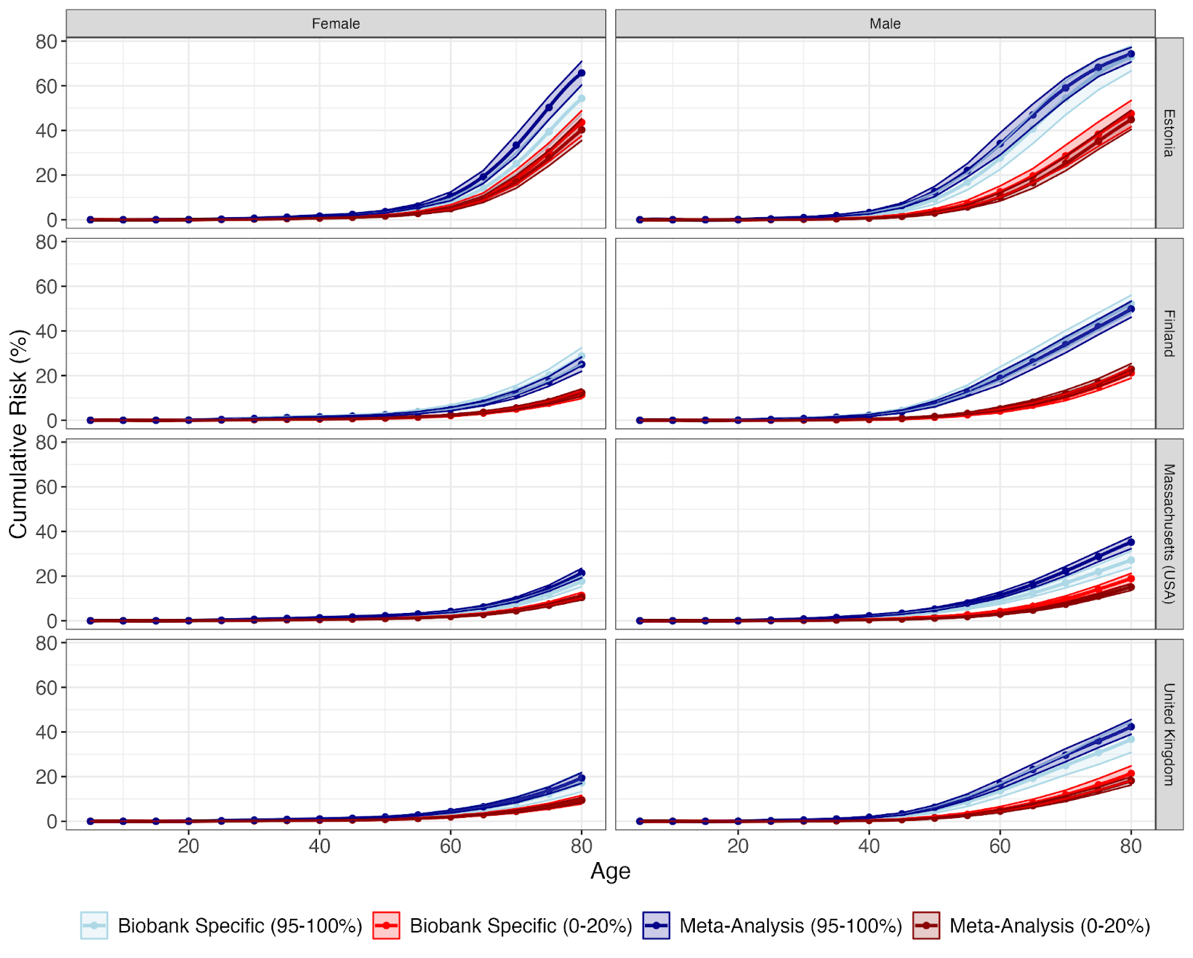

**Supplementary Figure 11.** Comparison of the cumulative incidence estimates resulting from the use of study specific hazard ratios or meta-analysed estimates. **a)** Type 2 Diabetes. **b)** Coronary Heart Disease

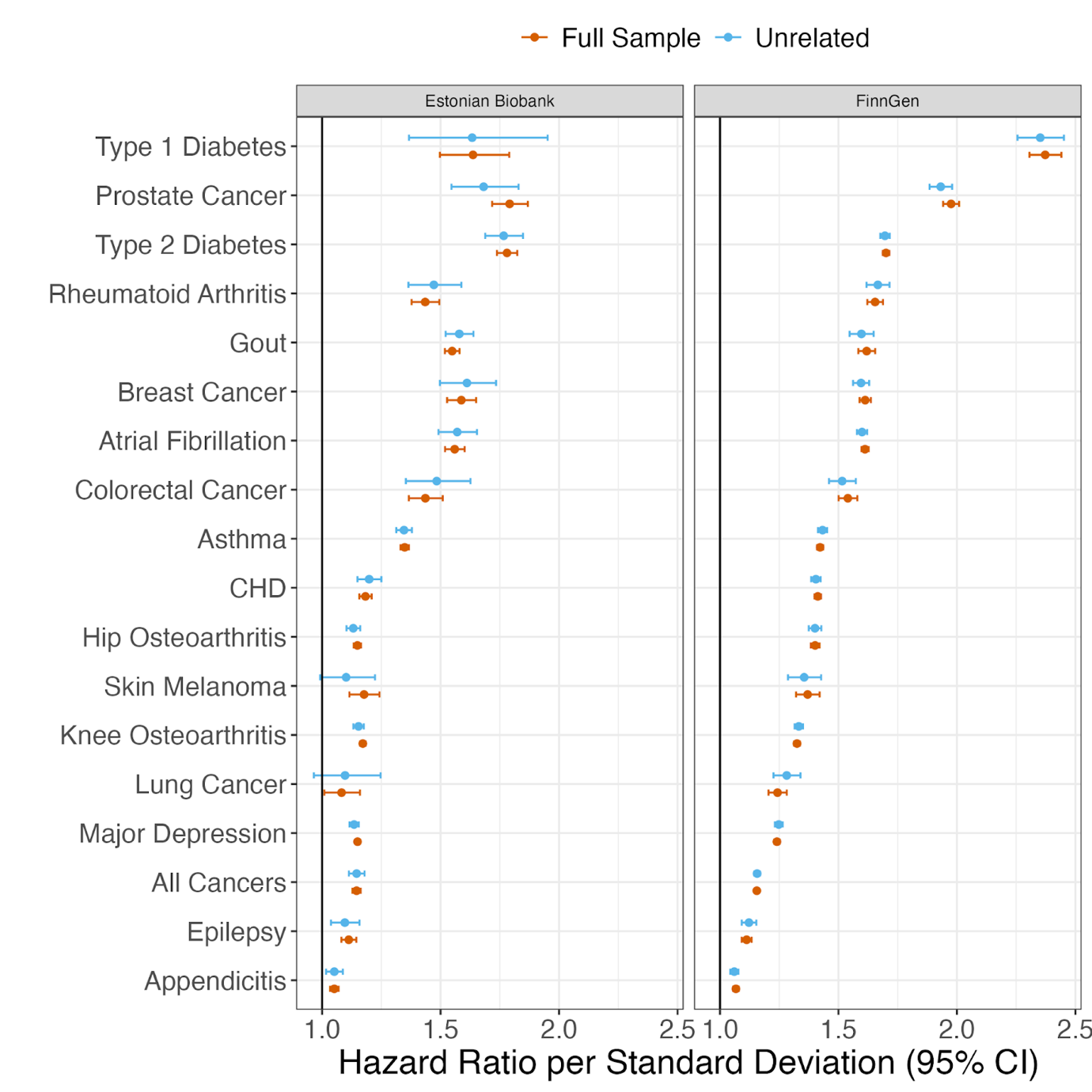

**Supplementary Figure 12.** Sensitivity analysis reviewing the impact of including relatives on the hazard ratios within FinnGen and Estonian Biobank. Third degree relatives and higher were removed and the hazard ratios were compared.

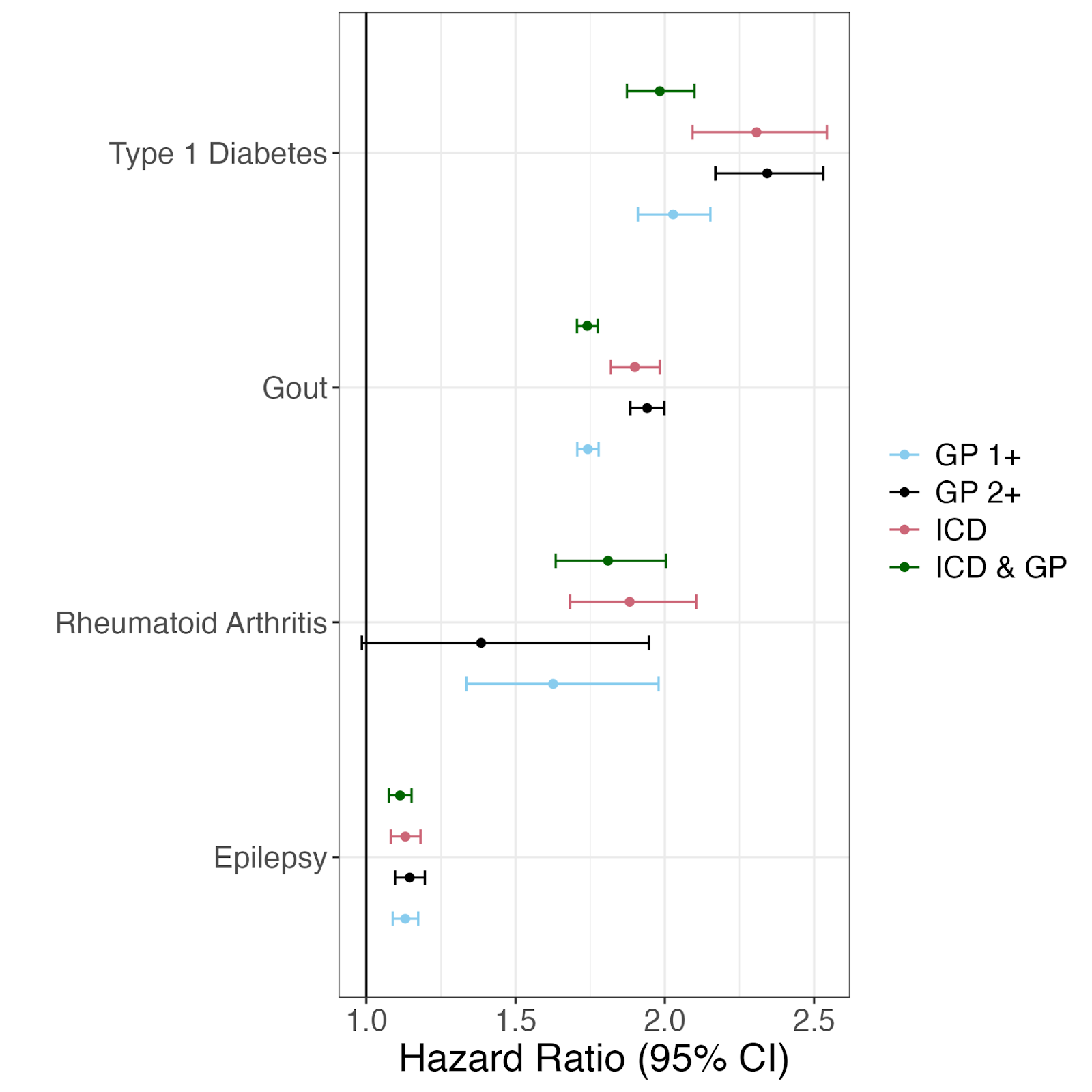

**Supplementary Figure 13.** Sensitivity analysis reviewing the impact of secondary care diagnoses (ICD codes) vs primary care diagnoses (Readv2 and CTV3 codes) on the hazard ratios within the UK Biobank.

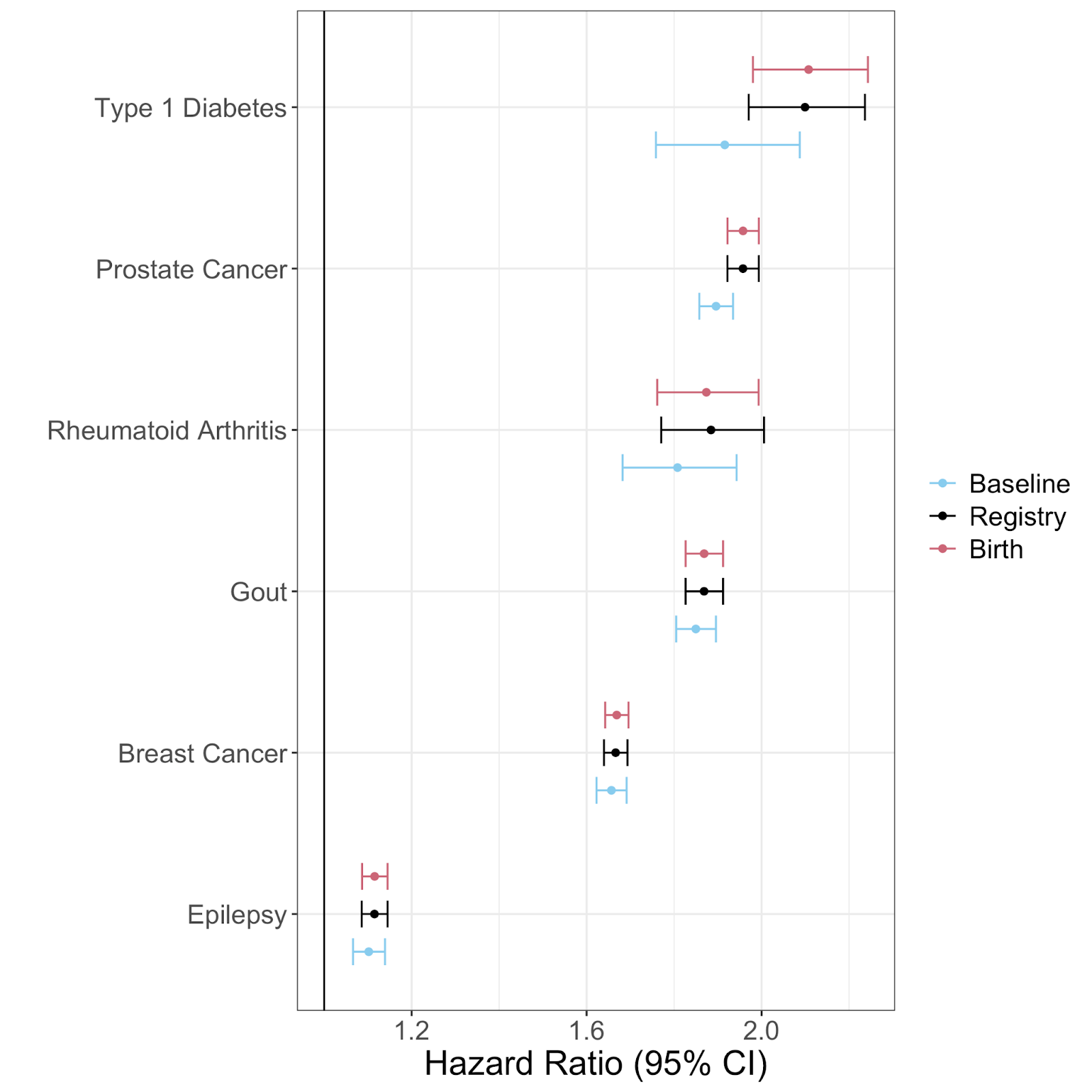

**Supplementary Figure 14.** Sensitivity analysis reviewing the impact of assuming follow-up begins at birth, at the start of the registry linkage or at recruitment (baseline) in the UK Biobank.

**Study co-authors**

We acknowledge the contribution of the Genomics England Research Consortium. The members of this consortium are: John C. Ambrose^1^, Prabhu Arumugam^1^, Roel Bevers^1^, Marta Bleda^1^, Freya Boardman-Pretty^1,2^, Christopher R. Boustred^1^, Helen Brittain^1^, Matt J. Brown^1^, Mark J. Caulfield^1,2^, Georgia C. Chan^1^, Adam Giess^1^, Angela Hamblin^1^, Shirley Henderson^1,2^, Tim J. P. Hubbard^1^, Rob Jackson^1^, Louise J. Jones^1,2^, Dalia Kasperaviciute^1,2^, Melis Kayikci^1^, Athanasios Kousathanas^1^, Lea Lahnstein^1^, Sarah E. A. Leigh^1^, Ivonne U. S. Leong^1^, Javier F. Lopez^1^, Fiona Maleady-Crowe^1^, Meriel McEntagart^1^, Federico Minneci^1^, Jonathan Mitchell^1^, Loukas Moutsianas^1,2^, Michael Mueller^1,2^, Nirupa Murugaesu^1^, Anna C. Need^1,2^, Peter O’Donovan^1^, Chris A.Odhams^1^, Christine Patch^1,2^, Daniel Perez-Gil^1^, Mariana Buongermino Pereira^1^, John Pullinger^1^, Tahrima Rahim^1^, Augusto Rendon^1^, Tim Rogers^1^, Kevin Savage^1^, Kushmita Sawant^1^, Richard H.Scott^1^, Afshan Siddiq^1^, Alexander Sieghart^1^, Samuel C. Smith^1^, Alona Sosinsky^1,2^, Alexander Stuckey^1^, Mélanie Tanguy^1^, Ana Lisa Taylor Tavares^1^, Ellen R. A. Thomas^1,2^, Simon R.Thompson^1^, Arianna Tucci^1,2^, Matthew J. Welland^1^, Eleanor Williams^1^, Katarzyna Witkowska^1,2^, Suzanne M. Wood^1,2^, Magdalena Zarowiecki^1^.

1. Genomics England, London, UK
2. William Harvey Research Institute, Queen Mary University of London, London, EC1M 6BQ, UK
